# Interventions to improve retention in HIV care: a systematic review and network meta-analysis of randomised controlled trials

**DOI:** 10.64898/2026.04.18.26351146

**Authors:** Nadia Rehman, Gordon Guyatt, Mou Jin Jin, Lucas Kallas Silva, Jessica Gu, Mehnaz Munir, Raha Sadagari, Michelle Li, Daniel Xie, Sahith Rajkumar, Yan Lijiao, Elina Najmabadi, Varun Dhanam, Dominik Mertz, Aaron Jones

**Affiliations:** Department of Health Research Methods, Evidence, and Impact, McMaster University, Hamilton, Ontario, Canada; Department of Medicine, McMaster University, Hamilton, Ontario, Canada; MAGIC Evidence Ecosystem Foundation, Oslo, Norway; Department of Pharmacy, Beijing United Family Hospital, Beijing, China; Department of Infectious Diseases, Federal University of São Paulo, São Paulo, Brazil; Michael G. DeGroote School of Medicine, McMaster University, Hamilton, ON, Canada; Mary Heersink School of Global Health and Social Medicine, McMaster University, Hamilton, Ontario, Canada; Department of Pharmacy, Tehran University of Medical Sciences, Tehran, Iran; Department of Integrated Biomedical Engineering and Health Sciences, McMaster University, Hamilton, Ontario, Canada; Institute of Basic Research in Clinical Medicine, China Academy of Chinese Medical Sciences, Beijing, China; Department of Biological Sciences, University of Guelph, Guelph, Ontario, Canada; Department of Biochemistry and Biomedical Sciences, McMaster University, Hamilton, Ontario, Canada

**Author notes:** **Correspondance:** Dr Aaron Jones, Assistant Professor, Department of Health Research Methods, Evidence & Impact McMaster University, Hamilton, Ontario, Canada.

**Keywords:** HIV, people living with HIV (PLHIV), retention, viral load suppression, GRADE, network meta-analysis, randomised controlled trials

## Abstract

**Background:** Sustained retention in care supports continuous access to antiretroviral therapy, routine clinical monitoring, and long-term viral suppression.

**Objective:** To compare the effectiveness of interventions for improving retention in care among people living with HIV (PLHIV).

**Design:** Systematic review and network meta-analysis

**Data sources:** PubMed, Embase, CINAHL, PsycINFO, Web of Science, and the Cochrane Library from 1995 to December 2024.

**Eligibility criteria:** Randomised controlled trials (RCTs) evaluating interventions to improve retention in care, viral load suppression, or quality of life (QoL) among PLHIV, compared with standard of care (SoC) or other interventions.

**Data extraction and synthesis:** Pairs of reviewers independently screened studies, extracted data, and assessed risk of bias using ROBUST-RCT. We conducted a fixed-effect frequentist network meta-analysis and rated interventions categories relative to SoC based on effect estimates effects and the certainty of evidence.. Dichotomous outcomes were summarized as odds ratios (ORs) with 95% confidence intervals (CIs), and continuous outcomes as mean differences (MDs) with 95% CI.

**Results:** Eighty-four trials enrolling 107 137 PLHIV evaluated 13 intervention categories. For retention in care, five interventions supported by moderate or high certainty evidence proved superior to SoC: multi-month dispensing (OR 2.02, 95% CI 1.32 to 3.09), task shifting (OR 1.94, 95% CI 1.42 to 2.66), differentiated service delivery (OR 1.47, 95% CI 1.22 to 1.76), behavioural counselling (OR 1.36, 95% CI 1.21 to 1.54), and supportive interventions (OR 1.31, 95% CI 1.11 to 1.55). For viral load suppression, two interventions supported by moderate or high certainty evidence proved superior to SoC: task shifting (OR 2.07, 95% CI 1.25 to 3.43) and behavioural counselling (OR 1.34, 95% CI 1.11 to 1.67). Across outcomes, no intervention demonstrated convincing superiority over other active interventions.

**Conclusions:** Among 13 intervention categories, only a subset provided moderate or high-certainty evidence of superiority to the standard of care, and no superiority to other interventions. Persistent evidence gaps for key populations, diverse settings, and long-term outcomes support the need for context-sensitive and patient-centred interventions.

**Registration:** PROSPERO CRD42024589177

**Strengths and limitations of this study:**

➢ This systematic review followed Cochrane methods and was reported in accordance with PRISMA-NMA guidelines.
➢ The network meta-analysis integrated direct and indirect evidence to compare multiple intervention categories within a single framework.
➢ Risk of bias and certainty of evidence were assessed using ROBUST-RCT and the GRADE approach for network meta-analysis, respectively.
➢ Some networks were sparse, and limited representation of key populations and long-term follow-up constrained the strength and generalisability of inferences.

## Introduction

HIV remains a serious public health problem, with more than 40.8 million people living with the disease (PLHIV). New infections remain common, with 1.3 million incident diagnoses reported in 2024.^1, 2^ Effective management relies on adherence to antiretroviral therapy (ART) to maintain viral suppression.^3–5^ While global initiatives have expanded access to potent ART regimens,^6^ optimal retention in HIV care remains central to achieving the UNAIDS goal of ending the HIV/AIDS epidemic by 2030.^7^

Retention in HIV care ensures continuity of treatment, improves HIV-related health outcomes, supports longevity, and enhances quality of life (QoL).^8, 9^ Socioeconomic barriers, particularly among key populations, restrict access and contribute to persistent attrition.^10, 11^ Evidence from low– and middle-income settings demonstrates a gradual decline in retention following ART initiation, from 72% at 12 months to 69% at 48 months, across studies conducted between 2015 and 2024.^12^ In high-income settings, such as U.S., cohort data from 2020 to 2023 report 12-month retention of only 50% to 55%,^13, 14^ consistent with global monitoring reports documenting persistent attrition across the HIV care continuum.^15,16^ These patterns underscore the need for robust, evidence-based strategies that sustain long-term engagement in care.^15, 16^

The World Health Organization recommends a broad set of interventions to strengthen retention in HIV care.^17^ Research evaluating these interventions has expanded, yet substantial variability exists across study designs, populations, intervention characteristics, settings, and outcome definitions. Interpreting intervention effects remains challenging, and applicability to clinical settings is often uncertain, leaving ambiguity about which interventions most effectively improve retention in care.^18–20^

Network meta-analysis (NMA) extends conventional meta-analysis by enabling simultaneous comparisons of multiple interventions within a single analytical framework through the integration of direct and indirect evidence.^21, 22^ In HIV research, NMAs have demonstrated particular value for evaluating interventions targeting antiretroviral therapy (ART) adherence in PLHIV and people at risk of HIV.^23^However, adherence-focused NMAs do not directly address retention in care or broader patient-important outcomes such as QoL.^84^

Given the wide range of interventions targeting retention in HIV care and the limited availability of direct head-to-head comparisons, we conducted a systematic review and network meta-analysis of randomised controlled trials (RCTs) to evaluate the effects of interventions on retention in care, viral load suppression, and quality of life among PLHIV, relative to standard of care (SoC) or other interventions. We integrated all available direct and indirect evidence and rate interventions by relative treatment effects and certainty ratings, generating a hierarchy of interventions to improve retention in HIV care.^24, 25^ This approach provides a coherent assessment of which intervention categories demonstrate reliable benefit and highlights key evidence gaps relevant to clinical and policy decision-making.^21^

## Methods

We conducted the review using Cochrane methods,^26^reported it according to PRISMA-NMA,^27^ and rated certainty using the GRADE approach for network meta-analysis.^28^ We registered the protocol prospectively in PROSPERO (CRD42024589177).

## Expert Oversight

A multidisciplinary steering group, including a content expert, a statistician, an HIV specialist, and a methodologist, oversaw the review process. The group refined the research question, reviewed the protocol, classified interventions, prioritised outcome measures, proposed subgroup analyses, evaluated the precision of pooled estimates and certainty of evidence, and recommended baseline risks for calculating absolute treatment effects.

## Eligibility criteria

The review included RCTs that enrolled PLHIV and evaluated at least one retention intervention against standard of care (SoC) or another intervention. We imposed no limits on participant age, study setting, or publication language. We excluded studies that lacked original trial data, secondary analyses that did not preserve randomisation, reviews, conference abstracts, trial registries, and protocols. Trials enrolling mixed-serostatus populations were excluded when data for HIV-positive participants could not be disaggregated.

## Interventions

To maintain network connectivity and enable meaningful comparisons, following classifications from previously published reviews of HIV care, we classified interventions into 13 categories. SoC was defined as the usual clinical care described in each trial. Because SoC varied across trials, we defined enhanced SoC (eSoC) as any intervention that offered additional patient-level support compared to other settings and guidelines.^2, 29^ We aggregated interventions that shared a common theoretical framework into single nodes, irrespective of differences in intensity, timing, or delivery method (Table 1). We excluded studies which were not jointly randomizable. Two independent clinical experts, blinded to study outcomes, reviewed and agreed on all node classifications.

**Table 1.**
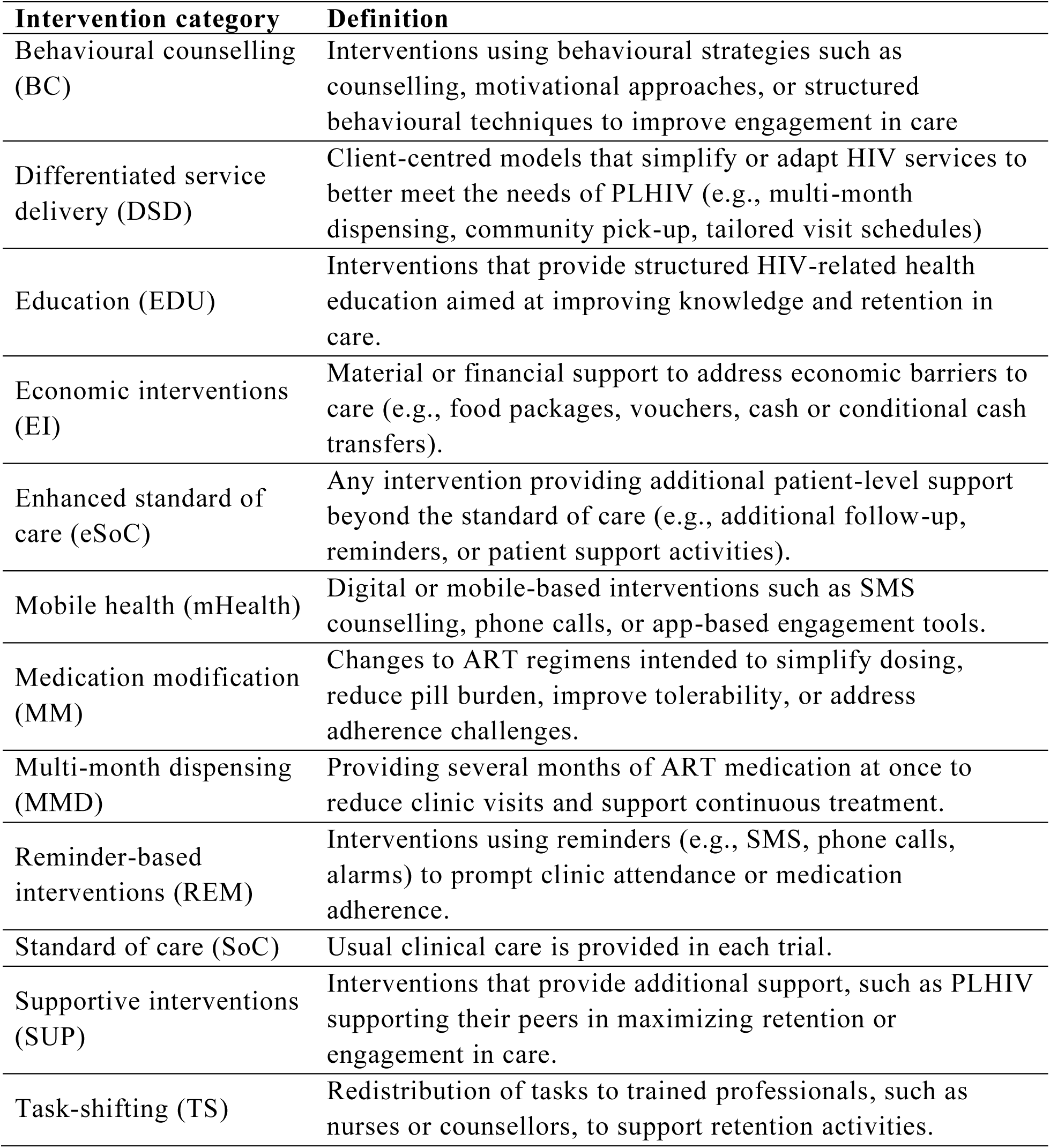
| Definitions used to classify interventions (nodes) in the network meta-analysis.

## Data sources and searches

The search, developed by a health sciences librarian, included Embase (Excerpta Medica Database), CINAHL (Cumulative Index to Nursing and Allied Health Literature), PsycINFO, Web of Science, the Cochrane Central Register of Controlled Trials (CENTRAL), and PubMed (U.S. National Library of Medicine), with no language or geographical restrictions from 1995to December 2024. We screened reference lists of included trials and relevant reviews to identify additional studies. Appendix S1 provides detailed database search strategies.

## Data collection

We downloaded search results and removed duplicates using EndNote X21 (Clarivate, 2023). We developed and pilot-tested screening questions based on predefined eligibility criteria. Pairs of reviewers (MJJ, LKS, JG, MM, RS, ML, DX, SR, YL, EN, VD and NR) independently screened all records in duplicate, first titles and abstracts, and if potentially eligible, full texts. Rayyan, an online systematic review platform, facilitated screening and documentation (Ouzzani et al., 2016).

We extracted data from eligible studies using a standardized, pilot-tested extraction form. The form captured study characteristics (e.g., bibliographic information, country, study design), participant characteristics (e.g., sample size, age, sex, key populations, baseline viral load), intervention characteristics (e.g., theoretical framework, provider, setting, delivery mode, frequency, and tailoring or modification), comparator characteristics, and outcome characteristics (e.g., outcome definitions, measurement tools, and follow-up time points). We classified countries by income level using World Bank income group classifications,^30^ and by geographic region using WHO regional groupings.^31^ We defined key populations as adolescents and young adults (14–24 years), women, African, Caribbean and Black (ACB) populations, people who inject drugs (PWID), and incarcerated individuals (World Health Organization, 2012). Reviewers resolved discrepancies through discussion or, when needed, adjudication by a senior author (AJ).

## Outcomes and follow-up

We assessed three outcomes: retention in care, viral load suppression, and QoL. Retention in care was defined as the proportion of participants who met trial-defined retention criteria and was measured using record-based (objective) or participant-reported (subjective) data.^32^ Viral load suppression was defined as the proportion of participants achieving the trial-defined suppression threshold, with preference for a 40 copies/mL threshold, and was ascertained using laboratory records (objective) or, when unavailable, participant self-report (subjective).^33^ QoL was measured using trial-specified, validated questionnaires and evaluated across four domains: mental health, physical health, general health, and HIV-specific quality of life.

We analyzed participants in the groups to which they were randomized. When trials reported multiple follow-up time points, we pooled effects at 12 months; when a 12-month assessment was not reported, we used the nearest available time point.

## Risk of Bias Assessment

We assessed risk of bias for all reported outcomes using the Risk of Bias Instrument for Use in Systematic Reviews of Randomised Controlled Trials (ROBUST-RCT). We evaluated bias across the instrument’s core domains, including random sequence generation, allocation concealment, blinding, and missing outcome data (>20% missingness classified as high risk).^34^

Given variability in outcome definitions and measurement methods, we also assessed two optional domains: Item 5 (validity of outcome measures), which evaluates bias in outcome measurement, and Item 7 (selective reporting), which evaluates bias from reporting multiple outcome definitions and time points.^34^

Because many interventions could not be blinded, we drew on meta-epidemiological evidence showing that, for objectively measured outcomes, lack of blinding does not systematically bias treatment effect estimates.^35, 36^ For cluster-randomized trials, we additionally assessed identification and recruitment bias using the Cochrane Risk of Bias Tool to evaluate potential bias from imbalances in prognostic factors.^37^

ROBUST-RCT was applied using a two-step evaluation process. First, individual reviewers (MJJ, LKS, JG, MM, RS, ML, DX, SR, YL, EN and VD) evaluated whether methodological safeguards were implemented, rating each item as definitely yes, probably yes, probably no, or definitely no. A senior author (GG) adjudicated unresolved disagreements. Second, a single reviewer (NR) assigned risk-of-bias judgments for each domain and the overall outcome, classifying judgments as definitely low, probably low, probably high, or definitely high.^34^

## Data synthesis and analysis

We used a frequentist framework in R (version 4.4.3) using the netmeta package. Netmeta applies a graph-theoretical, weighted least-squares approach to estimate relative treatment effects.^38^

In sparse networks, random-effects models may produce unstable τ² estimates that overweight small studies, inflate uncertainty, and conflate structural disagreement with statistical uncertainty.^28, 39^ Therefore, given network sparsity, indicated by limited direct evidence across several contrasts, we prespecified a fixed-effect model that assumes negligible between-study heterogeneity (τ²).^40^ The contrast-based model implemented within this framework integrated direct and indirect evidence while accounting for correlations in multi-arm trials, thereby avoiding double-counting.^26, 41^

## Assumptions

Before analysis, we assessed transitivity by comparing the distribution of prespecified effect modifiers (World Bank country income group, population type, WHO geographical regions) across intervention comparisons.^8, 30, 31^ We judged transitivity as plausible based on a graphical assessment of effect-modifier distributions (Figures S1–S3). We assessed global incoherence using the design-by-treatment decomposition of Cochran’s Q statistic and reported its within– and between-design components; given network sparsity, we interpreted the results cautiously.^39^

## Network Visualization and Structural Assessment

We visualized the network using node-and-edge plots and examined connectivity, sparsity, and the distribution of evidence across interventions and populations.^42,43^ We scaled nodes by sample size and weighted edges by the number of contributing trials.^44^We excluded contrasts with fewer than 100 participants because such estimates are unstable and imprecise.^45^

For dichotomous outcomes (retention and viral load suppression), we extracted the number of participants with events and the total number in each group at 12 months, and calculated treatment effects as odds ratios (ORs) with 95% confidence intervals (CIs). For cluster-randomised trials, we used cluster-adjusted estimates when available or applied the effective sample size method when adjustment was not reported.^37^

We synthesized dichotomous outcomes using ORs with 95% CIs. We converted pooled odds ratios to absolute effects using a baseline risk derived from the median control event rate and expressed results as risk differences per 1,000 participants. To maximize precision and comparability across nodes, we selected the most connected intervention as the reference comparator for each network.

For the continuous outcome, QoL, we extracted means and standard deviations (SDs) for each QoL instrument. When SDs were unavailable, we imputed them using values borrowed from similar randomised trials.^46^ For studies reporting medians with ranges or interquartile ranges, we estimated means and SDs using Cochrane-recommended methods.^47^ Because studies used different QoL instruments to measure similar constructs, we standardised all QoL scores to a 0 – 100 scale using linear rescaling procedures consistent with SF-36 domain conventions.^48^ We pooled outcomes as weighted mean differences (WMDs) with 95% confidence intervals (CIs) across studies.^49^ When domains lacked overlapping comparisons or networks were disconnected, we summarized study-level findings descriptively rather than conducting pairwise or network meta-analysis.

## Subgroup analysis

We explored heterogeneity across outcomes using subgroup analyses guided by the Instrument for Assessing the Credibility of Effect Modification Analyses (ICEMAN). ICEMAN assesses the credibility of a subgroup across eight criteria: within-versus between-trial comparisons, consistency of findings, number of contributing trials, pre-specification of hypotheses, statistical interaction testing, the number of effect modifiers examined, use of random-effects models, and the rationale for cut-offs applied to continuous variables. Because formal interaction testing was often not feasible in the NMA due to sparsity, we reported interaction tests from available pairwise meta-analyses. We rated the credibility of each subgroup effect as high, moderate, low, or very low.^50^ If the ICEMAN application indicated low or very low certainty of a possible subgroup effect, we used a single estimate across the comparison.

We predefined nine subgroups to explain potential over– or under-estimation of effects due to effect modification. Subgroup network estimates were limited to direct comparisons.

1. Populations: We hypothesized that the intervention effect on retention would be larger in key populations than in the general population, given lower baseline retention and greater structural barriers to sustained retention in HIV care.^51^
2. Baseline viral load: We hypothesized larger intervention effects among participants with higher baseline viral load, who have greater potential for clinical improvement.^52,53^
3. Setting: In terms of geographical regions and income-level classifications,^30, 31^ we anticipated smaller effects in lower-income countries and in some WHO regions compared with high-income settings, reflecting differences in health-system capacity and contextual constraints.^1,2^ We expected counselling-based interventions to favour high-income settings and supporter, mHealth, and reminder-based interventions to favour lower-income settings.^2^
4. Follow-up duration: We expected intervention effects to attenuate with longer follow-up durations and therefore planned subgroup comparisons at 6 months, 12 months, and beyond 12 months.^2^
5. Outcome measures: We did not expect systematic differences in intervention effects across retention measures or viral load thresholds.^2^ Findings from our qualitative study suggest that outcome measures are often tailored to purpose and context, and such adaptation helps capture the intended construct.^54^
6. Risk of bias: We expected larger observed effects in trials at higher risk of bias, as reduced internal validity is known to be associated with overestimation of treatment effects.^39^
7. Intervention intensity: We hypothesized that higher-intensity interventions would be associated with larger effects, reflecting greater exposure and reinforcement of intervention components.^55^
8. Pragmatism: We assessed trial pragmatism using the Rating of Included Trials on the Efficacy – Effectiveness Spectrum (RITES) tool.^56^We expected larger effects in more explanatory trials, where controlled conditions may amplify observed benefits relative to real-world implementation.^57^
9. Design: We treated study design (cluster vs individually randomised) as an exploratory subgroup. Although design features may influence variance, contamination risk, and implementation fidelity, there is no empirical evidence that cluster randomisation systematically modifies the underlying intervention effect; therefore, no directional hypothesis was prespecified.^58^

## Certainty of evidence

We assessed the certainty of evidence for each network estimate using the GRADE (Grading of Recommendations, Assessment, Development, and Evaluation) framework, classifying evidence as high, moderate, low, or very low.^24^ We used the null (no-effect) line as the decision threshold and based certainty ratings on evidence indicating a non-zero effect.^59^ One reviewer (NR) with formal training in GRADE methodology assessed the certainty of evidence. Any uncertainties were discussed with and adjudicated by a senior author (GG).

## Assessment of direct estimates

We first evaluated certainty across direct comparisons by assessing four domains using the Core Grade Approach: risk of bias (limitations in study design or conduct), inconsistency (heterogeneity of effect estimates), indirectness (differences in populations, interventions, or outcomes relative to the network target), and publication bias.^60^When a direct estimate contributed ≥ 80% of the network weight, we based certainty ratings primarily on the direct evidence, otherwise, we proceeded to assess the certainty of the indirect estimate.

## Assessment of indirect estimates

For each indirect estimate, we assigned the lowest certainty rating of the direct comparisons contributing to the most influential first-order loop in the network. When no first-order loop was available, we based our certainty ratings on the relevant direct evidence. We rated down for intransitivity when the direct comparisons informing the indirect comparison differed in population characteristics with high or moderate credibility using ICEMAN criteria.^61^

## Assessment of NMA estimates

For each network estimate, we began with the certainty from the evidence source (direct or indirect) contributing the highest weight. If both direct and indirect estimates contributed similar weight to the network estimate, we used the higher certainty of the direct and indirect.^28^

We examined incoherence between direct and indirect evidence using a cautious, stepwise approach given the sparsity of the network. First, we applied the SIDDE test and considered a p-value < 0.01 as evidence of potential incoherence. When the test suggested potential incoherence (p < 0.01), we conducted a ratio-of-ratios (RoR) test, where a value of 1 denotes perfect agreement, and a wide 95% confidence interval reflects low statistical power rather than true incoherence for a given comparison.

When only second-order indirect evidence contributed, with no direct evidence or first-order loops, we did not assign a certainty rating.^28, 40^

We judged network estimates as imprecise if the associated 95% confidence intervals included our decision threshold “null”.

## Categorization of interventions

We rated interventions relative to the SoC, based on the magnitude of the NMA estimates, ordered from most to least effective. We further distinguished interventions with high or moderate GRADE certainty as “convincingly effective” and those with low or very low certainty as “possibly effective.” We presented ratings using a gradient color-coding framework: green categories denote high or moderate certainty, whereas grey categories denote low or very low certainty. We verified that ratings were consistent with pairwise results and P-scores.^25, 40^

## Results

Of 4,826 records identified through the literature search, after title and abstract screening, we assessed 3,922 full-text records for eligibility and identified 96 randomised trials enrolling 107,137 PLHIV. We included 84 trials in the network meta-analysis; Figure 1 presents details of the screening process (PRISMA flow diagram). We excluded 12 studies from the quantitative synthesis because their interventions were not aligned with current WHO guidelines or were not comparable to the interventions included in the network with respect to comparators, populations, or intervention structures (Table S1).

**Figure 1.**
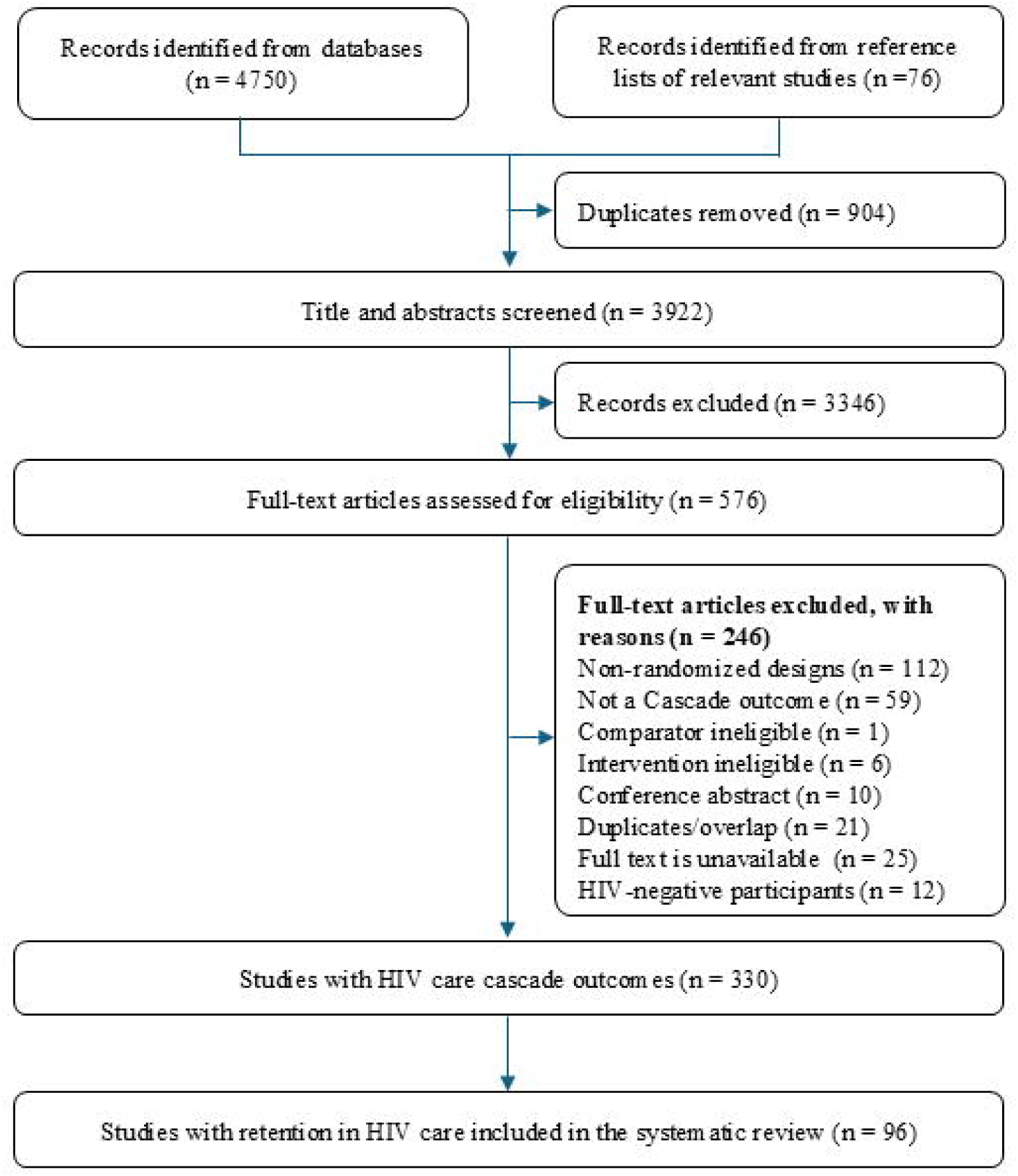
PRISMA flow diagram of selection of randomised controlled trials evaluating interventions to improve retention in HIV care among people living with HIV.

## Study characteristics

Table 2 summarizes study characteristics, with additional details provided in Tables S2-S4. Typical studies enrolled adult participants with similar proportions of males and females. Across World Bank income classifications, most studies were conducted in high-income countries.^30^ Across the World Health Organization geographical regions, most studies were conducted in the WHO African Region.^31^ Studies predominantly used parallel-group, individually randomized designs, and interventions were delivered primarily in clinic-based settings.

**Table 2.** | Characteristics of included studies (84 trials: 107,137 participants)

| **Study characteristic** | **No. (%) of studies/**<br>**Median (Q1–Q3) \*** |
| --- | --- |
| Sample size | 305 (109–623) |
| Female participants | 48 (31–67) |
| **Country income level<sup>†</sup>** |  |
| Low-income | 15 (18.1%) |
| Lower-middle-income | 18 (21.7%) |
| Upper-middle-income | 19 (22.9%) |
| High-income | 32 (38.6%) |
| **Population type<sup>‡</sup>** |  |
| General population | 52 (61.9%) |
| African, Caribbean and Black populations | 9 (10.7%) |
| Men who have sex with men | 2 (2.4%) |
| Incarcerated populations | 4 (4.8%) |
| People who inject drugs | 2 (4.8%) |
| Youth | 8 (9.5%) |
| **WHO geographical region<sup>§</sup>** |  |
| African Region | 44 (53.7%) |
| Region of the Americas | 31 (37.8%) |
| European Region | 5 (6.1%) |
| South-East Asia Region | 2 (2.4%) |
| Western Pacific Region | 1 (1.2%) |
| **Study design** |  |
| Multi-arm | 18 (21.4%) |
| Parallel arm | 66 (78.6%) |
| **Study setting** |  |
| Clinic-based | 40 (47.6%) |
| Community-based | 29 (34.5%) |
| Hybrid or phone-based | 15 (17.9%) |

\*Values are number (%) of studies unless otherwise indicated; continuous variables are reported as median (Q1–Q3)
<sup>†</sup> Country income level classified according to the World Bank country income classification
<sup>‡</sup> Population categories are not mutually exclusive; totals may exceed 100%.
<sup>§</sup> WHO geographical regions are defined according to the WHO regional classifications

## Risk of Bias

### Retention in HIV care

The 84 eligible studies reported 87 retention outcomes, with three studies assessing retention using multiple measures. Overall, 21 studies (24.0%) were judged to have a definitely low risk of bias and 36 (41.4%) a probably low risk, whereas 26 (31.0%) were rated as probably high or definitely high risk, primarily due to missing outcome data. Outcome measurement was objective in most studies, with 74 (88.1%) judged to be definitely or probably low risk.

## Viral Load Suppression

Among the 49 studies reporting viral load outcomes, 29 (59%) were judged to be at high risk of bias, primarily due to missing data exceeding 20%. Two trials (Mbuagbaw et al., 2012 and Lucas et al., 2010) did not report viral load outcomes because attrition exceeded 90%.^62, 63^

## Quality of Life

Six studies reported QoL outcomes; 4 (66%) were judged to be at high risk of bias, primarily due to lack of blinding and reliance on self-reported questionnaires (4/6, 66%). Missing outcome data further contributed to bias, with five of six studies (83%) rated as high risk on this domain. In one study (Willis et al., 2019), incomplete reporting of the QoL instrument increased uncertainty about outcome measurement validity(1/6, 17%).^64^ Evidence across QoL domains was sparse, with most comparisons based on single studies, limiting confidence in pooled inference (Table S18-S20).

We summarized the risk-of-bias assessments in Figures S4–S6, generated using the ROBUST-RCT Figure Generator (MAGIC Evidence Ecosystem Foundation; https://magicevidence.org/robust-rct).

## Findings per outcome

### Retention in care

The network meta-analysis synthesized evidence from 84 trials across 13 intervention categories. The network had a hub-and-spoke structure anchored on SoC, with several multi-arm trials. MM and multi-month dispensing (MMD) were only connected to SoC (Figure 2).

**Figure 2.**
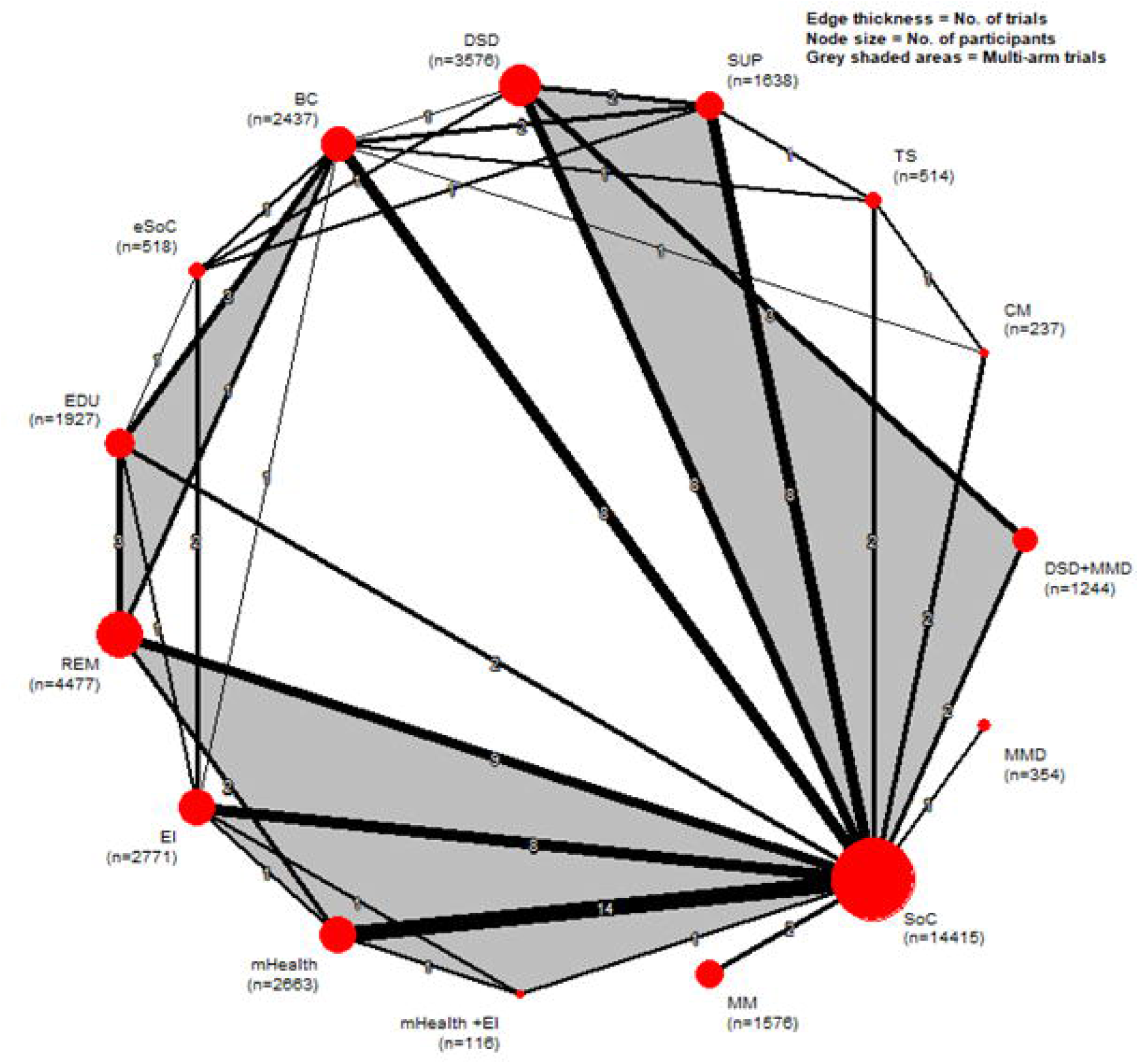
Netgraph of interventions for retention in care.

We included 84 trials that enrolled 107,137 PLHIV and assessed 13 intervention categories. For retention in care, no intervention proved better than any others. Five interventions proved superior to SoC with moderate or high certainty: MMD, task shifting (TS), differentiated service delivery (DSD), behavioral counselling (BC), and supporter (SUP). Among interventions supported by low or very low certainty evidence, only economic incentives (EI) proved superior to SoC Table 3 presents the categorization of interventions for retention in HIV care and viral load suppression. The league table displays all head-to-head comparisons (Table S14). Figure 3 provides the forest plot of network estimates of interventions against SoC.

**Figure 3.**
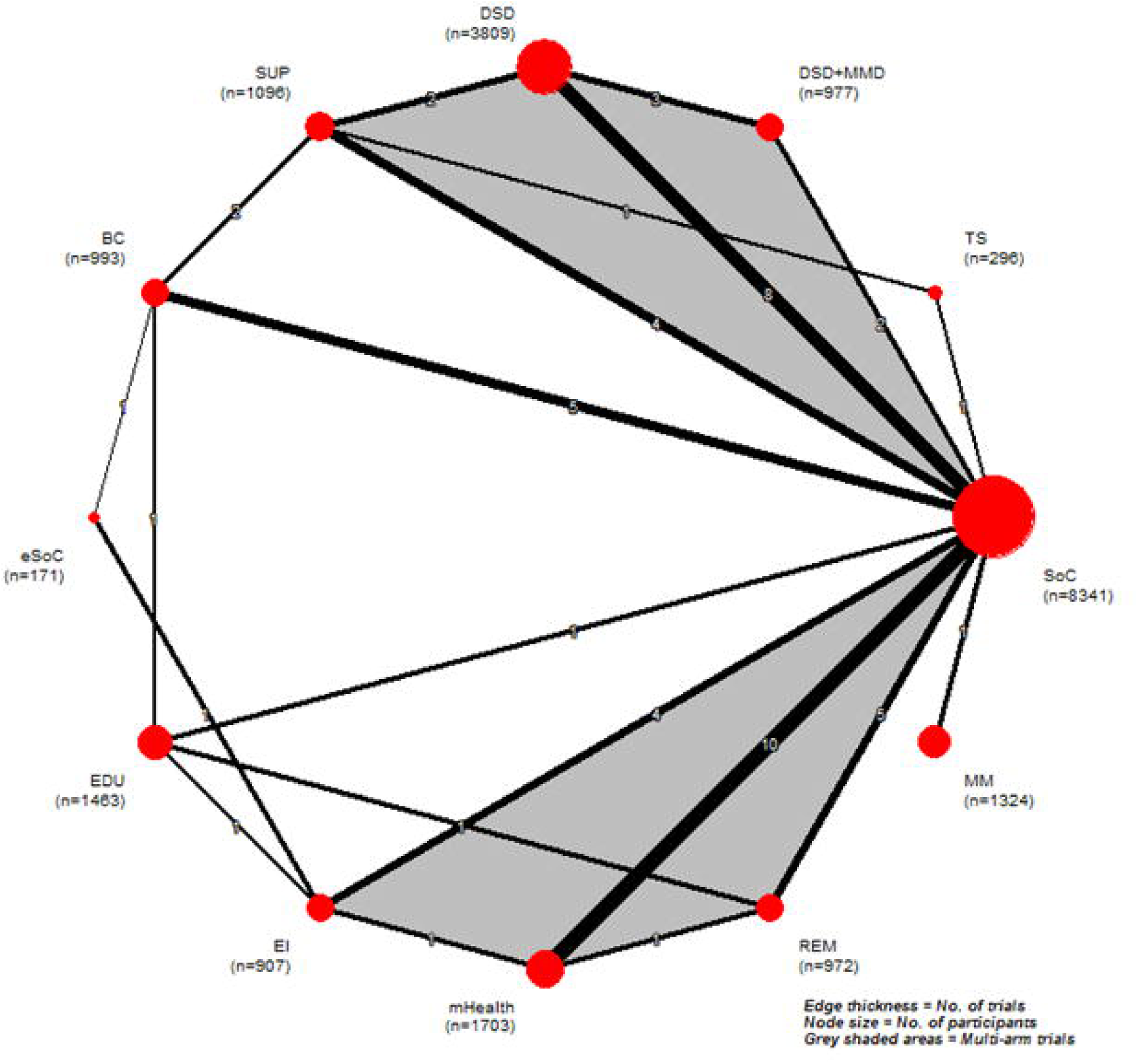
Forest plot of network meta-analysis estimates comparing each intervention with SoC using a fixed-effects model.

**Table 3.** | Network meta-analysis results, sorted by GRADE certainty of evidence for retention in care and viral load suppression. Table 3 | Gradient colour coding; no evidence =’-’; Reference group = standard of care (SoC); bold = statistically significant (95% confidence intervals that do not include 1). All ORs and RDs per 1000 are fixed-effect NMA mixed estimates from netsplit; OR odds ratio; CI, confidence interval; RD: risk difference; Baseline: median control-group risk

| Intervention | Retention in HIV care |  | Viral load suppression |  |
| --- | --- | --- | --- | --- |
|  | OR<br>(95% CI) | RD<br>560 per 1000<br>patients | OR<br>(95% CI) | RD<br>530 per 1000<br>patients |
| MMD | **2.02**<br>**(1.32 to 3.09)** | **160**<br>**(67 to 237)** | - | - |
| TS | **1.94**<br>**(1.42 to 2.66)** | **152**<br>**(84 to 212)** | **2.07**<br>**(1.25 to 3.43)** | **170**<br>**(55 to 265)** |
| DSD | **1.47**<br>**(1.22 to 1.76)** | **92**<br>**(48 to 131)** | **1.34**<br>**(1.18 to 1.52)** | **72**<br>**(41 to 102)** |
| BC | **1.36**<br>**(1.21 to 1.54)** | **74**<br>**(44 to 102)** | **1.36**<br>**(1.11 to 1.67)** | **75**<br>**(26 to 123)** |
| SUP | **1.31**<br>**(1.11 to 1.55)** | **63**<br>**(23 to 104)** | **1.43**<br>**(1.16 to 1.75)** | **87**<br>**(37 to 134)** |
| mHealth + EI | 1.21<br>(0.82 to 1.81) | 46<br>(-49 to 137) | - | - |
| REM | 1.15<br>(1.00 to 1.32) | 34<br>(0 to 67) | 0.96<br>(0.73 to 1.25) | -10<br>(-78 to 55) |
| EDU | 1.12<br>(0.94 to 1.34) | 28<br>(-15 to 70) | 0.92<br>(0.71 to 1.21) | -21<br>(-85 to 45) |
| DSD + MMD | 1.09<br>(0.72 to 1.63) | 21<br>(-82 to 115) | **0.58**<br>**(0.36 to 0.94)** | **-135**<br>**(-241 to -15)** |
| MM | **0.7**<br>**(0.55 to 0.88)** | **-89**<br>**(-148 to -32)** | 0.93<br>(0.71 to 1.22) | -18<br>(-85 to 49) |
| EI | **1.33**<br>**(1.13 to 1.55)** | **69**<br>**(30 to 104)** | 1.13<br>(0.84 to 1.52) | 30<br>(-44 to 102) |
| CM | 1.49<br>(0.99 to 2.25) | 95<br>(-2 to 181) | - | - |
| mHealth | 1.06<br>(0.93 to 1.21) | 14<br>(-18 to 44) | **0.56**<br>**(0.55 to 0.57)** | **-143**<br>**(-147 to -139)** |
| eSoC | **0.71**<br>**(0.51 to 0.97)** | **-85**<br>**(-166 to -8)** | **0.57**<br>**(0.33 to 1.00)** | **-139**<br>**(-259 to 0)** |

Table 3 | Gradient colour coding; no evidence = '-'; Reference group = standard of care (SoC); bold = statistically significant (95% 819 confidence intervals that do not include 1).
| BENEFIT OUTCOMES |  |
| --- | --- |
| High/Moderate Certainty Evidence | Low/Very Low Certainty Evidence |
| Convincingly superior to the SoC, but no better than any other interventions | Possibly superior to the SoC, but no better than other interventions |
| No apparent difference from SoC | No apparent difference from SoC |
| Convincingly worse than SoC | Possibly worse than SoC |

Global incoherence was detected under the fixed effect design-by-treatment but not after modelling heterogeneity (Q 25.61; df 26; p = 0.48) (Table S21). The net heat plot showed small, local clusters of inconsistencies with no network-wide pattern (Figure S13).

Across the 43 direct comparisons contributing to the network, most contrasts were informed by one or two trials. Only six comparisons had more than five contributing trials (mHealth vs SoC, DSD vs SoC, REM vs SoC, BC vs SoC, SUP vs SoC, and EI vs SoC). Pairwise results aligned directionally with the NMA, reinforcing the coherence between direct and indirect evidence.

Table S5 provides the results of pairwise meta-analysis, and Figures S7–S25 provide the pairwise forest plots.

Supplementary File 3 provides the direct, indirect, and network estimates along with their absolute effects, and the certainty of the evidence for each comparison.

## Subgroup findings

We reported the ICEMAN assessment only for the subgroup with an interaction p-value < 0.10.

1. WHO geographical regions. mHealth, REM, and SUP favoured the WHO African Region over the Region of the Americas; whereas BC favoured SoC across regions (ICEMAN credibility: low).
2. Country income level. mHealth, REM, and SUP versus SoC tended to favour lower-income settings over upper-middle– and high-income settings; BC and EI versus SoC showed similar directions across income groups (ICEMAN credibility: low)
3. Risk of bias. Observed effects were larger in studies at lower risk of bias and closer to the null in higher-risk studies (ICEMAN criteria: very low credibility)
4. Intervention intensity. Higher intensity did not consistently increase effects; only case management and reminder-based strategies suggested larger effects (ICMAN criteria: very low credibility).
5. Pragmatism (RITES). Trials with mid-range pragmatism scores sometimes showed larger effects; “Effective” trials were often near the null; “Efficacious” and “Definitely effective” trials were sparse and inconsistent (ICEMAN criteria: very low credibility).
6. Study design. Effects tended to be larger in parallel-group trials and attenuated in cluster-randomised trials (ICEMAN criteria: very low credibility).

Because none of the analysed prespecified modifiers achieved moderate or high credibility, we had no concerns about intransitivity. The ICEMAN assessment appears in Appendix S2, and detailed subgroup results are provided in Tables S24–S31.

## Viral load suppression

We excluded Metsch et al., 2021 because the case management arm (CM) (n = 15 of 26) provided limited precision and insufficient information for inclusion in the network.^65^ Of the remaining 46 studies, in the cluster trial by Inghel et al., 2024, we collapsed mHealth plus EI into mHealth to reduce network sparsity (Figure 4).^66^

**Figure 4.**
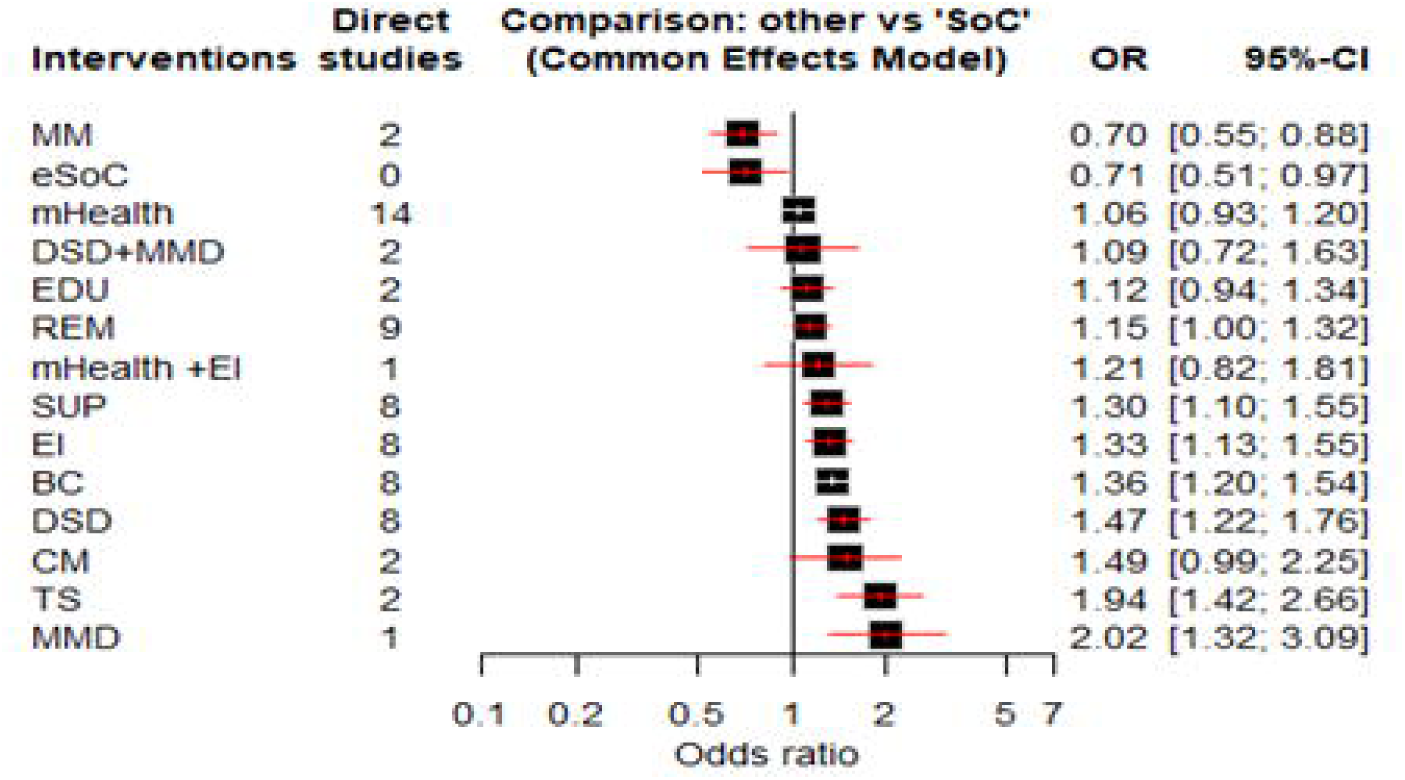
Netgraph of interventions for viral load suppression

For viral load suppression, two interventions TS and BC supported by moderate or high certainty evidence proved superior to SoC. Global incoherence appeared under the fixed-effect design-by-treatment test but resolved after modelling heterogeneity (Q 10.04; df 15; p = 0.817) (Table S22).

(Table 3). The league table displays all head-to-head comparisons (Table S15). Figure 5 visualizes the network estimates versus SoC.

**Figure 5.**
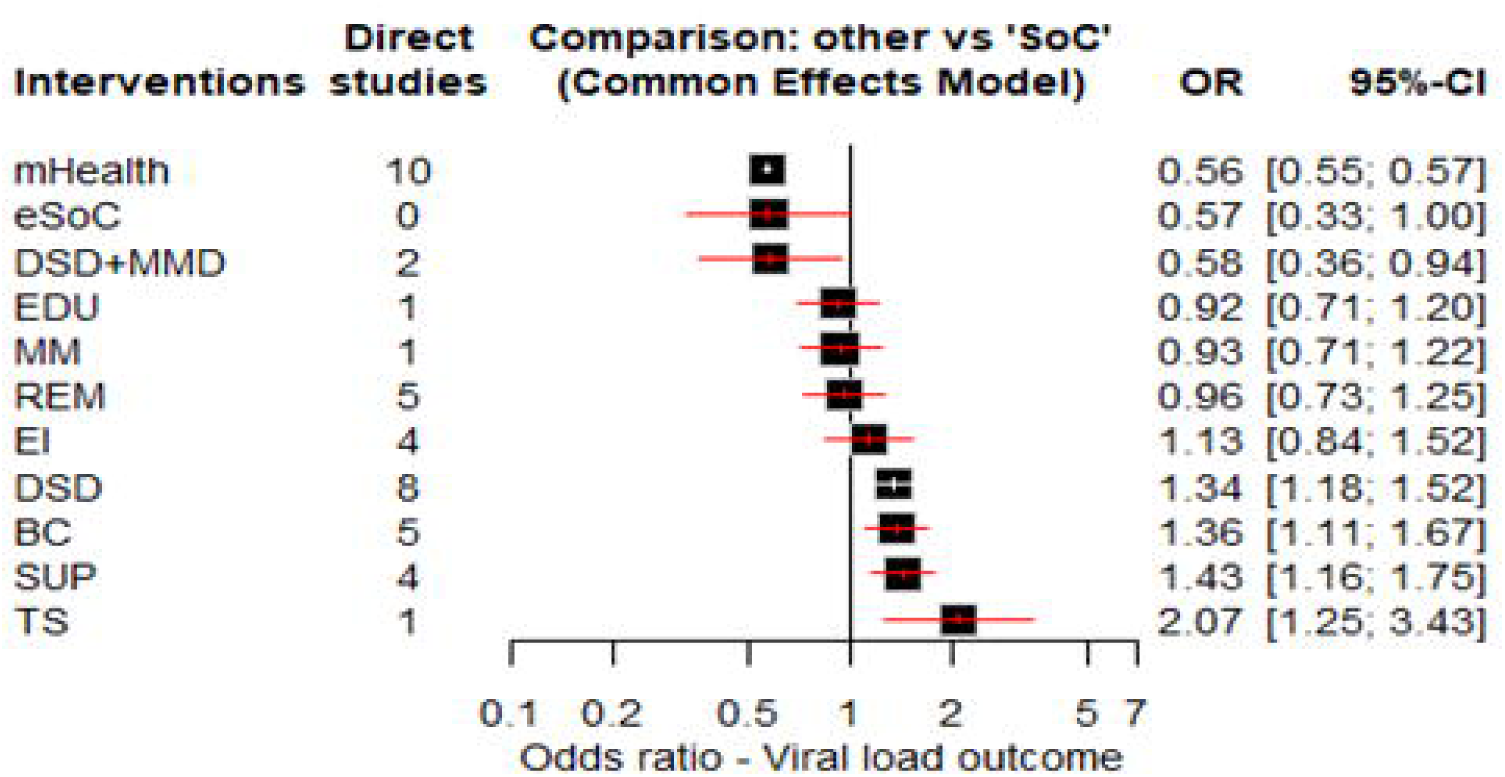
Forest plot of network meta-analysis estimates comparing each intervention with SoC using a fixed-effects model.

## Pairwise vs network results

Pairwise meta-analyses included 17 direct comparisons (Table S13). Across the direct comparisons contributing to the viral load network, most contrasts were informed by one or two trials, and only a small number had more than two contributing studies (DSD vs SoC, mHealth vs SoC, BC vs SoC, EI vs SoC). Pairwise results aligned directionally with the NMA, reinforcing coherence between direct and indirect evidence. Table S13 provides the direct pairwise estimates and the heterogeneity statistics.

## Subgroup analysis

Across all subgroup analyses, none of the prespecified effect modifiers demonstrated moderate or high credibility, and we observed no meaningful differences in treatment effects. Detailed subgroup results are presented in Tables S32–S37.

## QoL

QoL evidence was sparse, with only one study contributing data per comparison in each domain. Estimates were small, imprecise, and inconsistent in direction, so neither pairwise nor network meta-analysis was possible. Individual study results by domain are provided in Tables S11–S13.

## Discussion

This is the first systematic review and network meta-analysis to compare all interventions aimed at improving retention in HIV care. We included 84 randomised trials and assessed three outcomes: retention, viral load suppression, and QoL. Compared with SoC, MMD, TS, DSD, BC, and SUP performed better for retention in care, with TS and BC also demonstrating improved viral load suppression. No intervention proved convincingly better than any other intervention. Using network meta-analysis, we compared multiple complex interventions by integrating direct and indirect evidence within a single analytical framework.

## Strengths and limitations

This review has several strengths. We conducted a comprehensive, up-to-date search with broad inclusion criteria and no language restrictions. To address substantial clinical and methodological heterogeneity across populations, interventions, settings, outcome definitions, and follow-up durations, we performed prespecified subgroup analyses and used ICEMAN to assess the credibility and implications of effect modification.

To preserve clinical and decision relevance, we grouped treatments into conceptually coherent nodes. For complex behavioural interventions, a component NMA may provide richer information, but the sparseness of the current data precluded applying the approach.^21, 67^

We further strengthened inference by applying GRADE to network meta-analysis and categorizing interventions by both effect magnitude and the certainty of the evidence. Our prespecified analytical model was another strength: large SoC-anchored trials dominated key comparisons; therefore, the pre-specified fixed-effect analyses avoided overweighting small studies.^22, 24^ Finally, multidisciplinary input from clinical experts and methodologists enhanced the clinical relevance and methodological rigour.

The limitations of this review largely reflect those of the included primary studies. Although we assessed three patient-important outcomes, only 46 trials reported viral load suppression, and six reported QoL. As HIV is now managed as a chronic condition, QoL is increasingly prioritised to capture health, mental, and social outcomes among people living with HIV;^68^ however, limited data and inconsistency across instruments and domains used in the studies disconnected the network and precluded pooling.

Key populations at highest risk of poor retention were underrepresented; few trials targeted these groups, and within-study evidence proved sparse, limiting the evidence base for targeted approaches to meet their unique needs.^69,70^ Several geographical regions were also under-represented, and because suboptimal retention reflects socio-economic and contextual factors, this gap further limits generalisability.^71^Page 25

Most studies assessed retention only within the first 12 months. Primary studies did not report potential effect modifiers such as baseline viral load.^53^ Moreover, intervention tailoring to key populations and context can be an important effect modifier in diverse settings and populations with diversified needs, but few trials described intervention tailoring, limiting our ability to evaluate adaptive or personalised approaches.^72^

In relation to prior work, we used network meta-analysis to synthesize evidence from randomized trials evaluating interventions to improve retention in HIV care. In contrast to previous reviews, which were limited by observational study designs, narrow intervention scopes, or narrative synthesis, we integrated evidence across a broad range of interventions within a single network, enabling simultaneous direct and indirect comparisons.^18, 20, 73, 74^ Previous reviewers often did not pool the evidence due to the heterogeneity of the measures.^75–78^ In our subgroup analysis, we found that retention measures and viral load thresholds were not effect modifiers, allowing us to pool the studies. Our approach supported coherent node classification across trials and a uniform selection of follow-up time points, and enabled assessment of population, setting, follow-up duration, and intervention intensity as potential effect modifiers. Though we conducted various subgroup analyses, the very low credibility of these findings and heterogeneity across trials indicate that these results are only hypothesis-generating. Thus, in contrast to prior work, we provide a comprehensive quantitative synthesis to inform comparative effectiveness, decision-making, and identification of evidence gaps.

Our findings are consistent with a prior systematic review and network meta-analysis showing that behavioural and counselling interventions improve engagement-related outcomes in HIV care.^2, 79^ Differences in intervention classification and the limited evidence base in the earlier review restricted direct comparability, but supporter-based interventions showed benefits in both reviews. Both reviews found no convincing benefit of reminder-based interventions compared with SoC.^2^

Although mHealth strategies have often been considered promising, robust evidence demonstrating consistent improvements in clinical outcomes remains limited.^80^ In our review, which included 14 studies assessing mHealth interventions for retention in care and nine for viral load suppression, these interventions did not convincingly outperform standard of care for either outcome. Taken together, these findings suggest that interventions addressing behavioural, supportive, and service delivery components (TS, SUP and DSD) may play a more substantial role in improving retention in HIV care.

## Implications for practice and research

Future research should focus on improving the quality of primary studies. Effective implementation requires attention to context, target populations, intervention complexity, and clinical applicability. Trials should prioritize underrepresented groups such as adolescents, young adults, and people who inject drugs, with strategies tailored to their structural and contextual barriers and informed by stakeholder input.^71, 81^ Most trials assessed retention over 6–12 months, despite retention being a lifelong process marked by patients cycling in and out of care; future studies should extend follow-up beyond 24 months and explicitly capture cycling in and out of care, especially in low– and middle-income settings.^17^

Standardized reporting of baseline viral load and clearer descriptions of intervention tailoring and adaptation are needed to support informative assessment of effect modifiers. Incorporating patient-centred outcome measures, including QoL and tools such as the HIV Index, will support a more comprehensive understanding of retention that aligns with evidence highlighting the role of social determinants and the need for equity-oriented care.^82^ Because quantitative effects alone cannot explain why interventions succeed, future trials should embed implementation frameworks and qualitative methods to assess feasibility, acceptability, and contextual adaptation, especially for multicomponent and digital strategies.^83, 84^

## Conclusion

This systematic review and network meta-analysis provide an up-to-date assessment of interventions to improve retention in HIV care. DSD, EI, EDU, and BC emerged as the most effective strategies, and BC also improved viral load outcomes. Evidence remains limited and of variable quality, and heterogeneity in intervention design may have influenced the results. Persistent gaps for key populations, diverse settings and long-term outcomes highlight the need for targeted, context-sensitive approaches.

## Declarations

## Supporting information

Supplementary file 2. Appendixes and tables

Supplementary File 1. Prisma checklist

## Acknowledgment

We thank the health sciences librarian for their guidance and support in developing and refining the electronic search strategies for this review.

## Patient and public involvement

Patients and the public were not involved in the design, conduct, reporting, or dissemination plans of this research.

## Ethics

Ethics approval was not required as this study synthesised data from previously published studies.

## Consent for publication

Not applicable.

## Authors’ contributions

N.R., A.J., G.G. and D.M. contributed to the review’s conception and design. N.R. led the literature search. M.J.J., L.K.S., J.G., M.M., R.S., M.L., D.X., S.R., Y.L., E.N., and V.D. contributed to screening, study selection, data extraction, and risk of bias assessment. N.R. and A.J. conducted the analysis. D.M. contributed clinical expertise and guidance to the manuscript. N.R. and G.G. conducted the GRADE assessments and categorization of interventions. N.R. drafted the initial manuscript. All authors contributed to the interpretation of findings, critically revised the manuscript for important intellectual content, approved the final version, and agreed to be accountable for all aspects of the work.

## Data availability

Made available upon request. For request

## References

1. World Health Organization. HIV statistics, globally and by WHO region, 2025:. information sheet. World Health Organization; 2025.

2. Kanters S, Park JJ, Chan K, Socias ME, Ford N, Forrest JI, et al. Interventions to improve adherence to antiretroviral therapy: a systematic review and network meta-analysis. Lancet HIV. 2017;4(1):e31–e40.

3. Manalel JA, Kaufman JE, Wu Y, Fusaris E, Correa A, Ernst J, et al. Association of ART regimen and adherence to viral suppression: an observational study of a clinical population of people with HIV. AIDS research and therapy. 2024;21(1):68.

4. Bezabhe WM, Chalmers L, Bereznicki LR, Peterson GM. Adherence to Antiretroviral Therapy and Virologic Failure: A Meta-Analysis. Medicine. 2016;95(15):e3361.

5. Milward de Azevedo Meiners MM, Araújo Cruz I, De Toledo MI. Adherence to antiretroviral therapy and viral suppression: Analysis of three periods between 2011 and 2017 at an HIV-AIDS center, Brazil. Frontiers in Pharmacology. 2023;14:1122018.

6. Department for HIV T, Hepatitis and Sexually Transmitted Infections. Global health sector strategies Geneva: World Health Organization; 2022 [Available from: https://www.who.int/teams/global-hiv-hepatitis-and-stis-programmes/strategies/global-health-sector-strategies.

7. United Nations. The Sustainable Development Goals Report. https://www.un.org/en/desa/sustainable-development-goals-report-2016. 2016.

8. World Health Organization. Retention in HIV programmes: defining the challenges and identifying solutions. Meeting report. 2012.

9. Spach DH, Budak JZ, Kalapila AG. Retention in HIV Care 2025 [updated February 3rd, 2025. Available from: https://www.hiv.uw.edu/go/basic-primary-care/retention-care/core-concept/all.

10. Mgbako O, Conard R, Mellins CA, Dacus JD, Remien RH. A systematic review of factors critical for HIV health literacy, ART adherence and retention in care in the U.S. for racial and ethnic minorities. AIDS Behav. 2022;26(11):3480–93.

11. Moges N, Olubukola A, Micheal O, Berhane Y. HIV patients retention and attrition in care and their determinants in Ethiopia: a systematic review and meta-analysis. BMC Infect Dis. 2020;20(1):439.

12. Zheng A, Kileel EM, Brennan AT, Flynn DB, Rosen S, Fox MP. Systematic review and meta-analysis of retention and disengagement after initiation on antiretroviral therapy in low– and middle-income countries after the introduction of universal test and treat policies. J Int AIDS Soc. 2025;28(9):e70026.

13. Lyons SJ, Johnson AS, Hu X, Hou P, Helton B, Elenwa F, et al. Monitoring selected national HIV prevention and care objectives by using HIV surveillance data: United States and 6 dependent areas, 2020. 2022.

14. Center for Disease Control (CDC). National HIV prevention and care outcomes. 2025.

15. UNAIDS. Understanding measures of progress towards the 95–95–95 HIV testing, treatment and viral suppression targets. UNAIDS Geneva, Switzerland; 2023.

16. World Health Organization. Supporting re-engagement in HIV treatment services: policy brief. World Health Organization; 2024.

17. World Health Organization. Consolidated guidelines on the use of antiretroviral drugs for treating and preventing HIV infection: recommendations for a public health approach. World Health Organization; 2016. Report No.: 9241549688.

18. Higa DH, Marks G, Crepaz N, Liau A, Lyles CM. Interventions to improve retention in HIV primary care: a systematic review of U.S. studies. Curr HIV/AIDS Rep. 2012;9(4):313–25.

19. Casale M, Carlqvist A, Cluver L. Recent Interventions to Improve Retention in HIV Care and Adherence to Antiretroviral Treatment Among Adolescents and Youth: A Systematic Review. AIDS Patient Care STDS. 2019;33(6):237–52.

20. Penn AW, Azman H, Horvath H, Taylor KD, Hickey MD, Rajan J, et al. Supportive interventions to improve retention on ART in people with HIV in low-and middle-income countries: A systematic review. PLoS one. 2018;13(12):e0208814.

21. Brignardello-Petersen R, Guyatt GH. Introduction to network meta-analysis: understanding what it is, how it is done, and how it can be used for decision-making. American journal of epidemiology. 2025;194(3):837–43.

22. Salanti G, Higgins JP, Ades A, Ioannidis JP. Evaluation of networks of randomized trials. Statistical methods in medical research. 2008;17(3):279–301.

23. Garcia C, Rehman N, Matos-Silva J, Deng J, Ghandour S, Huang Z, et al. Interventions to Improve Adherence to Oral Pre-exposure Prophylaxis: A Systematic Review and Network Meta-analysis. AIDS and Behavior. 2024:1–13.

24. Brignardello-Petersen R, Bonner A, Alexander PE, Siemieniuk RA, Furukawa TA, Rochwerg B, et al. Advances in the GRADE approach to rate the certainty in estimates from a network meta-analysis. J Clin Epidemiol. 2018;93:36–44.

25. Phillips MR, Sadeghirad B, Busse JW, Brignardello-Petersen R, Cuello-Garcia CA, Nampo FK, et al. Development and design validation of a novel network meta-analysis presentation tool for multiple outcomes: a qualitative descriptive study. BMJ open. 2022;12(6):e056400.

26. Higgins JP, Chandler J, Cumpston M, Li T, Page M, Welch V. Cochrane handbook for systematic reviews of interventions version 6.4 (updated August 2023). cochrane, 20232024.

27. Hutton B, Salanti G, Caldwell DM, Chaimani A, Schmid CH, Cameron C, et al. The PRISMA extension statement for reporting of systematic reviews incorporating network meta-analyses of health care interventions: checklist and explanations. Annals of internal medicine. 2015;162(11):777–84.

28. Brignardello-Petersen R, Murad MH, Walter SD, McLeod S, Carrasco-Labra A, Rochwerg B, et al. GRADE approach to rate the certainty from a network meta-analysis: avoiding spurious judgments of imprecision in sparse networks. J Clin Epidemiol. 2019;105:60–7.

29. Mbuagbaw L, Hajizadeh A, Wang A, et al. Overview of systematic reviews on strategies to improve treatment initiation, adherence to antiretroviral therapy and retention in care for people living with HIV: part 1. BMJ Open. 2020;10(9):e034793.

30. World Bank. World Bank income groups. 2023.

31. World Health Organization. Classifications and standards – Country groupings [Available from: https://www.who.int/observatories/global-observatory-on-health-research-and-development/classifications-and-standards/country-groupings

32. Rehman N, Wu M, Garcia C, Leenus A, El-Kechen H, Bhandari M, et al. Measures of retention in HIV care: a study within a review. AIDS Patient Care STDs. 2023;37(4):192–8.

33. Doyle T, Smith C, Vitiello P, Cambiano V, Johnson M, Owen A, et al. The importance of HIV RNA detection below 50 copies per mL in HIV-positive patients on antiretroviral therapy: an observational study. The Lancet. 2014;383:S44.

34. Wang Y KS, Briel M, Glasziou P, Brignardello-Petersen R,. Development of the Risk of Bias Instrument for Use in Systematic Reviews – for Randomized Controlled Trials (ROBUST-RCT). Under review by BMJ. 2025.

35. Page MJ, Higgins JP, Clayton G, Sterne JA, Hróbjartsson A, Savović J. Empirical evidence of study design biases in randomized trials: systematic review of meta-epidemiological studies. PloS one. 2016;11(7):e0159267.

36. Saltaji H, Armijo-Olivo S, Cummings GG, Amin M, da Costa BR, Flores-Mir C. Influence of blinding on treatment effect size estimate in randomized controlled trials of oral health interventions. BMC medical research methodology. 2018;18(1):42.

37. Higgins JP, Eldridge S, Li T. Including variants on randomized trials. Cochrane handbook for systematic reviews of interventions 2024. p. 569–93.

38. Rücker G. Network meta-analysis, electrical networks and graph theory. Research synthesis methods. 2012;3(4):312–24.

39. Chaimani A, Caldwell DM, Li T, Higgins JP, G S. Chapter 11: Undertaking network meta-analyses. In: al. HJTJCJCMLTPMe, editor. Cochrane Handbook for Systematic Reviews of Interventions. Version 6.5 (updated August 2024) ed. London: Cochrane; 2024.

40. Izcovich A, Chu DK, Mustafa RA, Guyatt G, Brignardello-Petersen R. A guide and pragmatic considerations for applying GRADE to network meta-analysis. BMJ. 2023;381:e074495.

41. Rücker G, Schwarzer G. Reduce dimension or reduce weights? Comparing two approaches to multi-arm studies in network meta-analysis. Statistics in medicine. 2014;33(25):4353–69.

42. Belhadi D, Pacou M, Gauthier A, Taieb V, Mesana L. Checklist to assess the feasibility of a network meta-analysis. Value in Health. 2016;19(3):A100.

43. Cope S, Zhang J, Saletan S, Smiechowski B, Jansen JP, Schmid P. A process for assessing the feasibility of a network meta-analysis: a case study of everolimus in combination with hormonal therapy versus chemotherapy for advanced breast cancer. BMC medicine. 2014;12:1–17.

44. Chaimani A, Caldwell DM, Li T, Higgins JP, Salanti G. Chapter 11: Undertaking network meta-analyses. Cochrane Handbook for Systematic Reviews of Interventions, version 65. London 2024. p. n/a (online).

45. Liu XS. Sample Size and the Precision of the Confidence Interval in Meta-analyses. Ther Innov Regul Sci. 2015;49(4):593–8.

46. Furukawa TA, Barbui C, Cipriani A, Brambilla P, Watanabe N. Imputing missing standard deviations in meta-analyses can provide accurate results. J Clin Epidemiol. 2006;59(1):7–10.

47. Higgins JP, Li T, Deeks JJ. Choosing effect measures and computing estimates of effect. Cochrane handbook for systematic reviews of interventions 2024. p. 143–76.

48. Thorlund K, Walter SD, Johnston BC, Furukawa TA, Guyatt GH. Pooling health-related quality of life outcomes in meta-analysis-a tutorial and review of methods for enhancing interpretability. Res Synth Methods. 2011;2(3):188–203.

49. Ward MM, Guthrie LC, Alba MI. Clinically important changes in short form 36 health survey scales for use in rheumatoid arthritis clinical trials: the impact of low responsiveness. Arthritis care & research. 2014;66(12):1783–9.

50. Schandelmaier S, Briel M, Varadhan R, Schmid CH, Devasenapathy N, Hayward RA, et al. Development of the Instrument to assess the Credibility of Effect Modification Analyses (ICEMAN) in randomized controlled trials and meta-analyses. Cmaj. 2020;192(32):E901–E6.

51. World Health Organization. HIV: fact sheet on Sustainable Development Goals (SDGs): health targets. 2017.

52. Zhou C, Zhang W, Lu R-R, Ouyang L, Xing H, Shao Y-M, et al. Baseline Viral Load Predicts Antiretroviral Therapy Outcomes Among HIV-Infected Patients: An Observational Cohort Study. 2021.

53. Phillips AN, Staszewski S, Weber R, Kirk O, Francioli P, Miller V, et al. HIV Viral Load Response to Antiretroviral Therapy According to the Baseline CD4 Cell Count and Viral Load. JAMA. 2001;286(20):2560–7.

54. Rehman N, Guyatt G, Sabin L, Xiong J, English M, Rae G, et al. Aligning Definitions with Realities: An Interpretive Descriptive Study on the Complexities of Measuring Retention in HIV Care in the Global Context. medRxiv [Internet]. 2026 2026-02-17. Available from: 10.64898/2026.02.13.26345822.

55. Rau H, Nicolai S, Stoll-Kleemann S. A systematic review to assess the evidence-based effectiveness, content, and success factors of behavior change interventions for enhancing pro-environmental behavior in individuals. Frontiers in Psychology. 2022;13:901927.

56. Wieland LS, Berman BM, Altman DG, Barth J, Bouter LM, D’Adamo CR, et al. Rating of included trials on the efficacy–effectiveness spectrum: development of a new tool for systematic reviews. Journal of clinical epidemiology. 2017;84:95–104.

57. Glasgow RE, Bull SS, Gillette C, Klesges LM, Dzewaltowski DA. Behavior change intervention research in healthcare settings: a review of recent reports with emphasis on external validity. American journal of preventive medicine. 2002;23(1):62–9.

58. Hemming K, Taljaard M. Key considerations for designing, conducting and analysing a cluster randomized trial. International journal of epidemiology. 2023;52(5):1648–58.

59. Guyatt G, Zeng L, Brignardello-Petersen R, Prasad M, De Beer H, Murad MH, et al. Core GRADE 2: choosing the target of certainty rating and assessing imprecision. BMJ. 2025;389:e081904.

60. Guyatt G, Agoritsas T, Brignardello-Petersen R, Mustafa RA, Rylance J, Foroutan F, et al. Core GRADE 1: overview of the Core GRADE approach. Bmj. 2025;389.

61. Brignardello-Petersen R, Tomlinson G, Florez I, Rind DM, Chu D, Morgan R, et al. Grading of recommendations assessment, development, and evaluation concept article 5: addressing intransitivity in a network meta-analysis. Journal of Clinical Epidemiology. 2023;160:151–9.

62. Mbuagbaw L, Thabane L, Ongolo-Zogo P, Lester RT, Mills EJ, Smieja M, et al. The Cameroon Mobile Phone SMS (CAMPS) trial: a randomized trial of text messaging versus usual care for adherence to antiretroviral therapy. PLoS One. 2012;7(12):e46909.

63. Lucas GM, Chaudhry A, Hsu J, Woodson T, Lau B, Olsen Y, et al. Clinic-based treatment of opioid-dependent HIV-infected patients versus referral to an opioid treatment program: a randomized trial. Annals of internal medicine. 2010;152(11):704–11.

64. Willis N, Milanzi A, Mawodzeke M, Dziwa C, Armstrong A, Yekeye I, et al. Effectiveness of community adolescent treatment supporters (CATS) interventions in improving linkage and retention in care, adherence to ART and psychosocial well-being: a randomised trial among adolescents living with HIV in rural Zimbabwe. BMC Public Health. 2019;19(1):117.

65. Metsch LR, Feaster DJ, Gooden LK, Masson C, Perlman DC, Jain MK, et al., editors. Care facilitation advances movement along the hepatitis C care continuum for persons with human immunodeficiency virus, hepatitis C, and substance use: a randomized clinical trial (CTN-0064). Open forum infectious diseases; 2021: Oxford University Press US.

66. Inghels M, Kim HY, Mathenjwa T, Shahmanesh M, Seeley J, Wyke S, et al. Population impacts of conditional financial incentives and a male-targeted digital decision support application on the HIV treatment cascade in rural KwaZulu Natal: findings from the HITS cluster randomized clinical trial. Journal of the International AIDS Society. 2024;27(5):e26248.

67. Dias S, Welton NJ, Caldwell DM, Ades AE. Checking consistency in mixed treatment comparison meta-analysis. Statistics in medicine. 2010;29(7-8):932–44.

68. O’Brien N, Chi YL, Krause KR. Measuring Health Outcomes in HIV: Time to Bring in the Patient Experience. Ann Glob Health. 2021;87(1):2.

69. Yehia BR, Stewart L, Momplaisir F, Mody A, Holtzman CW, Jacobs LM, et al. Barriers and facilitators to patient retention in HIV care. BMC Infect Dis. 2015;15:246.

70. World Health Organization. Retention in HIV programmes: defining the challenges and identifying solutions: meeting report, 13–15 September 2011. 2012.

71. Bleasdale J, Leone LA, Morse GD, Liu Y, Taylor S, Przybyla SM. Socio-Structural Factors and HIV Care Engagement among People Living with HIV during the COVID-19 Pandemic: A Qualitative Study in the United States. Trop Med Infect Dis. 2022;7(10.).

72. Anderson AN, Higgins CM, Haardorfer R, Holstad MM, Nguyen MLT, Waldrop-Valverde D. Disparities in Retention in Care Among Adults Living with HIV/AIDS: A Systematic Review. AIDS Behav. 2020;24(4):985–97.

73. Murray KR, Dulli LS, Ridgeway K, Dal Santo L, Darrow de Mora D, Olsen P, et al. Improving retention in HIV care among adolescents and adults in low– and middle-income countries: A systematic review of the literature. PLOS ONE. 2017;12(9):e0184879.

74. Risher KA, Kapoor S, Daramola AM, Paz-Bailey G, Skarbinski J, Doyle K, et al. Challenges in the Evaluation of Interventions to Improve Engagement Along the HIV Care Continuum in the United States: A Systematic Review. AIDS Behav. 2017;21(7):2101–23.

75. Mugavero MJ, Westfall AO, Zinski A, Davila J, Drainoni ML, Gardner LI, et al. Measuring retention in HIV care: the elusive gold standard. J Acquir Immune Defic Syndr. 2012;61(5):574–80.

76. Mugglin C, Kläger D, Gueler A, Vanobberghen F, Rice B, Egger M. The HIV care cascade in sub-Saharan Africa: systematic review of published criteria and definitions. J Int AIDS Soc. 2021;24(7):e25761.

77. Mugglin C, Wandeler G, Estill J, Egger M, Bender N, Davies MA, et al. Retention in care of HIV-infected children from HIV test to start of antiretroviral therapy: systematic review. PLoS One. 2013;8(2):e56446.

78. McClarty LM, Kasper K, Ireland L, Loeppky C, Blanchard JF, Becker ML. The HIV care cascade in Manitoba, Canada: Methods, measures, and estimates to meet local needs. Journal of Clinical Epidemiology. 2021;132:26-33.

79. Jhuti D, Zakaryan G, El-Kechen H, Rehman N, Youssef M, Garcia C, et al. Describing Engagement in the HIV Care Cascade: A Methodological Study. HIV AIDS (Auckl). 2023;15:257–65.

80. Lepere P, Babington-Ashaye A, Martínez-Pérez GZ, Ekouevi DK, Labrique AB, Calmy A. How mHealth Can Contribute to Improving the Continuum of Care: A Scoping Review Approach to the Case of Human Immunodeficiency Virus in Sub-Saharan Africa. Public Health Reviews. 2022;Volume 43 – 2022.

81. Abebe Moges N, Olubukola A, Micheal O, Berhane Y. HIV patients retention and attrition in care and their determinants in Ethiopia: a systematic review and meta-analysis. BMC Infect Dis. 2020;20(1):439.

82. Johnson MO, Neilands TB, Koester KA, Wood T, Sauceda JA, Dilworth SE, et al. Detecting Disengagement From HIV Care Before It Is Too Late: Development and Preliminary Validation of a Novel Index of Engagement in HIV Care. J Acquir Immune Defic Syndr. 2019;81(2):145–52.

83. Hughes J, Jelsma J, Maclean E, Darder M, Tinise X. The health-related quality of life of people living with HIV/AIDS. Disability and rehabilitation. 2004;26(6):371–6.

84. Kim MT, Han H-R. Instrumentation development. In: M F-S, SJ O, editors. Instruments for clinical health-care research: Jones & Barlett; 2004. p. 73–82.

