## Supplementary file 2. Appendixes and tables for "Interventions to improve retention in HIV care: a systematic review and network meta-analysis of randomised controlled trials"

**Supplementary file 1**

**Interventions to improve retention in HIV care: A systematic review and network meta-analysis of randomised controlled trials**

[Database: Embase < 1974 to 2020 December 18 > via Ovid	5](#_Toc222597595)

[Database: PsycINFO < 1987 to December 2020 > via Ovid	6](#_Toc222597596)

[Table S25. Follow-up time (< 12 vs 12 mos): Subgroup analysis of retention in HIV care	42](#_Toc222597630)

[Table S33. Follow-up time (< 12 vs 12 mos): Subgroup analysis for viral load suppression	53](#_Toc222597639)

### Abbreviations

#### Intervention categories

BC = behavioural counselling

CM = case management

DSD = differentiated service delivery

EDU = education

EI = economic incentives

eSoC = enhanced standard of care

mHealth = mobile health interventions

MM = medication modification

MMD = multi‑month dispensing

REM = reminder systems

SoC = standard of care

SUP = peer support interventions

TS = task shifting

#### World Bank country income classification

HIC = high‑income country

UMIC = upper‑middle‑income country

LMIC = lower‑middle‑income country

LIC = low‑income country

#### WHO geographical regions

AFR = African Region

AMR = Region of the Americas

EMR = Eastern Mediterranean Region

EUR = European Region

SEAR = South‑East Asia Region

WPR = Western Pacific Region

Population groups (WHO key population definitions)
ACB = African, Caribbean, or Black
MSM = men who have sex with men
PWID = people who inject drugs

Risk‑of‑bias and assessment tools
ROBUST‑RCT = Risk Of Bias tool for randomised trials
ICEMAN = Instrument for Credibility of Effect Modification Analyses
RITES = Rating of Included Trials on the Efficacy–Effectiveness Spectrum

Quality‑of‑life instruments
SF‑12, SF‑36 = Short Form Health Surveys
MOS‑HIV = Medical Outcomes Study HIV Health Survey
HRQoL = Health‑related quality of life
PACIC = Patient Assessment of Chronic Illness Care

#### Meta‑analytic notation

k = number of studies

m = number of effect estimates

n = sample size per arm

The credibility of any apparent subgroup effect

(regression coefficient’s credible interval excludes

null effect) was rated using the ICEMAN tool.26 If no

credible subgroup effect was indicated, we assumed

the constancy of relative effects across populations

### Appendix S1. Search strategy

#### Database: CINAHL via EBSCOhost Research Databases

1. (MH "Randomised controlled trials")
2. (MH "Patient Compliance+") OR (MH "Treatment Refusal") OR (MH "Medication Compliance")
3. (MH "Research Subject Retention")
4. (MH "Research Dropouts") OR (MH "Patient Dropouts")
5. (MH "After Care")
6. adhere* OR nonadhere* OR complian* OR uncomplian* OR retention OR dropout OR lost to follow-up OR attrition OR treatment refusal OR persistence OR non-persistence OR initiat* OR start* OR uptake
7. S2 OR S3 OR S4 OR S5 OR S6
8. (MH "Human Immunodeficiency Virus+")
9. (MH "HIV-Infected Patients+") OR (MH "HIV Infections+")
10. HIV OR human immun* deficiency virus
11. S8 OR S9 OR S10
12. (MH "Antiretroviral Therapy, Highly Active")
13. antiretroviral therapy OR antiretrovirals OR antiretroviral treatment OR anti-HIV agents

OR anti-retroviral agents OR Highly Active Antiretroviral Therapy OR HAART

14. S12 OR S13
15. S1 AND S7 AND S11 AND S14

#### Database: Cochrane Central Register of Controlled Trials (CENTRAL)

1. adhere* OR nonadhere* OR complian* OR uncomplian* OR retention OR dropout OR lost to follow-up OR attrition OR treatment refusal OR persistence OR non-persistence OR initiat* OR start* OR uptake in All Text AND HIV OR human immun* adj2 deficiency virus in All Text AND antiretroviral therapy OR antiretrovirals OR antiretroviral treatment OR anti-HIV agents OR anti-retroviral agents OR Highly Active Antiretroviral Therapy OR ART OR HAART in All Text

#### Database: Embase < 1974 to 2020 December 18 > via Ovid

1. randomised controlled trial.mp. or "exp randomised controlled trial"/
2. (complian* or uncomplian*).mp. or exp "medication compliance"/
3. retention.mp.
4. dropout.mp. or exp "patient dropout"/
5. (los* adj2 to follow up).mp. [mp=title, abstract, heading word, drug trade name, original title, device manufacturer, drug manufacturer, device trade name, keyword, floating subheading word, candidate term word]
6. attrition.mp.
7. (adhere* or nonadhere*).mp. [mp=title, abstract, heading word, drug trade name, original title, device manufacturer, drug manufacturer, device trade name, keyword, floating subheading word, candidate term word]
8. treatment refus*.mp. or exp "treatment refusal"/
9. persistence.mp.
10. initiat*.mp.
11. start*.mp
12. uptake.mp.
13. 2 or 3 or 4 or 5 or 6 or 7 or 8 or 9 or 10 or 11 or 12
14. (HIV or human immune-deficiency virus or human immuno-deficiency virus).mp.
15. exp "human immunodeficiency virus"/ or exp "human immunodeficiency virus infection"/ or exp "human immunodeficiency virus infected patient"/
16. 14 or 15
17. antiretroviral therapy.mp. or exp "antiretroviral therapy"/
18. antiretrovirals.mp.
19. antiretroviral treatment.mp.
20. exp "antiretrovirus agent"/
21. highly active antiretroviral therapy.mp. or exp "highly active antiretroviral therapy"/
22. (ART or HAART).mp.
23. 17 or 18 or 19 or 20 or 21 or 22
24. 1 and 13 and 16 and 23
25. limit year

#### Database: PsycINFO < 1987 to December 2020 > via Ovid

1. randomised controlled trials.mp. or exp randomised controlled trials/
2. exp "compliance"/ or exp "treatment compliance"/
3. (complian* or uncomplian*).mp.
4. dropout.mp. or exp "treatment dropout"/
5. retention.mp.
6. attrition.mp. or exp "experimental attrition"/
7. (los* adj2 to follow up).mp. [mp=title, abstract, heading word, table of contents, key concepts, original title, tests & measures]
8. adhere*.mp.
9. treatment refus*.mp. or exp "treatment refusal"/
10. persistence.mp.
11. initiate.mp.
12. start*.mp.
13. uptake.mp.
14. nonadherence.mp.
15. 2 or 3 or 4 or 5 or 6 or 7 or 8 or 9 or 10 or 11 or 12 or 13 or 14
16. HIV.mp. or exp "HIV"/
17. (human immunodeficiency virus or human immune-deficiency virus or human immuno-deficiency virus).mp.
18. 16 or 17
19. exp "drug therapy"/
20. (antiretroviral or antiretroviral therapy or antiretroviral treatment or ART or HAART).mp.
21. 19 or 20
22. 1 and 15 and 18 and
23. limit 22 to yr="1995 -Current"

#### Database: PubMed

1. (randomised controlled trial) AND (adhere* OR nonadhere* OR complian* OR uncomplian* OR retention OR dropout OR lost to follow-up OR attrition OR treatment refusal OR persistence OR non-persistence OR initiat* OR start* OR uptake) AND (HIV OR human immune-deficiency virus OR human immuno-deficiency virus) AND (antiretroviral therapy OR antiretrovirals OR antiretroviral treatment OR Highly Active Antiretroviral Therapy OR ART OR HAART OR anti-HIV agents OR anti-retroviral agents) Filters: Publication date from 2018/12/31 to onwards

MeSH headings [mh], captured via ‘All Fields’ search:

1. patient dropouts / patient compliance (patient adherence/nonadherence) / treatment adherence and compliance (therapeutic adherence/compliance) / medication adherence (nonadherence, compliance/noncompliance) /lost to follow-up
2. HIV / anti-HIV agents / anti-retroviral agents / antiretroviral therapy, highly active (HAART)
3. randomised controlled trial	"randomised controlled trial"[Publication Type] OR "randomised controlled trials as topic"[MeSH Terms] OR "randomised controlled trial"[All Fields] OR "randomised controlled trial"[All Fields]	And
4. retention	"retention (psychology)"[MeSH Terms] OR ("retention"[All Fields] AND "(psychology)"[All Fields]) OR "retention (psychology)"[All Fields] OR "retention"[All Fields]	OR
5. lost to follow-up	"lost to follow-up"[MeSH Terms] OR ("lost"[All Fields] AND "follow-up"[All Fields]) OR "lost to follow-up"[All Fields] OR ("lost"[All Fields] AND "follow"[All Fields] AND "up"[All Fields]) OR "lost to follow up"[All Fields]
6. attrition	"tooth attrition"[MeSH Terms] OR ("tooth"[All Fields] AND "attrition"[All Fields]) OR "tooth attrition"[All Fields] OR "attrition"[All Fields] OR
7. treatment refusal	"treatment refusal"[MeSH Terms] OR ("treatment"[All Fields] AND "refusal"[All Fields]) OR "treatment refusal"[All Fields]	AND
8. HIV	"hiv"[MeSH Terms] OR "hiv"[All Fields]
9. human	"humans"[MeSH Terms] OR "humans"[All Fields] OR "human"[All Fields]	OR
10. immune-deficiency	"immunologic deficiency syndromes"[MeSH Terms] OR ("immunologic"[All Fields] AND "deficiency"[All Fields] AND "syndromes"[All Fields]) OR "immunologic deficiency syndromes"[All Fields] OR ("immune"[All Fields] AND "deficiency"[All Fields]) OR "immune deficiency"[All Fields]
11. virus	"viruses"[MeSH Terms] OR "viruses"[All Fields] OR "virus"[All Fields]
12. immuno-deficiency	"immunologic deficiency syndromes"[MeSH Terms] OR ("immunologic"[All Fields] AND "deficiency"[All Fields] AND "syndromes"[All Fields]) OR "immunologic deficiency syndromes"[All Fields] OR ("immuno"[All Fields] AND "deficiency"[All Fields]) OR "immuno deficiency"[All Fields]	AND
13. therapy	"therapy"[Subheading] OR "therapy"[All Fields] OR "therapeutics"[MeSH Terms] OR "therapeutics"[All Fields]
14. treatment	"therapy"[Subheading] OR "therapy"[All Fields] OR "treatment"[All Fields] OR "therapeutics"[MeSH Terms] OR "therapeutics"[All Fields]
15. Highly Active Antiretroviral Therapy	"antiretroviral therapy, highly active"[MeSH Terms] OR ("antiretroviral"[All Fields] AND "therapy"[All Fields] AND "highly"[All Fields] AND "active"[All Fields]) OR "highly active antiretroviral therapy"[All Fields] OR ("highly"[All Fields] AND "active"[All Fields] AND "antiretroviral"[All Fields] AND "therapy"[All Fields])
16. ART	"art"[MeSH Terms] OR "art"[All Fields]
17. HAART	"antiretroviral therapy, highly active"[MeSH Terms] OR ("antiretroviral"[All Fields] AND "therapy"[All Fields] AND "highly"[All Fields] AND "active"[All Fields]) OR "highly active antiretroviral therapy"[All Fields] OR "haart"[All Fields]
18. anti-HIV agents	"anti-hiv agents"[Pharmacological Action] OR "anti-hiv agents"[MeSH Terms] OR ("anti-hiv"[All Fields] AND "agents"[All Fields]) OR "anti-hiv agents"[All Fields] OR ("anti"[All Fields] AND "hiv"[All Fields] AND "agents"[All Fields]) OR "anti hiv agents"[All Fields]
19. anti-retroviral agents	"anti-retroviral agents"[Pharmacological Action] OR "anti-retroviral agents"[MeSH Terms] OR ("anti-retroviral"[All Fields] AND "agents"[All Fields]) OR "anti-retroviral agents"[All Fields] OR ("anti"[All Fields] AND "retroviral"[All Fields] AND "agents"[All Fields]) OR "anti retroviral agents"[All Fields]

#### Database: Web of Science

1. (randomised controlled trials)
2. adhere* OR nonadhere* OR complian* OR uncomplian* OR retention OR dropout OR los* follow-up OR attrition OR treatment refusal OR persistence OR non-persistence OR initiat* OR start* OR uptake)
3. HIV OR human immun* deficiency virus)
4. antiretroviral therapy OR antiretrovirals OR antiretroviral treatment OR anti-HIV agents OR anti-retroviral agents OR ART OR Highly Active Antiretroviral Therapy OR HAART)
5. #5	#4 AND #3 AND #2 AND #1
6. (Indexes=SCI-EXPANDED, SSCI, A&HCI, CPCI-S, CPCI-SSH, ESCI Timespan= 2018-current)

### Definitions, intervention descriptions, and study characteristics

#### Table S1. Trials eligible for retention outcomes but excluded from the network meta-analysis

| **Author** | **Study design** | **Reasons for exclusion** |
| --- | --- | --- |
| Fayorsey et al., 2019^1^ | Parallel RCT | Comparator not jointly randomizable (usual pregnancy care) |
| Washington., 2015^2^ | Cluster RCT | Intervention is not jointly randomizable (HIV care in antenatal clinics) |
| Maskew et al., 2020^3^ | Parallel RCT | Intervention not jointly randomizable due to guideline changes (early ART initiation) |
| Khan et al., 2020^4^ | Parallel RCT | Intervention not jointly randomizable due to guideline changes (early ART initiation) |
| Jani et al., 2018^5^ | Cluster RCT | Intervention not jointly randomizable (point-of-care early infant testing) |
| Sarna et al., 2019^6^ | Parallel RCT | Comparator not jointly randomizable (usual pregnancy care) |
| Audet et al., 2021^7^ | Parallel RCT | Intervention not jointly randomizable (sero-discordant family intervention) |
| Odeny et al., 2019^8^ | Cluster RCT | Intervention is not jointly randomizable, and the outcome is mother-to-child transmission retention and infant testing (usual pregnancy care) |
| Rotheram-Borus et al., 2012 ^9^ | Cluster RCT | Population and intervention not jointly randomizable (family intervention) |
| Dorvil et al., 2023 ^10^ | Parallel RCT | Treatment and population not jointly randomizable (Early ART or tuberculosis treatment) |
| Chandra et al., 2019 ^11^ | Parallel RCT | Treatment and population not jointly randomizable (opioid dependence in men with HIV) |
| Rosen et al., 2019 ^12^ | Parallel RCT | Intervention not jointly randomizable guideline changes (early ART initiation) |

Studies were excluded from the network meta-analysis when interventions, comparators, populations, or outcomes were not jointly randomizable or comparable within the network

#### Table S2. Retention definitions, viral load thresholds, and quality‑of‑life instruments

| **Author (Year)** | **Retention definition** | **Viral load threshold copies/mL** | **Quality of life instrument** |
| --- | --- | --- | --- |
| Mbuagbaw 2012^13^ | Number retained in care | NR | SF-12^1^ |
| Lucas 2010^14^ | Visits with primary HIV care providers | NR | NR |
| Keitz 2001^15^ | Scheduled and unscheduled visits | NR | SF-12 |
| Odeny 2014^16^ | Retention in PMTCT: Maternal Clinic attendance | NR | NR |
| Chang 2010^17^ | Patients were considered lost to follow-up if they had not had a pharmacy visit for medication pick-up in over 90 days | < 400 | NR |
| Wohl 2011^18^ | Access to medical care: Self-reported access to post-release medical care | NR | NR |
| Naar-King 2009^19^ | Retention in care was measured using medical chart review data. A primary care visit was defined as care provided by a physician, nurse practitioner, or physician assistant who could monitor CD4 and viral load counts and prescribe HIV medications. Medical visits to specialists for complications of HIV, emergency visits, or in-patient hospitalizations were not considered HIV primary care visits. | NR | NR |
| Gwadz 2015^20^ | Appointment attendance | < 50 | NR |
| Norton 2014^21^ | Clinic attendance: attendance at clinic appointments | NR | NR |
| MacGowan 2014^22^ | Health care at HIV clinic | NR | NR |
| Gardner 2014^23^ | Number of kept visits in 12 months divided by the total number of scheduled appointments, excluding cancellations | NR | NR |
| Konkle-Parker 2014^24^ | Visit constancy (at least one kept visit per time segment) (at least one kept HIV medical visit in each third of the year following baseline assessment) | NR | NR |
| Huang 2013^25^ | Visit | NR | NR |
| Wamalwa 2009^26^ | Clinic visits | < 100 | NR |
| Wohl 2006^27^ | Completed six-month follow-up period | < 400 | NR |
| Chander 2015^28^ | Appointment adherence: the number of completed visits over total number of scheduled visits. scheduled visits, completed visits, missed visits | < 50 | NR |
| Kunutsor 2011^29^ | Clinic attendance for each client was monitored every 4 weeks based on the given clinic appointment dates for ARV refills (these were usually scheduled at 4-weekly intervals). Clinic attendance was classified as attendance on or before appointment day, within 3 days of appointment day, after 3 days of appointment day and a missed visit. | NR | NR |
| Dulli 2020^30^ | Retained in HIV care, defined as not having missed a scheduled appointment by more than 28 days | NR | NR |
| Ammassari 2018^31^ | Number of visits with +95% level adherence | NR | NR |
| Grave 2018^32^ | Retained in care if they attended any ART clinic appointment at least once over the last 3 months of the study period | NR | NR |
| El-Sadr 2019^33^ | Proportion having evidence of a clinical visit (ie, a CD4+ cell count or viral load test data in the Surveillance Database) in 4 of the prior 5 quarters | NR | NR |
| Fahey 2020^34^ | Following PEPFAR guidelines, individuals considered not retained in ART care include those who died, disengaged from care or otherwise stopped ART, or had no evidence of facility-based care for 28 days or more after a missed appointment | < 1000 | NR |
| Myer 2018^35^ |  | < 1000 | NR |
| McLaughlin 2018^36^ | Retention with HIV viral load suppression (<400 copies/mL) | NR | NR |
| Kadota 2018^37^ | Appointment attendance: assessed by the number of participants in each arm. The proportion of participants retained in care was defined as one minus the probability of LTFU (≥3 months since the last scheduled visit). | NR | NR |
| Kalichman 2018^38^ | Come for the office assessment | NR | NR |
| Neduzhko 2020^39^ | Retention, defined as at least one additional HIV clinical visit within 6 months after linkage to HIV care (yes/no) | NR | NR |
| Kim 2019^40^ | Short-term retention (retention at 1 month) in the ART clinic was defined as retained if a visit occurred between 14 and 61 days after the ART start date. | NR | NR |
| Mavhu 2020^41^ | ART continuation for the last 3 months | NR | NR |
| Horvath 2019^42^ | Retention at the 6 months | NR | NR |
| Samet 2019^43^ | Proportion of participants attending at least one follow-up visit | NR | NR |
| Sherman 2020^44^ | The number of participants in care at the clinic at the time of the follow-up visit was divided by the total number of patients enrolled in the treatment arm. | < 50 | NR |
| Willis 2019^45^ | Retention in care | NR | Study-specific  tool |
| Sabin 2020^46^ | Proportion of participants retained in care (defined as attending ≥80% of Clinic visits) | NR | NR |
| Pascoe 2019^47^ | Not transferred, become lost to follow-up, or died. | < 400 | NR |
| Silverman 2019^48^ | Attended 2 or more visits in the year (missing = not attended) | < 40 | NR |
| Goodrich 2021^49^ | Retained in study, assigned care | < 1000 | NR |
| Bynonanebye 2021^50^ | Appointment keeping at 12 months | NR | MOS-HIV^2^ |
| Hoffman 2021^51^ | The primary outcome was retention in care at 12 months, defined as the proportion of patients with less than 60 consecutive days without ART at any point during follow-up. | NR | NR |
| Cassidy 2020^52^ | The proportion of patients with less than | < 400 | NR |
| Kinuthia 2021^53^ | On-time clinic visit attendance during follow-up to 12 and 24 months postpartum was defined as the proportion of scheduled Clinic visits attended on time and | NR | NR |
| Ayer 2021^54^ | If they had 100% of ARV pick-ups on time (within 2 days of the scheduled date) | NR | NR |
| Cedric H 2021^55^ | Completed the 3-month lab visit | < 400 | NR |
| Graham 2021^56^ | Number of participants retained by the end of the study | NR | NR |
| Ndhlovu 2021^57^ | Proportion of participants retained in HIV care at 12 months | NR | NR |
| Fatti 2020^58^ | 1-participant attrition | < 1000 | NR |
| Wagner 2021^59^ | Attended their most recent scheduled routine care visit or had been seen by their provider in the past 6 months | < 20 | NR |
| Stephenson 2021^60^ | Has attended clinical appointment for HIV care with a viral load test in past 6 months | NR | NR |
| Tukei 2020^61^ | The proportion of participants remaining in care 12 months after study enrollment. | < 1000 | NR |
| Roy 2020^62^ | First late drug pickup (>7 days late) | NR | NR |
| Drain 2021^63^ | Retained in care was defined as collecting ART at the study clinic or a community pick-up point between 44 weeks and 56 weeks after enrolment. Participants not retained in care at 56 weeks were tracked by the study team for up to 60 weeks for viral load testing. | < 200 | NR |
| Giordona 2016^64^ | Appointment data from TSHC were electronically available and imported into study databases. If participants reported using medical facilities outside the Harris Health System during follow- up, those medical records were reviewed and abstracted. | < 400 | HRQoL^3^ |
| Liu 2022^65^ | No of participants retained in care | < 50 | NR |
| Hickey 2021^66^ | Proportion of follow-up time spent adhering to clinic visit schedules over 12 months of follow-up. We calculated gaps in care by determining the number of days between a missed clinic visit and the patient's return to any clinic within Homa Bay County. Participants were censored on the date of death or transfer to a facility outside Homa Bay County. Thus, 90-day disengagement indicates missing an appointment by at least 90 days and not known to have first transferred to another facility or died. Time in care is the proportion of follow up time that a participant adhered to clinic appointments and was calculated as [(total follow up time)–(sum of gaps in care)]/(total | NR | NR |
| Kebabya 2021^67^ | Proportion of children seen at the well-baby clinic at six and ten weeks | NR | NR |
| Hightow-Weidman 2023^68^ | Has at least had 1 HIV related visit in 3 months | < 1000 | NR |
| Lewis 2022^69^ | Participant having at least one visit in each 6-month period within 12 months post-randomisation, with the 2 visits separated by at least 2 months. | < 200 | NR |
| Chang 2023^70^ | Proportion with a 12-month clinical visit) | < 50 | NR |
| Fahey 2022^71^ | Clinic attendance records at 24 months after enrolment. | NR | NR |
| Metsch 2021^72^ | Attendance at HIV primary care visit. Self-report and medical record abstraction | < 200 | NR |
| Amone 2024^73^ | Proportion of participants who were not terminated. Complete adherence to visits was defined as having attended the 4 scheduled quarterly visits at the end of each of the first and second years of postpartum follow-up as defined by the MOH | NR | NR |
| Ayieko 2023^74^ | Proportion of time in care, with “out-of-care” time starting 14 days after a missed visit and ending with reengagement in care | < 400 | NR |
| Naggirinya 2024^75^ | Proportion of participants who turned up for care | < 1000 | NR |
| Derose 2022^76^ | Missed 1+ appointments in the past 6 months | NR | NR |
| Horvath 2024^42^ | Completed visit | < 20 | NR |
| Inghels 2024^77^ | Individuals living with HIV were considered on ART if they were documented as on ART in the Tier.Net datasets or have a recent ART appointment (less than 3 months) in the AHRILink datasets. | Change in HIV VL | NR |
| Luoma 2023^78^ | Self-reported engagement in services in the past 6 months | NR | NR |
| Mabuto 2024^79^ | Number enrolled in HIV treatment services after leaving the correctional health facility | NR | NR |
| Martin 2024^80^ | Number with 2+ HIV primary care visits separated by at least 90 days apart by 24 weeks | < 50 | NR |
| Ndongo 2024^81^ | Compliance with the scheduled medical visit (including drugs’ pick-up count, drugs’ delivery, minimal medical check-up, and routine viral load testing if applicable), i.e. not having more than 3 months of interval from the scheduled medical visit date without having been received in person at the health facility | NR | NR |
| Njau 2024^82^ | Proportion of visits attended on time: Proportion of visits attended within 4 days | < 1000 | NR |
| Njuguna 2024^83^ | ART-related Clinic Visit: Participants returned to clinic within less than or equal to 45 days after first message. | NR | NR |
| Novak 2023^84^ | Attended 2+ visits in year | < 200 | NR |
| Onoya 2024^85^ | Retention in care is defined as being within 28 days  late for the last scheduled appointment in the first 12 months in care | < 50 | NR |
| Palar 2024^86^ | Study retention after 6 months | < 20 | SF-36^4^ |
| Marc 2022^87^ | Attended a visit from  24 to 72 weeks after enrollment | < 50 | NR |
| Parry 2023^88^ | Retention | < 50 | NR |
| Peck 2024^89^ | Retention in care was defined by an active ART prescription at 12 months provided by an HIV clinic visit. | NR | NR |
| Reid 2017^90^ | One visit with doctor | NR | NR |
| Samet 2023^91^ | 1 visit in 2 consecutive 6-month periods | < 40 | NR |
| Solomon 2024^92^ | Proportion retained to HIV care who completed one or more visits to the government HIV clinic in both 0–6 months and 6–12 months | < 200 | NR |
| Zani 2024^93^ | Retention in care: Patients with a clinic visit in the past 3 months | < 1000 | PACIC^5^ |
| Giovenco 2024^94^ | The reason for not receiving a delivery after enrollment was asked. | < 200 | NR |
| Limbada 2021^95^ | Proportion of people retained in their originally allocated group. For this outcome, participants were considered non-retained in the models of care if they transitioned back to SoC for any reason, including comorbidities, lost to follow-up, death, opting out of the intervention, or withdrawal. | NR | NR |

¹ SF‑12: 12 Item Short Form Survey

^2^ MOS‑HIV: Medical Outcomes Study HIV Health Survey
^3^ HRQoL: Health-related quality of life
^4^ SF‑36: 36 Item Short Form Survey
^5^ PACIC: Patient Assessment of Chronic Illness Care

#### Table S3. Intervention descriptions

| Author (Year) | Intervention vs Comparator | Setting | Design | Intensity | Pragmatism | Trial length (mon) | Intervention name | Rationale |
| --- | --- | --- | --- | --- | --- | --- | --- | --- |
| Mbuagbaw 2012^13^ | mHealth vs SoC | Cameroon | Parallel | High | 4 | 7 | Motivational text messages | Motivational SMS supporting ART adherence |
| Lucas 2010^14^ | TS vs CM | USA | Multi-arm | Moderate | 2 | 12 | Clinic-based treatment for OUD | Case management for PLHIV |
| Keitz 2001^15^ | TS vs SoC | USA | Parallel | Moderate | 4 | 12 | Educational program for clinicians | Clinician education strengthening HIV care processes |
| Odeny 2014^16^ | mHealth vs REM | Kenya | Parallel | High | 5 | 11 | Interactive text messages | Interactive SMS promoting clinic attendance and engagement |
| Chang 2010^17^ | SUP vs SoC | Uganda | Cluster | Moderate | 4 | 14 | Peer Health Worker intervention | Peer health worker counselling reinforcing HIV care |
| Wohl 2011^18^ | BC vs DSD | Kenya | Parallel | Moderate | 2 | 9 | Medication diaries + counselling | Medication diaries paired with brief counselling |
| Naar-King 2009^19^ | TS vs BC | USA | Parallel | Low | 3 | 36 | DAART | Directly observed ART enhancing adherence |
| Gwadz 2015^20^ | BC vs eSoC | Uganda | Parallel | Low | 3 | 12 | Heart to Heart (HTH) | Behavioural counselling addressing barriers to ART initiation |
| Norton 2014^21^ | REM vs SoC | European Countries | Parallel | High | 2 | 4 | Call‑em‑all | SMS reminders promoting appointment attendance |
| MacGowan 2014^22^ | BC vs CM | Tanzania | Parallel | Low | 3 | 13 | POST skills‑building (IMB model) | IMB‑based education and skills training |
| Gardner 2014^23^ | EDU+BC vs REM | USA | Multi-arm | Moderate | 3 | 12 | Enhanced personal contact ± skills | Structured contact and skills training supporting retention |
| Konkle-Parker 2014^24^ | BC vs SoC | USA | Parallel | Low | 3 | 24 | IMB model intervention | IMB‑based approach reinforcing retention and adherence |
| Huang 2013^25^ | REM vs EDU | China | Parallel | Low | 2 | 3 | Mobile health | Mobile support increasing engagement in HIV care |
| Wamalwa 2009^26^ | REM vs SoC | USA | Parallel | Continuous | 3 | 17 | Medication diaries + counselling | Caregivers supported diaries and counselling |
| Wohl 2006^27^ | DSD+SUP vs SoC | Nepal | Parallel | Continuous | 3 | 33 | DAART | CHW‑supported DOT with emotional support |
| Chander 2015^28^ | BC vs SoC | USA | Parallel | Low | 3 | 51 | Brief alcohol intervention | Brief alcohol intervention reducing hazardous drinking |
| Kunutsor 2011^29^ | SUP vs SoC | Uganda | Parallel | Low | 3 | 6 | Treatment supporter (TS) | Treatment supporters facilitating ART access and adherence |
| Dulli 2020^30^ | mHealth vs SoC | USA | Parallel | Low | 5 | 15 | SMART Connections | mHealth support for adherence and retention among youth |
| Ammassari 2018^31^ | MM vs SoC | European countries | Parallel | Moderate | 3 | 13 | RAL+DRV/r | Boosted INSTI regimen evaluated for efficacy and safety |
| Grave 2018^32^ | EDU vs SoC | Uganda | Cluster | Low | 5 | 6 | Family Clinic Day | Family‑centred ART support for children and adolescents |
| El-Sadr 2019^33^ | EI vs SoC | USA | Parallel | Moderate | 5 | 23 | Financial incentives (FI) | Financial incentives sustaining viral suppression and continuity |
| Fahey 2020^34^ | EI vs SoC | Tanzania | Parallel | Moderate | 5 | 9 | Financial incentives | Small incentives promoting retention and suppression |
| Myer 2018^35^ | DSD vs SoC | South Africa | Parallel | Low | 3 | 18 | ANC ART integration | Integration of ART into antenatal care |
| McLaughlin 2018^36^ | DSD vs SoC | Peru | Cluster | Moderate | 5 | 52 | DOT‑cART (community-based) | Community‑based DOT with supportive counselling |
| Kadota 2018^37^ | EI vs SoC | Tanzania | Parallel | Moderate | 3 | 19 | Food/cash transfers + NAC | Cash and food transfers with nutrition counselling |
| Kalichman 2018^38^ | mHealth vs SoC | USA | Parallel | Low | 5 | 40 | B‑TasP (enhanced In‑the‑Mix) | Social cognitive mHealth program enhances adherence and risk reduction |
| Neduzhko 2020^39^ | BC vs SoC | Ukraine | Parallel | Low | 3 | 29 | Modified ARTAS (MARTAS) | Linkage counselling addressing depression, stigma, and substance use |
| Kim 2019^40^ | mHealth vs SoC | Malawi | Parallel | Single | 5 | 14 | VITAL Start video | Video counselling facilitating ART initiation |
| Mavhu 2020^41^ | DSD vs SoC | Zimbabwe | Cluster | Moderate | 3 | 7 | Zvandiri (peer-led) | Peer‑led DSD supporting adolescent clinical and psychosocial outcomes |
| Horvath 2019^42^ | mHealth vs SoC | USA | Pilot | Continuous | 3 | 7 | APP+ | Adherence app supporting engagement among stimulant‑using MSM |
| Samet 2019^43^ | BC vs EDU | Russia | Parallel | Low | 3 | 22 | LINC case management | Strengths‑based case management supporting linkage and retention |
| Sherman 2020^44^ | REM vs SoC | USA | Parallel | High | 3 | 31 | SMS reminders | Daily SMS reminders reinforcing adherence |
| Willis 2019^45^ | SUP vs eSoC | Zimbabwe | Parallel | High | 5 | 11 | CATS | Peer adolescent support for linkage, retention, and well-being |
| Sabin 2020^46^ | mHealth vs SoC | Uganda | Parallel | High | 3 | 8 | Wireless pill monitors (WPM) | IMB‑guided wireless pill monitoring with counselling |
| Pascoe 2019^47^ | EDU vs SoC | South Africa | Cluster | Low | 4 | 12 | FTIC | Standardized counselling accelerating ART initiation |
| Silverman 2019^48^ | EDU vs EI | USA | Parallel | Continuous | 3 | 35 | Incentive groups | Financial incentives supporting viral suppression |
| Goodrich 2021^49^ | DSD vs SoC | Kenya | Cluster | Moderate | 5 | 16 | ART Co-ops (community-based) | Lay‑led community ART delivery model |
| Bynonanebye 2021^50^ | mHealth vs SoC | Uganda | Parallel | Single | 4 | 10 | Interactive voice response (IVR) | IMB‑based IVR reinforcing adherence and retention |
| Hoffman 2021^51^ | MMD vs SoC | Zambia; Malawi | Cluster | Low | 5 | 12 | 6-month ART dispensing | Six‑month ART dispensing reduces visit burden |
| Cassidy 2020^52^ | DSD vs DSD+MMD | South Africa | Cluster | Low | 5 | 24 | Six-month refill | Extended refill spacing reduces clinic visits |
| Kinuthia 2021^53^ | mHealth+REM vs SoC | Kenya | Multi-arm | High | 5 | 18 | Two-way SMS | One‑way and two‑way SMS compared for retention outcomes |
| Ayer 2021^54^ | EDU vs REM | Nepal | Parallel | Low | 5 | 8 | Nurse phone call reminder | Nurse phone calls supporting literacy and on‑time attendance |
| Cedric H 2021^55^ | BC vs EI | USA | Parallel | Low | 5 | 3 | Lottery-based incentives (Way to Health) | Regret‑aversion incentives reinforcing adherence |
| Graham 2021^56^ | BC vs SUP | Kenya | Parallel | Low | 3 | 22 | Shikamana | IMB‑based counselling supporting adherence among GBMSM |
| Ndhlovu 2021^57^ | SUP vs SoC | South Africa | Parallel | Moderate | 4 | 19 | Zvandiri | Layered peer psychosocial support |
| Fatti 2020^58^ | DSD & DSD+MMD vs SoC | Zimbabwe | Multi-arm | Low | 3 | 23 | Stronger Together (CHTC + Partner STEPS) | Dyadic counselling and couple testing supporting engagement |
| Wagner 2021^59^ | BC vs SoC | USA | Parallel | Low | 3 | 59 | START (IMB-based) | IMB‑based adherence readiness training |
| Stephenson 2021^60^ | BC vs EDU | USA | Parallel | Low | 3 | 48 | Stronger Together | Dyadic Life STEPS counselling supporting adherence |
| Tukei 2020^61^ | DSD & DSD+MMD vs SoC | Lesotho | Cluster | Low | 2 | 12 | Multi-month dispensing (MMD) | Extended refill intervals for stable patients |
| Roy 2020^62^ | DSD vs eSoC | Zambia | Cluster | Low | 5 | 17 | Adherence clubs | Group pickups with counselling and spaced visits |
| Drain 2021^63^ | SUP+TS vs BC | South Africa | Parallel | Single | 3 | 18 | eSTREAM | Point‑of‑care VL testing with same‑day counselling and task shifting |
| Giordona 2016^64^ | BC vs SUP | USA | Parallel | Low | 3 | 36 | MAPPS (modified) | Peer mentor services for patients not retained in care |
| Liu 2022^65^ | REM vs SoC | USA | Parallel | Continuous | 5 | 29 | Ingestible sensor (IS) | Ingestible sensor system confirming ingestion and triggering prompts |
| Hickey 2021^66^ | SUP vs SoC | Kenya | Cluster | Moderate | 3 | 30 | Microclinic (“kanyaklas”) | Microsocial networks reducing stigma and supporting engagement |
| Kebabya 2021^67^ | REM vs SoC | Kenya | Parallel | Moderate | 3 | 4 | Phone-based reminders | Phone‑based reminders supporting PMTCT adherence |
| Hightow-Weidman 2023^68^ | mHealth vs SoC | USA | Parallel | Low | 5 | 11 | Epic‑Allies app | Gamified mobile app supporting engagement and adherence among youth |
| Lewis 2022^69^ | mHealth vs SoC | USA | Parallel | Moderate | 5 | 13 | Positive Health Check | Tailored video visits supporting viral suppression and retention |
| Chang 2023^70^ | mHealth vs SoC | Nigeria | Parallel | Moderate | 3 | 24 | Point-of-care VL testing | Point‑of‑care VL results guiding clinical actions |
| Fahey 2022^71^ | EI vs SoC | Tanzania | Parallel | Low | 3 | 72 | Conditional incentives | Short‑term incentives supporting retention and adherence |
| Metsch 2021^72^ | CM vs SoC | USA | Parallel | Low | 2 | 23 | Care facilitation (CF) | Strengths‑based case management for complex needs |
| Amone 2024^73^ | DSD vs SoC | Uganda | Parallel | Low | 3 | 48 | Friends for Life Circles | Support circles facilitating adherence and retention |
| Ayieko 2023^74^ | SUP vs SoC | Dominican Republic | Cluster | Low | 3 | 12 | Nutrition counselling + gardening + cooking | Community‑partnered nutrition support |
| Naggirinya 2024^75^ | mHealth vs SoC | Uganda | Parallel | Single | 5 | 16 | Linkage intervention | Case management with SMS accelerating post‑discharge linkage |
| Derose 2022^76^ | EI vs SoC | Dominican Republic | Cluster | Low | 3 | 12 | Dynamic choice travel | Travel support offsetting mobility barriers |
| Horvath 2024^42^ | mHealth vs SoC | USA | Parallel | Moderate | 3 | 41 | Nutrition counselling + gardening + cooking | Community‑partnered strategies supporting engagement |
| Inghels 2024^77^ | EI vs mHealth | South Africa | Cluster | High | 5 | 12 | Conditional financial incentives | Conditional incentives supporting retention |
| Luoma 2023^78^ | EDU vs eSoC | Russia | Parallel | Low | 3 | 11 | Stigma group sessions | Stigma‑focused counselling reduces care avoidance |
| Mabuto 2024^79^ | SUP vs SoC | South Africa | Parallel | Moderate | 3 | 9 | Transitional community adherence support | Transitional adherence support after incarceration |
| Martin 2024^80^ | BC vs SoC | USA | Parallel | Low | 3 | 21 | InstaCare (MI + rapid ART start) | Motivational counselling with rapid ART start |
| Ndongo 2024^81^ | SUP vs SoC | Cameroon | Parallel | Low | 3 | 18 | CME‑FCB | Mentoring and feedback supporting retention |
| Njau 2024^82^ | EI vs eSoC | Tanzania | Cluster | Low | 3 | 25 | Financial incentives | Incentives promoting viral suppression and retention |
| Njuguna 2024^83^ | REM vs SoC | South Africa | Multi-arm | High | 3 | 9 | Fresh‑start SMS | Fresh‑start timed SMS nudging adherence |
| Novak 2023^84^ | EI vs SoC | USA | Parallel | High | 5 | 58 | Financial incentive | Incentives for reinforcing medication adherence |
| Onoya 2024^85^ | BC vs SoC | South Africa | Cluster | Moderate | 4 | 17 | Thusa‑Thuso MI training | Counsellor MI training supporting ART uptake and retention |
| Palar 2024^86^ | EDU vs EI | USA | Parallel | High | 2 | 14 | Medically tailored meals + education | Food support addressing insecurity and strengthening engagement |
| Marc 2022^87^ | MM vs SoC | Haiti | Pilot | Low | 5 | 31 | DSD models (MMD; spacing; decentralization) | DSD strategies reducing visit burden |
| Parry 2023^88^ | BC vs SoC | South Africa | Parallel | Low | 3 | 17 | Psychological intervention | Brief alcohol counselling reduces hazardous drinking |
| Peck 2024^89^ | BC vs SoC | Tanzania | Parallel | Low | 5 | 48 | Linkage intervention | Post‑discharge case management supporting ART initiation and follow‑up |
| Reid 2017^90^ | REM vs SoC | Botswana | Parallel | High | 5 | 15 | SMS for pharmacy pickups | SMS reminders supporting timely ART refills |
| Samet 2023^91^ | CM vs SoC | Russia | Parallel | Moderate | 2 | 10 | LINC‑II case management | Strengths‑based case management supporting engagement |
| Solomon 2024^92^ | EI vs eSoC | India | Cluster | Moderate | 3 | 11 | Voucher incentives | Voucher incentives promoting linkage and ART initiation |
| Zani 2024^93^ | SUP vs TS | South Africa | Cluster | Moderate | 5 | 15 | Strengthened APC training | Lay counsellor training supporting psychosocial care |
| Giovenco 2024^94^ | REM vs SoC | South Africa | Pilot | Moderate | 5 | 12 | SMS adherence support | SMS service models supporting youth adherence |
| Limbada 2021^95^ | DSD vs SoC | Zambia/ South Africa | Cluster | Low | 5 | 24 | Adherence clubs | DSD group models managing growing ART caseloads |

Pragmatism was scored using the RITES tool (Rating of Included Trials on the Efficacy–Effectiveness Spectrum). A higher score indicates greater pragmatism (1 = more explanatory, 5 = more pragmatic).^96^

Intervention intensity was classified according to Montgomery et al. as: single session (one-time contact), low (minimal or infrequent contact), moderate (periodic contact), high (frequent or structured contact), and continuous (ongoing delivery throughout follow-up).^97^

#### Table S4. Study eligibility characteristics and baseline clinical status of included trials

| Author (Year) | Age category | Key population groups | Female (n) | Randomised (n) | Baseline HIV labs (VL/CD4) | Baseline health status | HIV care cascade status | Other eligibility criteria |
| --- | --- | --- | --- | --- | --- | --- | --- | --- |
| Mbuagbaw 2012^13^ | Adults | General | 147 | 200 | NR | NR | On ART (≥ 1 month) | Mobile phone access |
| Lucas 2010^14^ | Adults | General | 26 | 96 | NR | Opioid use disorder | NR | Positive urine opioid test |
| Keitz 2001^15^ | Adults | General | 83 | 214 | NR | NR | NR | Ambulatory HIV care at DUMC; Uninsured |
| Odeny 2014^16^ | Adults | Women | 388 | 388 | NR | Pregnancy | NR | Mobile phone access |
| Chang 2010^17^ | Adults | General | 885 | 1336 | NR | NR | NR | NR |
| Wohl 2011^18^ | Adults | Incarcerated | 24 | 104 | NR | NR | NR | Incarcerated pre-release |
| Naar-King 2009^19^ | Adolescent | Youth/ACB | 35 | 87 | NR | NR | Enrolled in HIV care | NR |
| Gwadz 2015^20^ | Adults | ACB | 37 | 95 | CD4 ≥ 500 cells/mm³ | NR | ART‑naive or off ART (≤30 mon); active in HIV care | Ethnicity/ race targeted |
| Norton 2014^21^ | Adolescents + Adults | ACB | 13 | 52 | NR | NR | NR | Mobile phone access |
| MacGowan 2014^22^ | Adults | Prisoners/ MSM | 5 | 73 | NR | NR | NR | Incarcerated pre-release |
| Gardner 2014^23^ | Adults | General | 665 | 1838 | NR | NR | Prior missed visit | NR |
| Konkle-Parker 2014^24^ | Adults | General | 51 | 100 | NR | NR | Documented medication adherence or retention problem | NR |
| Huang 2013^25^ | Adults | General | 75 | 196 | CD4 < 350 cells/mm³ | WHO stage III/IV | NR | Mobile phone access |
| Wamalwa 2009^26^ | Peadiatric | General | 51 | 99 | CD4 < 15% | WHO moderate–severe HIV‑1 disease | ART‑naive | Caregiver literacy |
| Wohl 2006^27^ | Adults | PWID | 58 | 250 | VL ≥ 400 copies/mL | NR | Initiated ART (≤ 6 mon) | MMSE ≥23 |
| Chander 2015^28^ | Adults | Women/ PWID | 148 | 148 | NR | Hazardous alcohol use (TWEAK ≥2 or thresholds) | NR | NR |
| Kunutsor 2011^29^ | Adults | General | 118 | 174 | NR | NR | On ART; receiving routine adherence support | NR |
| Dulli 2020^30^ | Adolescents + Adults | Youth/ACB | 306 | 349 | NR | NR | On ART (≤12 mon) | Basic literacy for web chats |
| Ammassari 2018^31^ | Adults | General | 90 | 774 | VL > 1000 copies/mL; CD4 < 500 cells/µL | NR | NR | No major IAS‑USA resistance on genotype |
| Grave 2018^32^ | Peadiatric+ Adolescent | Youth | 2540 | 4420 | NR | NR | Active in HIV care during study period | NR |
| El-Sadr 2019^33^ | Adults | General | 25891 | 51782 | NR | NR | NR | NR |
| Fahey 2020^34^ | Adults | General | 330 | 530 | NR | NR | Initiated ART (≤ 1 mon) | NR |
| Myer 2018^35^ | Adults | Women | 471 | 471 | NR | Women with children | NR | NR |
| McLaughlin 2018^36^ | Adults | General | 128 | 356 | NR | Poverty (PPI <45) | Initiated ART | NR |
| Kadota 2018^37^ | Adults | General | 509 | 805 | NR | Food insecurity (Household Hunger Scale) | Initiated ART (≤ 3 mon) | NR |
| Kalichman 2018^38^ | Adults | General | 117 | 500 | NR | NR | NR | Sexually active |
| Neduzhko 2020^39^ | Adults | General | 100 | 276 | NR | Newly diagnosed HIV | NR | NR |
| Kim 2019^40^ | Adults | Women | 306 | 306 | NR | Pregnancy | ART naive | NR |
| Mavhu 2020^41^ | Adolescent | General | 257 | 608 | NR | NR | On ART or initiating ART | NR |
| Horvath 2019^42^ | Adults | PWID + MSM | 0 | 90 | NR | Stimulant use (past 6 months) | On ART, suboptimal adherence | Mobile phone access |
| Samet 2019^43^ | Adults + Older adults | PWID | 93 | 349 | NR | History of injection drug use | Willing to receive HIV care | Mobile phone access; Hospitalized (narcology) |
| Sherman 2020^44^ | Adults | General | 35 | 94 | NR | NR | ART‑naive or initiating (≤ 1 mon) | Mobile phone access |
| Willis 2019^45^ | Pediatric + Adolescent | Youth | 61 | 100 | NR | NR | On ART; Aware of HIV status | NR |
| Sabin 2020^46^ | Adults | General | 133 | 133 | NR | Pregnancy | ART‑naive; Initiating ART | Mobile phone access |
| Pascoe 2019^47^ | Adults | General | 426 | 730 | CD4 < 500 cells/mm³ | WHO stage III/IV | Initiating ART | NR |
| Silverman 2019^48^ | Adults | General | 47 | 102 | VL > 200 copies/mL | Newly diagnosed HIV | Not in HIV care (≥12 weeks) | NR |
| Goodrich 2021^49^ | Adults | General | 327 | 420 | CD4 ≥ 200 cells/µL; VL < 40 copies/mL | NR | On ART (≥ 6 mon) | NR |
| Bynonanebye 2021^50^ | Adults | General | 413 | 600 | NR | NR | NR | NR |
| Hoffman 2021^51^ | Adults | General | 5774 | 8719 | NR | NR | NR | NR |
| Cassidy 2020^52^ | Adults | General | 1659 | 2150 | VL < 400 copies/mL | NR | On ART (> 6 months); Enrolled in adherence club model | NR |
| Kinuthia 2021^53^ | Adolescents + Adults | Women | 0 | 824 | NR | NR | NR | Mobile phone access |
| Ayer 2021^54^ | Adults | General | 208 | 468 | NR | NR | On first-line ART (UTT) | Mobile phone access |
| Cedric H 2021^55^ | Adults | General | 12 | 29 | VL > 400 copies/mL | NR | On first‑ or second‑line ART | NR |
| Graham 2021^56^ | Adults | MSM | 0 | 60 | NR | NR | Initiating ART | NR |
| Ndhlovu 2021^57^ | Adults | General | 105 | 212 | VL > 400 copies/mL | NR | NR | NR |
| Fatti 2020^58^ | Adults | ACB | 3463 | 4800 | VL < 1000 copies/mL | Weight ≥35 kg | On first‑line ART (> 6 mon) | NR |
| Wagner 2021^59^ | Adults | General | 24 | 176 | Detectable VL; CD4 < 200 cells/mm³ | NR | Starting or restarting ART; Stable health | NR |
| Stephenson 2021^60^ | Adults | ACB | 0 | 159 | NR | Stable patient | NR | NR |
| Tukei 2020^61^ | Adults | ACB | 3539 | 5336 | VL < 1000 copies/mL | NR | On first‑line ART (≥ 6 mon) | NR |
| Roy 2020^62^ | Adolescents + Adults | ACB | 678 | 1160 | CD4 ≥ 200 cells/mm³ | Not acutely ill | On ART (> 6 mon) | NR |
| Drain 2021^63^ | Adults | MSM | 235 | 390 | NR | NR | ART initiated (>6 mon) | NR |
| Giordona 2016^64^ | Adults | Women | 120 | 460 | NR | NR | Not in HIV care | NR |
| Liu 2022^65^ | Adults | General | 25 | 112 | NR | NR | NR | NR |
| Hickey 2021^66^ | Adults | General | 207 | 304 | NR | NR | Missed clinic visit > 3 days | NR |
| Kebabya 2021^67^ | Adults | Women | 150 | 150 | NR | Postpartum women | NR | Mobile phone access |
| Hightow-Weidman 2023^68^ | Adolescents + Adults | Youth | 0 | 146 | Detectable VL | NR | NR | Mobile phone access |
| Lewis 2022^69^ | Adults | General | 189 | 799 | VL 200 copies/mL | NR | Receiving care; New to care (≤ 12 mon) or out of care (> 12 mon) | NR |
| Chang 2023^70^ | Adults | General | 349 | 541 | NR | Newly diagnosed HIV | NR | NR |
| Fahey 2022^71^ | Adults | General | 509 | 800 | NR | Food insecure (Household Hunger Scale) | Initiated ART (≤ 3 mon) | NR |
| Metsch 2021^72^ | Adults | General | 47 | 113 | Detectable VL | OUD/stimulant/heavy alcohol (prior 12 months); AIDS‑defining illness | NR | NR |
| Amone 2024^73^ | Adults | Women | 540 | 540 | NR | Pregnancy (Option B+) | Initiating ART (PMTCT) | NR |
| Ayieko 2023^74^ | Adults | General | 58 | 109 | Detectable VL | Food insecurity; Adherence/  engagement problems | On ART (≥ 6 mon); Missed clinic or refill visit (past 6 mon) | NR |
| Naggirinya 2024^75^ | Adolescents + Adults | General | 167 | 206 | NR | NR | Initiating ART or on ART ≤ 6 months; Youth 15–24 | NR |
| Derose 2022^76^ | Adults | General | 58 | 109 | Detectable VL | Food insecurity | On ART ≥ 6 mon;  Adherence < 90%; clinic‑registered | NR |
| Horvath 2024^42^ | Adults | ACB + MSM | 0 | 401 | NR | NR | Adherence < 90% | Mobile phone access |
| Inghels 2024^77^ | Adults | General | 10805 | 15680 | NR | NR | Not on ART | NR |
| Luoma 2023^78^ | Adults | PWID | 49 | 100 | NR | Injection drug use | Not on ART | Mobile phone access |
| Mabuto 2024^79^ | Adults | Prisoners | 9 | 175 | NR | NR | On ART | Scheduled for release |
| Martin 2024^80^ | Adults | General | 46 | 51 | VL ≥ 200 copies | NR | Out of care or off ART (≥ 2 weeks); ≥ 2 missed appointments (18 mon) | NR |
| Ndongo 2024^81^ | Adolescents | Youth | 162 | 305 | NR | NR | NR | NR |
| Njau 2024^82^ | Adults | ACB | 1185 | 1990 | NR | NR | Initiating ART (< 1 mon) | Mobile phone access |
| Njuguna 2024^83^ | Adolescents + Adults | Youth | 5566 | 9630 | VL ≥ 200 copies/mL | NR | Not in HIV care (≥12 weeks); missed last ART visit (>28 days) | Mobile phone access |
| Novak 2023^84^ | Adults | General | 47 | 102 | VL ≥ 200 copies/mL | Newly diagnosed HIV | Not in HIV care (≥ 3 mon) | NR |
| Onoya 2024^85^ | Adults | General | 335 | 548 | NR | Newly diagnosed HIV | NR | NR |
| Palar 2024^86^ | Adults | General | 36 | 191 | NR | NR | NR | Income < 200% FPL |
| Marc 2022^87^ | Adults | General | 110 | 245 | NR | WHO Stage 1–2 | NR | NR |
| Parry 2023^88^ | Adults | General | 358 | 623 | NR | Harmful/hazardous drinking (AUDIT‑C thresholds) | On ART (≥ 3 mon) | NR |
| Peck 2024^89^ | Adults | General | 384 | 500 | NR | NR | ART‑naive or off ART (> 7 days) | Mobile phone access |
| Reid 2017^90^ | Adults | General | 48 | 108 | NR | OUD; History of IDU | On ART; Receiving HIV care at the clinic | NR |
| Samet 2023^91^ | Adults | PWID | 90 | 225 | NR | OUD; History of IDU | Not on ART (< mon) | NR |
| Solomon 2024^92^ | Adults | General | 154 | 1200 | NR | NR | ART‑naive or on ART (<12 mon) |  |
| Zani 2024^93^ | Adults | ACB | 1642 | 2002 | NR | Depression (PHQ-9≥ 9) | On ART | NR |
| Giovenco 2024^94^ | Adults | Youth | 124 | 215 | NR | NR | On ART (> 12 mon) | Mobile phone access |
| Limbada 2021^95^ | Adults | General | 1757 | 2489 | VL < 1000 copies/mL | Stable disease | On firstline ART (≥ 6 mon) | NR |

Age categories were defined as pediatric (0–12 years), adolescent (13–17 years), adult (18–49 years), and older adult (≥50 years), consistent with WHO and UNAIDS definitions used in HIV research.^98^

Key population categories follow WHO definitions: MSM, PWID, sex workers, transgender people, and people in prisons/closed settings.^99^

Abbreviations: ACB = African, Caribbean, or Black; ART = antiretroviral therapy; AUDIT‑C = Alcohol Use Disorders Identification Test – Consumption; CD4 = CD4 lymphocyte count; CHW = community health worker; cART = combination antiretroviral therapy; DOT = directly observed therapy; FPL = Federal Poverty Level; HHS = Household Hunger Scale; IDU = injection drug use; MMSE = Mini‑Mental State Examination; mon = months; MSM = men who have sex with men; NR = not reported; OUD = opioid use disorder; PHQ‑9 = Patient Health Questionnaire‑9; PMTCT = prevention of mother‑to‑child transmission; PWID = people who inject drugs; PPI = Progress out of Poverty Index; UTT = universal test and treat; VL = viral load; WHO stage = World Health Organization clinical staging.

### Meta‑analysis tables: Direct estimates, network estimates, and P‑scores

#### Table S5. Direct evidence from fixed‑effects pairwise meta‑analyses (OR > 1 favours first‑named intervention)

| Comparisons | No. of studies (k) | OR (95 %CI) | | Q | | I^2^ |
| --- | --- | --- | --- | --- | --- | --- |
| mHealth vs SoC | 14 | 1.00 (0.88 to 1.14) | 24.5 | | | 47 % |
| TS vs CM | 1 | 4.18 (1.73 to 10.06) | 0 | | | NA |
| TS vs SoC | 2 | 1.34 (0.85 to 2.09) | 4.0 | | | 75 % |
| mHealth vs REM | 2 | 1.34 (0.91 to 1.99) | 2.1 | | | 52 % |
| SoC vs SUP | 2 | 1.35 (0.88 to 2.08) | 1.4 | | | 31 % |
| BC vs DSD | 1 | 1.19 (0.30 to 4.75) | 0 | | | NA |
| TS vs BC | 1 | 0.92 (0.34 to 2.48) | 0 | | | NA |
| BC vs eSoC | 1 | 1.60 (0.71 to 3.61) | 0 | | | NA |
| REM vs SoC | 8 | 1.16 (0.97 to 1.39) | 29.7 | | | 76 % |
| BC vs CM | 1 | 3.18 (0.93 to 10.90) | 0 | | | NA |
| EDU vs BC | 1 | 1.06 (0.82 to 1.36) | 0 | | | NA |
| EDU vs REM | 1 | 1.31 (1.02 to 1.67) | 0 | | | NA |
| BC vs REM | 1 | 1.24 (0.97 to 1.58) | 0 | | | NA |
| BC vs SoC | 6 | 1.56 (1.27 to 1.91) | 41.4 | | | 87 % |
| REM vs EDU | 2 | 1.88 (1.32 to 2.69) | 0.1 | | | 0.00 % |
| DSD vs SoC | 8 | 1.53 (1.24 to 1.87) | 34.1 | | | 79 % |
| DSD vs SUP | 2 | 0.95 (0.63 to 1.43) | 0 | | | 0.0 % |
| SUP vs SoC | 5 | 1.68 (1.23 to 2.29) | 8.9 | | | 55 % |
| MM vs SoC | 2 | 0.70 (0.55 to 0.88) | 0.1 | | | 0.0 % |
| EDU vs SoC | 1 | 0.71 (0.37 to 1.36) | 0 | | NA | |
| EI vs SoC | 6 | 1.44 (1.13 to 1.84) | 21.9 | | 77 % | |
| BC vs EDU | 2 | 1.34 (0.84 to 2.14) | 0.3 | | 0 % | |
| SUP vs eSoC | 1 | 3.86 (1.67to 8.91) | 0 | | NA | |
| MMD vs SoC | 1 | 2.02 (1.32 to 3.09) | 0 | | NA | |
| DSD vs DSD+MMD | 2 | 1.26 (0.80 to 1.98) | 0.5 | | 0 % | |
| EI vs BC | 1 | 0.18 (0.02 to 1.85) | 0 | | NA | |
| SUP vs BC | 2 | 0.93 (0.64 to 1.36) | 0.7 | | 0 % | |
| SoC vs DSD+MMD | 1 | 0.96 (0.52 to 1.78) | 0 | | NA | |
| DSD+MMD vs DSD | 1 | 0.59 (0.14 to 2.45) | 0 | | NA | |
| DSD+MMD vs SoC | 1 | 0.48 (0.12 to 1.97) | 0 | | NA | |
| DSD vs eSoC | 1 | 2.88 (1.15 to 7.23) | 0 | | NA | |
| CM vs SoC | 2 | 2.63 (1.60 to 4.30) | 7.7 | | 87 % | |
| mHealth +EI vs EI | 1 | 1.36 (0.83 to 2.23) | 0 | | NA | |
| mHealth +EI vs mHealth | 1 | 1.01 (0.62 to 1.64) | 0 | | NA | |
| mHealth +EI vs SoC | 1 | 1.10 (0.73 to 1.68) | 0 | | NA | |
| EI vs mHealth | 1 | 0.74 (0.46 to 1.17) | 0 | | NA | |
| EDU vs eSoC | 1 | 4.24 (1.15 to 15.61) | 0 | | NA | |
| SoC vs BC | 1 | 1.26 (0.40 to 3.93) | 0 | | NA | |
| EI vs eSoC | 1 | 1.62 (1.03 to 2.56) | 0 | | NA | |
| EI vs EDU | 1 | 1.21 (0.50 to 2.90) | 0 | | NA | |
| SoC vs REM | 1 | 0.41 (0.16 to 1.07) | 0 | | NA | |
| SUP vs TS | 1 | 0.53 (0.31 to 0.91) | 0 | | NA | |

Q = Cochran’s Q statistic for heterogeneity within each direct comparison; I² = percentage of variability due to heterogeneity rather than chance. All estimates are fixed‑effect

NA indicates heterogeneity statistics were not estimable or not applicable (e.g., k = 1)

### Pairwise subgroup analysis (Interaction tests: p‑values)

#### Table S6. Pairwise meta‑analysis by population group (fixed‑effect model) (n=78)

| Population group | m (estimates) | Pooled OR (95% CI) | I² (%) |
| --- | --- | --- | --- |
| General | 69 | 1.21 (1.14 to 1.28) | 72.6 |
| Key | 24 | 1.23 (1.08 to 1.40) | 59.3 |
| Overall | 93 | 1.21 (1.16 to 1.28) | 69.9 |

General versus key population classification followed study reporting.^99^

Q=305.10, df=92, p<0.0001; I²=69.9%.
Test for subgroup differences: Q_between=0.05, df=1, p=0.820

#### Table S7. Pairwise meta‑analysis by WHO region (fixed‑effect model) (n=78)

| WHO region | k | Pooled OR (95% CI) | I² (%) |
| --- | --- | --- | --- |
| AFRO | 54 | 1.26 (1.17 to 1.35) | 70.8 |
| AMRO | 32 | 1.11 (1.00 to 1.22) | 54.7 |
| EURO | 5 | 1.15 (0.95 to 1.38) | 91.0 |
| WPRO | 1 | 1.64 (0.61 to 4.43) | NA |
| SEARO | 1 | 1.92 (1.31 to 2.81) | NA |
| Overall | 93 | 1.21 (1.15 to 12.28 | 69.9 |

Subgroup analysis for WHO geographical regions.^100^

Overall Q=305.10, df=92, p<0.0001; I²=69.9%.
Test for subgroup differences: Q_between=10.66, df=4, p=0.031.

#### ****Table S8. Pairwise meta‑analysis by World Bank income group (fixed‑effect model) (n=78)****

| Income group | k | Pooled OR (95% CI) | I² (%) |
| --- | --- | --- | --- |
| Low‑income (LIC) | 13 | 1.65 (1.41 to 1.93) | 63.8 |
| Lower‑middle income (LMIC) | 23 | 1.30 (1.14 to 1.48) | 73.5 |
| Upper‑middle income (UMIC) | 23 | 1.19 (1.08 to 1.31) | 71.8 |
| High‑income (HIC) | 34 | 1.07 (0.98 to 1.18) | 62.2 |

Country income level was classified using the World Bank income groups.^101^

**Overall (common‑effect):** OR 1.21 (95% CI 1.15 to 1.28); Q=305.10, df=92, p<0.0001; I²=69.9%.
**Test for subgroup differences:** Q_between=23.50, df=3, **p<0.0001**.

#### ****Table S9. Pairwise meta‑analysis by follow-up time (fixed‑effect model)**** (n=78)

| Follow-up time (months) | k | Pooled OR (95% CI) | I² (%) |
| --- | --- | --- | --- |
| 3 | 13 | 1.11 (0.95 to 1.29) | 54.6 |
| 6 | 23 | 1.27 (1.13 to 1.43) | 74.7 |
| 9 | 2 | 0.54 (0.34 to 0.86) | 26.0 |
| 12 | 52 | 1.23 (1.15 to 1.32) | 70.3 |
| 15 | 2 | 1.43 (0.88 to 2.32) | 77.4 |
| 24 | 1 | 1.08 (0.48 to 2.46) | — |

**Overall (common‑effect):** OR 1.21 (95% CI 1.15 to 1.28); Q=305.10, df=92, p<0.0001; I²=69.9%.
**Test for subgroup differences:** Q_between=14.37, df=5, **p=0.013.**

#### ****Table S10. Pairwise meta‑analysis by trial risk of bias (fixed‑effect model) (n=78)****

| Risk of bias group | k | Pooled OR (95% CI) | I² (%) |
| --- | --- | --- | --- |
| **Definitely low** | 24 | 1.37 (1.25 to 1.50) | 55.8 |
| **Probably low** | 34 | 1.40 (1.24 to 1.58) | 68.9 |
| **Probably high** | 31 | 1.01 (0.93 to 1.11) | 72.1 |
| **Definitely high** | 4 | 0.80 (0.44 to 1.45) | 65.5 |

**Overall (common‑effect):** OR **1.21** (95% CI **1.15 to 1.28);** Q=305.10, df=92, p<0.0001; I²=69.9%.
**Test for subgroup differences:** Q_between=30.87, df=3, **p<0.0001**.

Table S11. Pairwise meta‑analysis by trial design (fixed‑effect model) ****(n=78)****

| Trial design | k | Pooled OR (95% CI) | I² (%) |
| --- | --- | --- | --- |
| Parallel | 70 | 1.25 (1.17 to 1.33) | 68.8 |
| Multiarm | 23 | 1.11 (0.99 to 1.23) | 72.7 |

Overall (common‑effect): OR 1.21 (95% CI 1.15 to 1.28), Q=305.10, df=92, p<0.0001, I²=69.9%.
Test for subgroup differences: Q_between=3.63, df=1, p=0.0569.

Table S12. Pairwise meta-analysis by trial pragmatism (fixed‑effect model) (n=78)

| Pragmatism category | k | Pooled OR (95% CI) | I² (%) |
| --- | --- | --- | --- |
| Effective | 55 | 1.19 (1.11 to 1.28) | 66.9 |
| Efficacious | 3 | 0.84 (0.66 to 1.05) | 87.9 |
| Both | 33 | 1.32 (1.21 to 1.45) | 67.1 |
| Definitely effective | 2 | 1.21 (0.86 to 1.69) | 93.2 |

Overall (common‑effect): OR 1.21 (95% CI 1.15 to 1.28); Q=305.10, df=92, p<0.0001; I²=69.9%.
Test for subgroup differences: Q_between=13.64, df=3, p=0.0034.

### Table S13. Direct evidence for viral load suppression from pairwise meta‑analyses

| Comparisons | k | OR (95%-CI) | Q | I^2 |
| --- | --- | --- | --- | --- |
| MM vs SoC | 1 | 0.93 (0.71 to 1.22) | 0 | NA |
| DSD vs SoC | 8 | 1.38 (1.19 to 1.59) | 54.2 | 87% |
| mHealth vs SoC | 9 | 1.10 (0.93 to 1.30) | 11.3 | 29% |
| DSD vs DSD+MMD | 2 | 2.74 (1.46 to 5.15) | 4.8 | 79% |
| BC vs SoC | 4 | 1.51 (1.18 to 1.94) | 4.0 | 0.26 |
| SoC vs SUP | 1 | 1.22 (0.68 to 2.18) | 0.0 | NA |
| TS vs SoC | 1 | 2.85 (1.45 to 5.61) | 0.0 | NA |
| EI vs SoC | 4 | 1.19 (0.87 to 1.64) | 25.9 | 0.88 |
| SoC vs DSD+MMD | 1 | 8.16 (2.36 to 28.28) | 0.0 | NA |
| EDU vs REM | 1 | 0.56 (0.55 to 0.57) | 0.0 | NA |
| SUP vs BC | 2 | 1.37 (0.94 to 1.99) | 0.8 | 0.00 |
| REM vs SoC | 5 | 1.06 (0.72 to 1.58) | 12.1 | 0.67 |
| BC vs eSoC | 0 | NA | NA | NA |
| EI vs mHealth | 1 | 0.60 (0.31 to 1.14) | 0.00 | NA |
| mHealth vs REM | 1 | 1.63 (0.96 to 2.75) | 0.00 | NA |
| SoC vs BC | 1 | 1.38 (0.36 to 5.34) | 0.00 | NA |
| DSD vs SUP | 2 | 0.96 (0.66 to 1.40) | 0.1 | 0.00 |
| SUP vs SoC | 3 | 1.23 (0.89 to 1.69) | 4.7 | 0.58 |
| EI vs EDU | 1 | 1.99 (0.76 to 5.2) | 3.2 | NA |
| EDU vs SoC | 1. | 1.33 (0.88 to 2.00) | 0.0 | NA |
| EI vs eSoC | 1 | 1.79 (1.06 to 3.01) | 0 | NA |
| BC vs EDU | 1 | 1.62 (0.75 to 3.51) | 0 | NA |
| DSD+MMD vs DSD | 1 | 1.14 (0.46 to 2.82) | 0 | NA |
| DSD+MMD vs SoC | 1 | 0.81 (0.33 to 2.03) | 0 | NA |
| SUP vs TS | 1 | 1.00 (0.48 to 2.06) | 0 | NA |

Q = Cochran’s Q statistic for heterogeneity within each direct comparison; I² = percentage of variability due to heterogeneity rather than chance. All estimates are fixed‑effect

NA indicates heterogeneity statistics were not estimable or not applicable (e.g., k = 1)

#### Table S14. Network meta-analysis league table for retention in HIV care

| TS | 1.04 (0.61 to 1.77) | 0.75 (0.53 to 1.08) | 0.77 (0.48 to 1.23) | 0.70 (0.50 to 0.97) | 0.68 (0.48 to 0.97) | 0.67 (0.48 to 0.93) | 0.63 (0.38 to 1.04) | 0.59 (0.42 to 0.83) | 0.58 (0.40 to 0.83) | 0.56 (0.34 to 0.93) | 0.54 (0.39 to 0.76) | 0.51 (0.38 to 0.70) | 0.36 (0.23 to 0.57) | 0.36 (0.24 to 0.53) |
| --- | --- | --- | --- | --- | --- | --- | --- | --- | --- | --- | --- | --- | --- | --- |
| 0.96 (0.57 to 1.63) | MMD | 0.73 (0.46 to 1.15) | 0.74 (0.41 to 1.33) | 0.67 (0.43 to 1.05) | 0.66 (0.42 to 1.03) | 0.64 (0.41 to 1.02) | 0.60 (0.34 to 1.08) | 0.57 (0.36 to 0.89) | 0.55 (0.35 to 0.88) | 0.54 (0.30 to 0.97) | 0.52 (0.34 to 0.81) | 0.49 (0.32 to 0.76) | 0.35 (0.21 to 0.60) | 0.34 (0.21 to 0.56) |
| 1.32 (0.93 to 1.90) | 1.38 (0.87 to 2.19) | DSD | 1.02 (0.65 to 1.59) | 0.93 (0.75 to 1.15) | 0.90 (0.71 to 1.15) | 0.89 (0.71 to 1.12) | 0.83 (0.53 to 1.28) | 0.78 (0.62 to 0.98) | 0.76 (0.59 to 0.99) | 0.74 (0.50 to 1.09) | 0.72 (0.58 to 0.90) | 0.68 (0.57 to 0.82) | 0.48 (0.34 to 0.69) | 0.48 (0.35 to 0.64) |
| 1.30 (0.81 to 2.10) | 1.36 (0.75 to 2.45) | 0.98 (0.63 to 1.54) | CM | 0.91 (0.60 to 1.40) | 0.89 (0.57 to 1.38) | 0.87 (0.56 to 1.36) | 0.81 (0.46 to 1.45) | 0.77 (0.50 to 1.19) | 0.75 (0.48 to 1.18) | 0.73 (0.41 to 1.30) | 0.71 (0.46 to 1.09) | 0.67 (0.44 to 1.01) | 0.47 (0.28 to 0.80) | 0.47 (0.29 to 0.75) |
| 1.43 (1.03 to 1.99) | 1.49 (0.95 to 2.32) | 1.08 (0.87 to 1.34) | 1.10 (0.72 to 1.68) | BC | 0.97 (0.80 to 1.19) | 0.96 (0.79 to 1.17) | 0.89 (0.59 to 1.36) | 0.84 (0.72 to 0.99) | 0.82 (0.69 to 0.99) | 0.80 (0.52 to 1.22) | 0.78 (0.65 to 0.92) | 0.73 (0.65 to 0.83) | 0.52 (0.37 to 0.72) | 0.51 (0.39 to 0.67) |
| 1.46 (1.03 to 2.08) | 1.52 (0.97 to 2.40) | 1.11 (0.87 to 1.40) | 1.12 (0.72 to 1.75) | 1.03 (0.84 to 1.25) | EI | 0.98 (0.78 to 1.24) | 0.92 (0.60 to 1.39) | 0.87 (0.70 to 1.06) | 0.85 (0.67 to 1.07) | 0.82 (0.53 to 1.26) | 0.80 (0.65 to 0.97) | 0.75 (0.64 to 0.88) | 0.53 (0.39 to 0.73) | 0.53 (0.39 to 0.70) |
| 1.49 (1.07 to 2.07) | 1.55 (0.98 to 2.46) | 1.13 (0.90 to 1.42) | 1.14 (0.74 to 1.78) | 1.04 (0.86 to 1.27) | 1.02 (0.81 to 1.28) | SUP | 0.93 (0.60 to 1.44) | 0.88 (0.71 to 1.09) | 0.86 (0.68 to 1.10) | 0.83 (0.54 to 1.28) | 0.81 (0.66 to 1.00) | 0.77 (0.65 to 0.91) | 0.54 (0.38 to 0.77) | 0.53 (0.40 to 0.72) |
| 1.60 (0.96 to 2.66) | 1.66 (0.93 to 2.98) | 1.21 (0.78 to 1.87) | 1.23 (0.69 to 2.18) | 1.12 (0.74 to 1.70) | 1.09 (0.72 to 1.66) | 1.07 (0.69 to 1.66) | mHealth +EI | 0.95 (0.62 to 1.44) | 0.92 (0.60 to 1.43) | 0.89 (0.51 to 1.58) | 0.87 (0.58 to 1.31) | 0.82 (0.55 to 1.23) | 0.58 (0.35 to 0.96) | 0.57 (0.36 to 0.91) |
| 1.69 (1.20 to 2.38) | 1.76 (1.13 to 2.75) | 1.28 (1.02 to 1.60) | 1.30 (0.84 to 2.00) | 1.18 (1.01 to 1.39) | 1.16 (0.94 to 1.42) | 1.13 (0.91 to 1.41) | 1.06 (0.69 to 1.61) | REM | 0.98 (0.82 to 1.16) | 0.95 (0.62 to 1.45) | 0.92 (0.77 to 1.10) | 0.87 (0.76 to 1.00) | 0.62 (0.44 to 0.87) | 0.61 (0.46 to 0.80) |
| 1.73 (1.21 to 2.48) | 1.80 (1.14 to 2.86) | 1.31 (1.01 to 1.69) | 1.33 (0.85 to 2.08) | 1.21 (1.02 to 1.45) | 1.18 (0.94 to 1.49) | 1.16 (0.91 to 1.48) | 1.08 (0.70 to 1.68) | 1.02 (0.86 to 1.22) | EDU | 0.97 (0.62 to 1.51) | 0.94 (0.76 to 1.17) | 0.89 (0.75 to 1.07) | 0.63 (0.44 to 0.90) | 0.62 (0.46 to 0.84) |
| 1.79 (1.08 to 2.98) | 1.86 (1.04 to 3.35) | 1.35 (0.91 to 2.00) | 1.37 (0.77 to 2.45) | 1.25 (0.82 to 1.91) | 1.22 (0.79 to 1.88) | 1.20 (0.78 to 1.85) | 1.12 (0.63 to 1.97) | 1.06 (0.69 to 1.62) | 1.03 (0.66 to 1.61) | DSD+MMD | 0.97 (0.64 to 1.48) | 0.92 (0.61 to 1.38) | 0.65 (0.39 to 1.08) | 0.64 (0.40 to 1.03) |
| 1.84 (1.31 to 2.58) | 1.91 (1.23 to 2.98) | 1.39 (1.11 to 1.73) | 1.41 (0.92 to 2.17) | 1.29 (1.08 to 1.54) | 1.26 (1.03 to 1.53) | 1.23 (1.00 to 1.53) | 1.15 (0.76 to 1.73) | 1.09 (0.91 to 1.30) | 1.06 (0.86 to 1.32) | 1.03 (0.67 to 1.57) | mHealth | 0.95 (0.84 to 1.07) | 0.67 (0.48 to 0.94) | 0.66 (0.50 to 0.86) |
| 1.94 (1.42 to 2.66) | 2.02 (1.32 to 3.09) | 1.47 (1.22 to 1.76) | 1.49 (0.99 to 2.25) | 1.36 (1.20 to 1.54) | 1.33 (1.13 to 1.55) | 1.30 (1.10 to 1.55) | 1.21 (0.82 to 1.81) | 1.15 (1.00 to 1.32) | 1.12 (0.94 to 1.34) | 1.09 (0.72 to 1.63) | 1.06 (0.93 to 1.20) | SoC | 0.71 (0.51 to 0.97) | 0.70 (0.55 to 0.88) |
| 2.75 (1.76 to 4.27) | 2.86 (1.68 to 4.86) | 2.07 (1.45 to 2.95) | 2.11 (1.25 to 3.54) | 1.92 (1.38 to 2.68) | 1.87 (1.37 to 2.56) | 1.84 (1.30 to 2.60) | 1.72 (1.04 to 2.84) | 1.62 (1.15 to 2.28) | 1.58 (1.11 to 2.26) | 1.53 (0.92 to 2.55) | 1.49 (1.06 to 2.10) | 1.41 (1.03 to 1.94) | eSoC | 0.98 (0.66 to 1.46) |
| 2.79 (1.88 to 4.14) | 2.90 (1.78 to 4.72) | 2.11 (1.56 to 2.84) | 2.14 (1.33 to 3.44) | 1.95 (1.49 to 2.56) | 1.90 (1.43 to 2.53) | 1.87 (1.39 to 2.51) | 1.74 (1.10 to 2.77) | 1.65 (1.25 to 2.17) | 1.61 (1.19 to 2.17) | 1.56 (0.97 to 2.49) | 1.52 (1.16 to 1.98) | 1.43 (1.13 to 1.82) | 1.02 (0.68 to 1.51) | MM |

Cells show odds ratio (OR) and 95% CI from the fixed‑effect network meta‑analysis

Lower triangle: row vs column; upper triangle: column vs row—values are reciprocals

OR > 1 favors the first‑named intervention.

Bold indicates statistical significance (CI excludes 1)

Diagonal indicates the same intervention; NE, non‑estimable

Interventions are ordered by P‑score (lower P‑score = worse)

#### Table S15. Network meta-analysis league table for viral load suppression

| SoC | 2.07  (1.25 to 3.43) | 1.43  (1.16 to 1.75) | 1.36  (1.11 to 1.67) | 1.34  (1.18 to 1.52) | 1.13  (0.84 to 1.52) | 0.96  (0.73 to 1.25) | 0.93  (0.71 to 1.22) | 0.92  (0.71 to 1.20) | 0.56  (0.55 to 0.57) | 0.58  (0.36 to 0.94) | 0.57  (0.33 to 1.00) |
| --- | --- | --- | --- | --- | --- | --- | --- | --- | --- | --- | --- |
| 0.48  (0.29 to 0.80) | TS | 0.69  (0.41 to 1.14) | 0.66  (0.39 to 1.12) | 0.65  (0.39 to 1.08) | 0.54  (0.30 to 0.98) | 0.46  (0.26 to 0.82) | 0.45  (0.25 to 0.80) | 0.44  (0.25 to 0.78) | 0.27  (0.16 to 0.45) | 0.28  (0.14 to 0.56) | 0.28  (0.13 to 0.59) |
| 0.70  (0.57 to 0.86) | 1.45  (0.88 to 2.41) | SUP | 0.96  (0.75 to 1.22) | 0.94 (0.75 to 1.18) | 0.79  (0.55 to 1.13) | 0.67  (0.48 to 0.94) | 0.65  (0.47 to 0.92) | 0.64  (0.46 to 0.90) | 0.40  (0.32 to 0.49) | 0.41  (0.24 to 0.68) | 0.40  (0.22 to 0.73) |
| 0.73  (0.60 to 0.90) | 1.52  (0.89 to 2.59) | 1.05  (0.82 to 1.34) | BC | 0.98  (0.78 to 1.25) | 0.83  (0.58 to 1.18) | 0.70  (0.51 to 0.97) | 0.68  (0.49 to 0.96) | 0.67  (0.49 to 0.93) | 0.41  (0.34 to 0.51) | 0.43  (0.26 to 0.72) | 0.42  (0.23 to 0.75) |
| 0.75  (0.66 to 0.85) | 1.55  (0.92 to 2.59) | 1.06  (0.85 to 1.33) | 1.02  (0.80 to 1.29) | DSD | 0.84  (0.61 to 1.16) | 0.71  (0.53 to 0.96) | 0.70  (0.52 to 0.94) | 0.69  (0.51 to 0.92) | 0.42  (0.37 to 0.48) | 0.44  (0.27 to 0.70) | 0.43  (0.24 to 0.76) |
| 0.89  (0.66 to 1.19) | 1.83  (1.02 to 3.29) | 1.26  (0.88 to 1.81) | 1.21  (0.85 to 1.72) | 1.19  (0.86 to 1.63) | EI | 0.85  (0.58 to 1.24) | 0.83  (0.55 to 1.23) | 0.81  (0.56 to 1.19) | 0.50  (0.37 to 0.67) | 0.52  (0.29 to 0.90) | 0.51  (0.31 to 0.83) |
| 1.05  (0.80 to 1.37) | 2.17  (1.23 to 3.83) | 1.49  (1.07 to 2.08) | 1.43  (1.03 to 1.97) | 1.40  (1.04 to 1.88) | 1.18  (0.80 to 1.74) | REM | 0.98  (0.67 to 1.43) | 0.96  (0.80 to 1.16) | 0.59  (0.45 to 0.77) | 0.61  (0.35 to 1.05) | 0.60  (0.33 to 1.10) |
| 1.07  (0.82 to 1.41) | 2.22  (1.25 to 3.94) | 1.53  (1.09 to 2.15) | 1.46  (1.04 to 2.05) | 1.44  (1.06 to 1.94) | 1.21  (0.81 to 1.81) | 1.03  (0.70 to 1.50) | MM | 0.99  (0.68 to 1.44) | 0.61  (0.46 to 0.79) | 0.62  (0.36 to 1.08) | 0.62  (0.33 to 1.14) |
| 1.09  (0.84 to 1.42) | 2.26  (1.28 to 3.98) | 1.55  (1.11 to 2.16) | 1.48  (1.08 to 2.04) | 1.46  (1.09 to 1.95) | 1.23  (0.84 to 1.79) | 1.04  (0.86 to 1.25) | 1.01  (0.70 to 1.48) | EDU | 0.61  (0.47 to 0.80) | 0.63  (0.37 to 1.09) | 0.62  (0.34 to 1.15) |
| 1.77  (1.74 to 1.80) | 3.67  (2.22 to 6.08) | 2.52  (2.05 to 3.11) | 2.41  (1.97 to 2.96) | 2.37  (2.08 to 2.70) | 2.00  (1.49 to 2.69) | 1.69  (1.30 to 2.21) | 1.65  (1.26 to 2.16) | 1.63  (1.25 to 2.12) | mHealth | 1.03  (0.64 to 1.66) | 1.02  (0.58 to 1.78) |
| 1.72  (1.07 to 2.76) | 3.56  (1.78 to 7.10) | 2.45  (1.47 to 4.09) | 2.34  (1.40 to 3.92) | 2.30  (1.44 to 3.67) | 1.94  (1.11 to 3.39) | 1.64  (0.95 to 2.83) | 1.60  (0.93 to 2.77) | 1.58  (0.92 to 2.72) | 0.97  (0.60 to 1.56) | DSD+MMD | 0.98  (0.47 to 2.05) |
| 1.74  (1.00 to 3.05) | 3.61  (1.70 to 7.66) | 2.49  (1.38 to 4.49) | 2.38  (1.33 to 4.26) | 2.33  (1.32 to 4.14) | 1.97  (1.20 to 3.22) | 1.67  (0.90 to 3.07) | 1.63  (0.87 to 3.02) | 1.60  (0.87 to 2.94) | 0.98  (0.56 to 1.72) | 1.02  (0.49 to 2.11) | eSoC |

Cells show odds ratio (OR) and 95% CI from the fixed‑effect network meta‑analysis

Lower triangle: row vs column; upper triangle: column vs row—values are reciprocals

OR > 1 favors the first‑named intervention.

Bold indicates statistical significance (CI excludes 1)

Diagonal indicates the same intervention; NE, non‑estimable

Interventions are ordered by P‑score (lower P‑score = worse)

#### Table S16. P-scores for retention in care

| Interventions | P-score |
| --- | --- |
| TS | 0.9392 |
| MMD | 0.9379 |
| DSD | 0.7648 |
| CM | 0.7297 |
| BC | 0.6736 |
| EI | 0.6302 |
| SUP | 0.5993 |
| mHealth +EI | 0.4985 |
| REM | 0.4126 |
| EDU | 0.3747 |
| DSD+MMD | 0.3632 |
| mHealth | 0.2899 |
| SoC | 0.2039 |
| eSoC | 0.0455 |
| MM | 0.0368 |

Higher P‑scores indicate improved retention

Aligned with Grade ratings

#### Table S17. P-scores for viral load suppression

| Interventions | P-score |
| --- | --- |
| TS | 0.9802 |
| SUP | 0.8453 |
| BC | 0.7992 |
| DSD | 0.7820 |
| SoC | 0.4760 |
| REM | 0.4305 |
| MM | 0.3983 |
| EDU | 0.3746 |
| DSD+MMD | 0.1114 |
| eSoC | 0.1106 |
| mHealth | 0.0843 |

Higher P‑scores indicate viral suppression

P‑scores placed TS, SUP, BC, and DSD among the highest-ranked interventions, but GRADE rated SUP and DSD with low or very low certainty because of inconsistency and incoherence, indicating that high statistical ranking did not always reflect high certainty of evidence

### Quality of life

Each comparison was informed by a single small study, we interpreted quality‑of‑life findings descriptively without pooling.

#### Table S18. General quality of life

| Study (year) | Comparison | TE (MD) | seTE | RoB | Follow-up (mos) |
| --- | --- | --- | --- | --- | --- |
| Mbuagbaw 2012 | mHealth vs SoC | 0.04 | 0.08 | Probably low | 6 |
| Giordano 2016 | SUP vs CBT | −2.06 | 2.50 | Probably high | 6 |
| Palar 2024 | EI vs EDU | −0.80 | 0.23 | Probably high | 6 |

TE is the mean difference (intervention − comparator) on a 0–100 scale (higher is better). One study per comparison; no pairwise or network meta-analysis was feasible. Estimates were interpreted descriptively.

#### Table S19. Mental health quality of life

| Study (year) | Comparison | TE (MD) | seTE | RoB | Follow‑up (mos) |
| --- | --- | --- | --- | --- | --- |
| Keitz 2001 | TS vs SoC | 1.60 | 0.17 | Definitely high | 12 |
| Bynonanebye 2021 | mHealth vs SoC | 0.20 | 0.42 | Definitely low | 12 |

TE is the mean difference (intervention − comparator) on a 0–100 scale (higher is better). TE and seTE were computed from arm‑level means and SDs. One study per comparison; no pairwise or network meta-analysis was feasible. Estimates were interpreted descriptively.

#### Table S20. Physical health quality of life

| Study (year) | Comparison | TE (MD) | seTE | RoB | Follow‑up (mos) |
| --- | --- | --- | --- | --- | --- |
| Keitz 2001 | TS vs SoC | 0.80 | 0.27 | Definitely high | 12 |
| Bynonanebye 2021 | mHealth vs SoC | −0.10 | 0.06 | Definitely low | 12 |
| Giordano 2016 | SUP vs CBT | 5.20 | 3.62 | Probably high | 6 |

TE is the mean difference (intervention − comparator) on a 0–100 scale (higher is better). One study per comparison; no pairwise or network meta-analysis was feasible. Estimates were interpreted descriptively.

### Incoherence

#### Table S21. Global incoherence (design‑by‑treatment) and component tests

| Component (fixed-effect model) | Q | df | p‑value |
| --- | --- | --- | --- |
| Total inconsistency | 252.9 | 77 | < 0.0001 |
| Within‑design | 184.4 | 51 | < 0.0001 |
| Between‑design | 68.4 | 26 | < 0.0001 |
| Modelling heterogeneity | 25.61 | 26 | 0.48 |

DBT = design‑by‑treatment global incoherence test; component tests decompose total incoherence into within-design and between-design contributions (fixed‑effect model)

Direct pairwise meta‑analyses showed high heterogeneity (overall I² = 69.9%), the fixed‑effect DBT likely over‑detects inconsistency that largely arises from between‑study variability rather than true disagreement between direct and indirect evidence.

#### Table S22. Global incoherence (Q statistics-design by treatment) and component tests

| Component (fixed effect model) | Q | df | p‑value |
| --- | --- | --- | --- |
| Total inconsistency | 207.3 | 40 | < 0.0001 |
| Within‑design | 150.3 | 25 | < 0.0001 |
| Between‑design | 57.0 | 15 | < 0.0001 |
| Modelling heterogeneity | 10.04 | 15 | 0.817 |

Fixed‑effect values reflect total, within‑design, and between‑design inconsistency under a design‑by‑treatment model.

Direct pairwise meta‑analyses showed high heterogeneity (overall I² = 69.9%), the fixed‑effect DBT likely over‑detects inconsistency that largely arises from between‑study variability rather than true disagreement between direct and indirect evidence.

### Subgroup analysis

### Appendix S2: Iceman assessment

Instrument to assess the Credibility of Effect Modification Analyses (ICEMAN)

in a meta-analysis of randomised controlled trials

Version 1.1

Consider the following important instructions informed by common misapplications of ICEMAN in studies using the instrument

Complete a separate credibility assessment per each effect modifier (e.g., age, comorbidity, drug dose, etc.), outcome (e.g., mortality, stroke, duration of hospital stay), time-point (e.g., 3 months, 6 months), and effect measure (e.g. relative risk, risk difference).

Do not apply ICEMAN if the interaction p-value is 0.1 or larger, i.e., provides very little statistical support for the existence of an effect modification (ICEMAN is designed to address the possible claim of an effect modification rather than the claim of no effect modification).

Response options on the left indicate definitely or probably reduced credibility, response options on the right probably or definitely increased credibility

Completely unclear should be interpreted as probably reduced credibility.

To ensure transparency, provide a supporting comment under each question that provides a rationale for the rating.

To ensure transparency, provide a copy of the completed ICEMAN instrument in the supplement of your article.

| CREDIBILITY ASSESSMENT | | | |
| --- | --- | --- | --- |
| Essential preliminary considerations to define the possible effect modification of interest | | | |
| State a single candidate effect modifier (e.g., age or comorbidity): WHO Geographical regions | | | |
| Was the effect modifier measured before or at randomisation? [ ] yes, continue [ x ] no, stop here and refer to manual for further instructions | | | |
| State a single outcome and time-point (e.g., mortality at 1 year follow-up): Retention in HIV care | | | |
| State a single effect measure (e.g., relative risk or risk difference): Odds ratio | | | |
| 1: Is the analysis of effect modification based on comparison within rather than between trials? | | | |
| [ x ] Completely between | [ ] Mostly between or unclear | [ ] Mostly within | [ ] Completely within |
| Subgroup analysis or meta-regression comparing overall effects of each individual trial. This is typical for aggregate data meta-analysis. | Subgroup analysis or meta-regression with most information coming from overall effects, but some trials providing within-trial subgroup information | Most trials providing within-trial subgroup information; or individual participant data analysis that combines within and between trial information | All trials providing within-trial subgroup information or individual participant data; and the analysis separates within from between trial information, e.g., meta-analysis of interactions |
| Comment: Some trials are conducted in two or more countries, but the countries were in the same geographical regions | | | |
| 2: For within-trial comparisons, is the effect modification similar from trial to trial? [ ] Not applicable: no or one within-RCT comparison | | | |
| [ ] Definitely not similar | [ ] Probably not similar or unclear | [ ] Mostly similar | [ ] Definitely similar |
| Effect modification reported for two or more trials and clearly different directions | Effect modification not reported for individual trials or too imprecise to tell | Effect modification reported for two or more trials, mostly similar in direction, but considerable differences in magnitude | Effect modification reported for two or more trials, similar in direction, only some differences in magnitude |
| Comment: NA | | | |
| 3: For between-trial comparisons, is the number of trials large? [ ] Not applicable: no between RCT comparison | | | |
| [ ] Very small | [ ] Rather small or unclear | [ ] Rather large | [ x ] Large |
| 1 or 2 or in smallest subgroup; 5 or less in continuous meta-regression | 3-4 in smallest subgroup; 6-10 in continuous meta-regression | 5-9 in smallest subgroup; 11 to 15 in continuous meta-regression | 10 or more in smallest subgroup; more than 15 in continuous meta-regression |
| Comment: 84 trials contributed to the network; we only included the direct comparisons for the subgroup analysis | | | |
| 4: Was the direction of effect modification correctly hypothesized a priori? | | | |
| [ ] Definitely no | [ ] Probably no or unclear | [ ] Probably yes | [ x ] Definitely yes |
| Clearly post-hoc or results inconsistent with hypothesized direction or biologically very implausible | Vague hypothesis or hypothesized direction unclear | No prior protocol available but unequivocal statement of a priori hypothesis with correct direction of effect modification | Prior protocol available and includes correct specification of direction of effect modification, e.g., based on a biologic rationale |
| Comment: In terms of geographical regions and income-level classifications,^100, 101^ we anticipated smaller effects in lower‑income countries and in some WHO regions compared with high‑income settings, reflecting differences in health‑system capacity and contextual constraints.98,102We further hypothesized that counselling-based behavioural and educational interventions would show larger effects in high-income settings, where trained personnel are more readily available, whereas supporter-, mHealth-, and reminder-based strategies would demonstrate greater effects in low- and middle-income settings.^102^ | | | |
| 5: Does a test for interaction suggest that chance is an unlikely explanation of the apparent effect modification? (consider irrespective of number of effect modifiers) | | | |
| [ ] Chance a very likely explanation | [ x ] Chance a likely explanation or unclear | [ ] Chance may not explain | [ ] Chance an unlikely explanation |
| Interaction or meta-regression p-value >0.05 | Interaction or meta-regression p-value ≤0.05 and >0.01, or no test of interaction reported and not computable | Interaction or meta-regression p-value ≤0.01 and >0.005 | Interaction or meta-regression p-value ≤0.005 |
| Comment: The p-value is 0.031 from the pairwise subgroup analysis | | | |
| 6: Did the authors test only a small number of effect modifiers or consider the number in their statistical analysis? | | | |
| [ ] Definitely no | [ x ] Probably no or unclear | [ ] Probably yes | [ ] Definitely yes |
| Explicitly exploratory analysis or large number of effect modifiers tested (e.g., greater than 10) and multiplicity not considered in analysis | No mention of number or 4-10 effect modifiers tested and number not considered in analysis | No protocol available but unequivocal statement of 3 or fewer effect modifiers tested | Protocol available and 3 or fewer effect modifiers tested or number considered in analysis |
| Comment: There were nine preplanned subgroup analysis | | | |
| 7: Did the authors use a random effects model? | | | |
| [ x ] Definitely no | [ ] Probably no or unclear | [ ] Probably yes | [ ] Definitely yes |
| Fixed (or common) effect or fixed effects model explicitly stated | Probably fixed effect(s) model | Probably random (or mixed) effects | Random (or mixed) effects explicitly stated |
| Comment: Due to the sparsity of data, all analysis were conducting in fixed-effect models | | | |
| 8: If the effect modifier is a continuous variable, were arbitrary cut points avoided? [ ] not applicable: not continuous | | | |
| [ ] Definitely no | [ ] Probably no or unclear | [ ] Probably yes | [ ] Definitely yes |
| Analysis based on exploratory cut point(s), e.g., picking cut point associated with highest interaction p-value | Analysis based on cut point(s) of unclear origin | Analysis based on pre-specified cut point(s), e.g., suggested by prior RCT | Analysis based on the full continuum, e.g., assuming a linear or logarithmic relationship |
| Comment: NA; dichotomous outcome assessment | | | |
| 9 Optional: Are there any additional considerations that may increase or decrease credibility? (manual section 3.9) [ ] not applicable | | | |
|  | [ ] Yes, probably decrease | [ x ] Yes, probably increase | |
| Comment: Likely increase due to high attrition rates 10: How would you rate the overall credibility of the proposed effect modification? The overall rating should be driven by the items that decrease credibility. The following provides a sensible strategy: All responses definitely or probably decrease credibility or unclear 🡪 very low Two or more responses definitely decrease credibility 🡪 maximum usually low even if all other responses satisfy credibility criteria One response definitely decreases credibility 🡪 maximum usually moderate even if all other responses satisfy credibility criteria Two responses probably decrease credibility 🡪 maximum usually moderate even if all other responses satisfy credibility criteria No response options definitely or probably decrease credibility 🡪 high very likely Place a mark on the continuous line (or type “x” in editable version) \| \| \| \| \|; \| x \| \| \| \|; \| Very low credibility \| Low credibility \| Moderate credibility \| High credibility \|; \| Minimal to no support for effect modification; Use overall effect for each subgroup \| Some but insufficient support for effect modification; Use overall effect for each subgroup but note remaining uncertainty \| Likely effect modification; Use separate effects for each subgroup but note remaining uncertainty \| Very likely effect modification; Use separate effects for each subgroup \|; Comment: \| \| \| \| \| | | | |

#### Table S23. Iceman assessment for subgroups

| Subgroups | Comparison | No. of trials between trial | Direction prior hypothesis | Interaction test | Small number of effect modifier | Model | Effect modifier continuous | Overall |
| --- | --- | --- | --- | --- | --- | --- | --- | --- |
| Geographical regions | Completely between | Large | Probably yes | Chance a likely explanation or unclear | Probably no or unclear | Definitely no | Definitely no | Low |
| Income regions | Completely between | Large | Probably yes | Chance an unlikely explanation | Probably no or unclear | Definitely no | Definitely no | Low |
| Follow-up time | Completely between | Large | Probably yes | Chance a likely explanation or unclear | Probably no or unclear | Definitely no | Definitely no | Low |
| RoB | Completely between | Large | Probably yes | Chance an unlikely explanation | Probably no or unclear | Definitely no | Definitely no | Very low |
| Design | Completely between | Large | Probably yes | Chance a very likely explanation | Probably no or unclear | Definitely no | Definitely no | Very low |
| Pragmatism | Completely between | Large | Probably yes | Chance an unlikely explanation | Probably no or unclear | Definitely no | Definitely no | Low |
| Intensity of intervention | Completely between | Large | Probably yes | Chance a very likely explanation | Probably no or unclear | Definitely no | Definitely no | Very low |

ICEMAN = Instrument for Assessing the Credibility of Effect Modification Analyses. Ratings were based on eight prespecified domains assessing the robustness of subgroup findings^103^

All subgroup analyses were rated as low/very low credibility; chance remains a likely explanation for all apparent subgroup differences

### Subgroup analysis for retention in HIV care

#### Table S24. Risk of bias: Subgroup analysis of retention in HIV care

| Comparison | Risk of bias | k | OR (95% CI) |
| --- | --- | --- | --- |
| BC vs CM | Definitely high | 1 | 3.17 (0.92 to 10.90) |
| BC vs DSD | Definitely high | 1 | 1.18 (0.29 to 4.75) |
| BC vs EDU | Definitely low | 2 | 1.12 (0.91 to 1.39) |
| BC vs EDU | Probably low | 1 | 1.65 (0.94 to 2.89) |
| BC vs EI | Probably low | 1 | 1.50 (1.06 to 2.14) |
| BC vs eSoC | Probably low | 1 | 2.42 (1.19 to 4.91) |
| BC vs REM | Definitely low | 1 | 1.11 (0.89 to 1.39) |
| BC vs SoC | Definitely low | 1 | 0.74 (0.47 to 1.16) |
| BC vs SoC | Probably high | 5 | 1.14 (0.95 to 1.36) |
| BC vs SoC | Probably low | 2 | 2.19 (1.65 to 2.90) |
| BC vs SUP | Probably high | 1 | 0.69 (0.42 to 1.12) |
| BC vs SUP | Probably low | 1 | 1.71 (1.26 to 2.31) |
| BC vs TS | Probably low | 1 | 0.47 (0.22 to 0.97) |
| DSD vs eSoC | Probably high | 1 | 2.88 (1.14 to 7.23) |
| DSD vs SoC | Definitely low | 3 | 1.85 (1.44 to 2.36) |
| DSD vs SoC | Probably high | 2 | 1.11 (0.71 to 1.72) |
| DSD vs SoC | Probably low | 3 | 1.23 (0.78 to 1.95) |
| DSD vs SUP | Definitely low | 1 | 1.17 (0.79 to 1.71) |
| DSD vs SUP | Probably high | 1 | 0.67 (0.44 to 1.02) |
| EDU vs EI | Probably high | 1 | 0.76 (0.38 to 1.49) |
| EDU vs eSoC | Probably low | 1 | 1.46 (0.65 to 3.25) |
| EDU vs REM | Definitely low | 2 | 0.99 (0.81 to 1.21) |
| EDU vs REM | Probably high | 1 | 0.67 (0.34 to 1.33) |
| EDU vs SoC | Definitely low | 1 | 0.66 (0.43 to 1.01) |
| EDU vs SoC | Probably low | 1 | 1.32 (0.79 to 2.21) |
| EI vs eSoC | Definitely high | 1 | 0.91 (0.40 to 2.08) |
| EI vs eSoC | Definitely low | 1 | 2.18 (1.43 to 3.33) |
| EI vs mHealth | Probably high | 1 | 0.89 (0.64 to 1.24) |
| EI vs SoC | Definitely low | 2 | 2.48 (1.44 to 4.07) |
| EI vs SoC | Probably high | 2 | 0.99 (0.74 to 1.32) |
| EI vs SoC | Probably low | 4 | 1.45 (1.17 to 1.80) |
| eSoC vs SUP | Definitely low | 1 | 0.70 (0.39 to 1.26) |
| mHealth vs REM | Definitely low | 1 | 1.70 (1.11 to 2.59) |
| mHealth vsREM | Probably high | 1 | 0.99 (0.76 to 1.28) |
| mHealth vsSoC | Definitely high | 1 | 0.28 (0.08 to 0.92) |
| mHealth vsSoC | Definitely low | 3 | 1.13 (0.92 to 1.39) |
| mHealth vsSoC | Probably high | 5 | 1.11 (0.90 to 1.35) |
| mHealth vs SoC | Probably low | 5 | 0.72 (0.54 to 0.96) |
| MM vs SoC | Probably high | 1 | 0.70 (0.54 to 0.90) |
| MM vs SoC | Probably low | 1 | 0.62 (0.27 to 1.41) |
| REM vs SoC | Definitely high | 1 | 0.51 (0.17 to 1.49) |
| REM vs SoC | Probably high | 5 | 1.12 (0.93 to 1.34) |
| REM vs SoC | Probably low | 3 | 2.85 (1.60 to 5.09) |
| SoC vs SUP | Definitely low | 3 | 0.63 (0.46 to 0.86) |
| SoC vs SUP | Probably high | 1 | 0.60 (0.38 to 0.95) |
| SoC vs SUP | Probably low | 4 | 0.77 (0.58 to 1.03) |
| SoC vs TS | Definitely low | 2 | 0.74 (0.47 to 1.16) |
| SUP vs TS | Probably high | 1 | 0.52 (0.30 to 0.90) |

Due to k ≤ 1 in many subgroups, estimates were interpreted descriptively. Risk of bias assessed with RoBUST-RCT^104^

#### Table S25. Follow-up time (< 12 vs 12 mos): Subgroup analysis of retention in HIV care

| Comparison | Follow-up time (mos) | k | OR (95 % CI) |
| --- | --- | --- | --- |
| BC vs CM | < 12 | 1 | 3.17 (0.93 to 10.90) |
| BC vs DSD | < 12 | 1 | 0.96 (0.52 to 1.76) |
| BC vs EDU | < 12 | 1 | 1.43 (1.05 to 1.94) |
| BC vs EDU | 12 | 2 | 1.07 (0.85 to 1.34) |
| BC vs EI | < 12 | 1 | 0.98 (0.73 to 1.32) |
| BC vs eSoC | < 12 | 1 | 2.32 (1.16 to 4.65) |
| BC vs REM | 12 | 1 | 1.35 (1.09 to 1.65) |
| BC vs SoC | < 12 | 3 | 1.26 (1.00 to 1.60) |
| BC vs SoC | 12 | 5 | 1.37 (1.16 to 1.62) |
| BC vs SUP | < 12 | 2 | 0.87 (0.64 to 1.19) |
| CM vs SoC | 12 | 2 | 1.72 (1.11 to 2.67) |
| DSD vs eSoC | 12 | 1 | 2.02 (1.31 to 3.12) |
| DSD vs SoC | < 12 | 1 | 1.32 (0.72 to 2.40) |
| DSD vs SoC | > 12 | 1 | 1.08 (0.48 to 2.46) |
| DSD vs SoC | 12 | 6 | 1.48 (1.21 to 1.81) |
| DSD vs SUP | < 12 | 1 | 0.91 (0.49 to 1.68) |
| DSD vs SUP | 12 | 1 | 1.32 (1.00 to 1.73) |
| EDU vs EI | < 12 | 1 | 0.69 (0.50 to 0.94) |
| EDU vs eSoC | < 12 | 1 | 1.63 (0.79 to 3.36) |
| EDU vs REM | < 12 | 2 | 0.68 (0.52 to 0.89) |
| EDU vs REM | 12 | 1 | 1.26 (1.00 to 1.59) |
| EDU vs SoC | < 12 | 1 | 0.89 (0.68 to 1.16) |
| EDU vs SoC | 12 | 1 | 1.28 (1.00 to 1.66) |
| EI vs eSoC | 12 | 2 | 1.87 (1.32 to 2.65) |
| EI vs mHealth | < 12 | 1 | 1.37 (1.03 to 1.82) |
| EI vs SoC | < 12 | 4 | 1.29 (1.07 to 1.56) |
| EI vs SoC | 12 | 4 | 1.37 (1.00 to 1.87) |
| eSoC vs SUP | 12 | 1 | 0.65 (0.42 to 1.01) |
| mHealth vs REM | 12 | 2 | 1.07 (0.83 to 1.37) |
| mHealth vs SoC | < 12 | 6 | 0.94 (0.75 to 1.19) |
| mHealth vs SoC | 12 | 8 | 1.09 (0.94 to 1.26) |
| MM vs SoC | < 12 | 1 | 0.62 (0.27 to 1.42) |
| MM vs SoC | 12 | 1 | 0.70 (0.55 to 0.90) |
| REM vs SoC | < 12 | 6 | 1.30 (1.09 to 1.56) |
| REM vs SoC | > 12 | 1 | 0.51 (0.18 to 1.50) |
| REM vs SoC | 12 | 2 | 1.02 (0.81 to 1.28) |
| SoC vs SUP | < 12 | 2 | 0.69 (0.49 to 0.96) |
| SoC vs SUP | > 12 | 1 | 0.54 (0.31 to 0.92) |
| SoC vs SUP | 12 | 5 | 0.89 (0.71 to 1.11) |

Due to k ≤ 1 in many subgroups, estimates were interpreted descriptively. Follow‑up was grouped as <12 or >12 months versus 12 months, the duration used for the original pooling.

#### Table S26. Country income level: Subgroup analysis for retention in HIV

| Comparisons | Country income-level | k | OR (95% CI) |
| --- | --- | --- | --- |
| BC vs DSD | HIC | 1 | 0.98 (0.53 to 1.79) |
| BC vs EDU | HIC | 2 | 0.95 (0.75 to 1.20) |
| BC vs EDU | UMIC | 1 | 1.51 (1.03 to 2.22) |
| BC vs EI | HIC | 1 | 0.79 (0.59 to 1.05) |
| BC vs eSoC | HIC | 1 | 2.07 (1.04 to 4.14) |
| BC vs REM | HIC | 1 | 1.26 (1.00 to 1.58) |
| BC vs SoC | HIC | 4 | 1.09 (0.89 to 1.32) |
| BC vs SoC | LMIC | 2 | 2.74 (1.93 to 3.89) |
| BC vs SoC | UMIC | 2 | 1.30 (1.00 to 1.68) |
| BC vs SUP | HIC | 1 | 1.09 (0.81 to 1.48) |
| BC vs SUP | LMIC | 1 | 1.22 (0.69 to 2.16) |
| BC vs TS | HIC | 1 | 0.74 (0.46 to 1.21) |
| DSD vs DSD+MMD | LMIC | 2 | 1.13 (0.63 to 2.03) |
| DSD vs DSD+MMD | UMIC | 1 | 1.10 (0.61 to 1.98) |
| DSD vs eSoC | LIC | 1 | 3.84 (1.89 to 7.78) |
| DSD vs SoC | HIC | 1 | 1.11 (0.61 to 2.03) |
| DSD vs SoC | LIC | 1 | 1.23 (0.72 to 2.12) |
| DSD vs SoC | LMIC | 4 | 0.85 (0.54 to 1.34) |
| DSD vs SoC | UMIC | 2 | 1.84 (1.44 to 2.35) |
| DSD vs SUP | HIC | 1 | 1.12 (0.61 to 2.06) |
| DSD vs SUP | UMIC | 1 | 1.12 (0.79 to 1.58) |
| DSD+MMD vs SoC | LMIC | 2 | 0.76 (0.44 to 1.30) |
| EDU vs EI | HIC | 1 | 0.83 (0.59 to 1.18) |
| EDU vs eSoC | HIC | 1 | 2.18 (1.07 to 4.44) |
| EDU vs REM | HIC | 1 | 1.32 (1.04 to 1.68) |
| EDU vs REM | LIC | 1 | 0.59 (0.41 to 0.85) |
| EDU vs REM | UMIC | 1 | 0.76 (0.50 to 1.17) |
| EDU vs SoC | LIC | 1 | 0.83 (0.50 to 1.39) |
| EDU vs SoC | UMIC | 1 | 0.86 (0.58 to 1.27) |
| EI vs eSoC | LMIC | 2 | 1.42 (0.95 to 2.11) |
| EI vs mHealth | UMIC | 1 | 0.71 (0.47 to 1.09) |
| EI vs SoC | HIC | 3 | 1.38 (1.10 to 1.72) |
| EI vs SoC | LIC | 2 | 2.11 (1.43 to 3.12) |
| EI vs SoC | LMIC | 1 | 2.01 (1.06 to 3.80) |
| EI vs SoC | UMIC | 2 | 0.97 (0.68 to 1.39) |
| eSoC vs SUP | LIC | 1 | 0.33 (0.17 to 0.65) |
| mHealth vs REM | LMIC | 1 | 0.62 (0.41 to 0.93) |
| mHealth vs REM | UMIC | 1 | 1.22 (0.86 to 1.72) |
| mHealth vs SoC | HIC | 5 | 0.90 (0.72 to 1.12) |
| mHealth vs SoC | LIC | 3 | 1.32 (1.00 to 1.73) |
| mHealth vs SoC | LMIC | 5 | 0.93 (0.74 to 1.17) |
| mHealth vs SoC | UMIC | 1 | 1.37 (0.99 to 1.88) |
| MM vs SoC | HIC | 1 | 0.70 (0.55 to 0.90) |
| MM vs SoC | LIC | 1 | 0.62 (0.27 to 1.42) |
| REM vs SoC | HIC | 3 | 0.87 (0.65 to 1.15) |
| REM vs SoC | LIC | 1 | 1.41 (0.79 to 2.51) |
| REM vs SoC | LMIC | 2 | 1.50 (1.02 to 2.22) |
| REM vs SoC | UMIC | 3 | 1.12 (0.92 to 1.36) |
| SoC vs SUP | HIC | 2 | 1.00 (0.74 to 1.37) |
| SoC vs SUP | LIC | 2 | 1.02 (0.70 to 1.50) |
| SoC vs SUP | LMIC | 2 | 0.44 (0.27 to 0.72) |
| SoC vs SUP | UMIC | 2 | 0.61 (0.44 to 0.84) |
| SoC vs TS | HIC | 1 | 0.68 (0.43 to 1.09) |
| SoC vs TS | UMIC | 1 | 0.37 (0.23 to 0.59) |
| SUP vs TS | UMIC | 1 | 0.61 (0.39 to 0.94) |
| Country income level was classified using the World Bank income groups.^101^ | | | |

| Comparisons | Geographical regions | k | OR (95% CI) |
| --- | --- | --- | --- |
| BC vs CM | AMRO | 1 | 1.37 (0.69 to 2.70) |
| BC vs DSD | AMRO | 1 | 0.97 (0.53 to 1.77) |
| BC vs EDU | AMRO | 2 | 0.97 (0.76 to 1.23) |
| BC vs EDU | EURO | 1 | 1.29 (0.79 to 2.10) |
| BC vs EI | AMRO | 1 | 0.76 (0.57 to 1.01) |
| BC vs eSoC | AMRO | 1 | 1.60 (0.71 to 3.61) |
| BC vs REM | AMRO | 1 | 1.27 (1.00 to 1.60) |
| BC vs SoC | AFRO | 3 | 1.49 (1.17 to 1.89) |
| BC vs SoC | AMRO | 4 | 1.07 (0.88 to 1.30) |
| BC vs SoC | EURO | 1 | 3.79 (2.30 to 6.24) |
| BC vs SUP | AFRO | 1 | 1.03 (0.75 to 1.42) |
| BC vs SUP | AMRO | 1 | 1.08 (0.80 to 1.47) |
| BC vs TS | AMRO | 1 | 0.85 (0.52 to 1.40) |
| CM vs SoC | AMRO | 1 | 0.78 (0.40 to 1.53) |
| CM vs SoC | EURO | 1 | 1.96 (1.15 to 3.34) |
| DSD vs eSoC | AFRO | 1 | 2.33 (1.52 to 3.58) |
| DSD vs SoC | AFRO | 7 | 1.50 (1.24 to 1.83) |
| DSD vs SoC | AMRO | 1 | 1.11 (0.61 to 2.01) |
| DSD vs SUP | AFRO | 1 | 1.04 (0.80 to 1.36) |
| DSD vs SUP | AMRO | 1 | 1.12 (0.61 to 2.07) |
| EDU vs EI | AMRO | 1 | 0.79 (0.56 to 1.12) |
| EDU vs eSoC | EURO | 1 | 4.24 (1.15 to 15.61) |
| EDU vs REM | AMRO | 1 | 1.31 (1.03 to 1.66) |
| EDU vs REM | SEARO | 1 | 0.52 (0.36 to 0.76) |
| EDU vs REM | WPRO | 1 | 0.61 (0.23 to 1.64) |
| EDU vs SoC | AFRO | 2 | 0.91 (0.60 to 1.40) |
| EI vs eSoC | AFRO | 1 | 2.01 (1.38 to 2.94) |
| EI vs eSoC | SEARO | 1 | 0.91 (0.40 to 2.08) |
| EI vs mHealth | AFRO | 1 | 1.14 (0.87 to 1.50) |
| EI vs SoC | AFRO | 4 | 1.30 (1.02 to 1.65) |
| EI vs SoC | AMRO | 4 | 1.40 (1.13 to 1.74) |
| eSoC vs SUP | AFRO | 1 | 0.45 (0.29 to 0.69) |
| mHealth vs REM | AFRO | 2 | 0.99 (0.80 to 1.22) |
| mHealth vs SoC | AFRO | 9 | 1.14 (0.98 to 1.32) |
| mHealth vs SoC | AMRO | 5 | 0.90 (0.72 to 1.12) |
| MM vs SoC | AMRO | 1 | 0.62 (0.27 to 1.42) |
| MM vs SoC | EURO | 1 | 0.70 (0.55 to 0.90) |
| REM vs SoC | AFRO | 6 | 1.15 (0.97 to 1.37) |
| REM vs SoC | AMRO | 3 | 0.84 (0.63 to 1.13) |
| SoC vs SUP | AFRO | 6 | 0.69 (0.56 to 0.86) |
| SoC vs SUP | AMRO | 2 | 1.01 (0.74 to 1.38) |
| SoC vs TS | AFRO | 1 | 0.40 (0.26 to 0.62) |
| SoC vs TS | AMRO | 1 | 0.80 (0.49 to 1.29) |
| SUP vs TS | AFRO | 1 | 0.58 (0.38 to 0.89) |

Country income level was classified using the World Bank income groups.^101^ Several subgroups had k ≤ 1, so estimates were interpreted descriptively.

#### Table S27. Population groups (general vs key populations). Subgroup analysis for retention in HIV

| Comparisons | Population | k | OR (95% CI) |
| --- | --- | --- | --- |
| BC vs CM | Key | 1 | 0.88 (0.52 to 1.48) |
| BC vs DSD | Key | 1 | 0.75 (0.50 to 1.11) |
| BC vs EDU | General | 2 | 1.22 (1.01 to 1.48) |
| BC vs EDU | Key | 1 | 1.12 (0.65 to 1.92) |
| BC vs eSoC | Key | 1 | 2.17 (1.07 to 4.39) |
| BC vs REM | General | 1 | 1.16 (0.98 to 1.38) |
| BC vs SoC | General | 7 | 1.44 (1.23 to 1.69) |
| BC vs SoC | Key | 1 | 1.34 (1.07 to 1.69) |
| BC vs SUP | General | 1 | 1.29 (0.99 to 1.70) |
| BC vs SUP | Key | 1 | 0.84 (0.63 to 1.13) |
| CM vs SoC | General | 1 | 1.35 (0.63 to 2.86) |
| CM vs SoC | Key | 1 | 1.53 (0.94 to 2.51) |
| DSD vs eSoC | General | 1 | 1.88 (1.26 to 2.80) |
| DSD vs SoC | General | 5 | 1.33 (1.07 to 1.65) |
| DSD vs SoC | Key | 3 | 1.79 (1.28 to 2.53) |
| DSD vs SUP | General | 1 | 1.19 (0.90 to 1.57) |
| DSD vs SUP | Key | 1 | 1.12 (0.74 to 1.70) |
| EDU vs eSoC | Key | 1 | 1.95 (0.88 to 4.30) |
| EDU vs REM | General | 3 | 0.95 (0.79 to 1.14) |
| EDU vs SoC | General | 1 | 1.18 (0.97 to 1.45) |
| EDU vs SoC | Key | 1 | 1.20 (0.72 to 2.01) |
| eSoC vs SUP | General | 1 | 0.63 (0.43 to 0.94) |
| mHealth vs REM | Key | 2 | 1.20 (0.84 to 1.71) |
| mHealth vs SoC | General | 8 | 1.12 (0.96 to 1.30) |
| mHealth vs SoC | Key | 6 | 0.81 (0.64 to 1.04) |
| REM vs SoC | General | 7 | 1.25 (1.07 to 1.46) |
| REM vs SoC | Key | 2 | 0.68 (0.46 to 0.99) |
| SoC vs SUP | General | 5 | 0.90 (0.71 to 1.12) |
| SoC vs SUP | Key | 3 | 0.63 (0.47 to 0.83) |

General versus key population classification followed study reporting.^99^ Estimates were descriptive because many subgroups had k ≤ 1.

#### Table S28. WHO geographical regions: Subgroup analysis for retention in HIV

| Comparisons | Regions | k | OR (95% CI) |
| --- | --- | --- | --- |
| BC vs CM | AMRO | 1 | 1.37 (0.69 to 2.70) |
| BC vs DSD | AMRO | 1 | 0.97 (0.53 to 1.77) |
| BC vs EDU | AMRO | 2 | 0.97 (0.76 to 1.23) |
| BC vs EDU | EURO | 1 | 1.29 (0.79 to 2.10) |
| BC vs EI | AMRO | 1 | 0.76 (0.57 to 1.01) |
| BC vs eSoC | AMRO | 1 | 1.60 (0.71 to 3.61) |
| BC vs REM | AMRO | 1 | 1.27 (1.00 to 1.60) |
| BC vs SoC | AFRO | 3 | 1.49 (1.17 to 1.89) |
| BC vs SoC | AMRO | 4 | 1.07 (0.88 to 1.30) |
| BC vs SoC | EURO | 1 | 3.79 (2.30 to 6.24) |
| BC vs SUP | AFRO | 1 | 1.03 (0.75 to 1.42) |
| BC vs SUP | AMRO | 1 | 1.08 (0.80 to 1.47) |
| BC:TS | AMRO | 1 | 0.85 (0.52 to 1.40) |
| CM vs SoC | AMRO | 1 | 0.78 (0.40 to 1.53) |
| CM vs SoC | EURO | 1 | 1.96 (1.15 to 3.34) |
| DSD vs eSoC | AFRO | 1 | 2.33 (1.52 to 3.58) |
| DSD vs SoC | AFRO | 7 | 1.50 (1.24 to 1.83) |
| DSD vs SoC | AMRO | 1 | 1.11 (0.61 to 2.01) |
| DSD:SUP | AFRO | 1 | 1.04 (0.80 to 1.36) |
| DSD:SUP | AMRO | 1 | 1.12 (0.61 to 2.07) |
| EDU vs EI | AMRO | 1 | 0.79 (0.56 to 1.12) |
| EDU vs eSoC | EURO | 1 | 4.24 (1.15 to 15.61) |
| EDU vs REM | AMRO | 1 | 1.31 (1.03 to 1.66) |
| EDU vs REM | SEARO | 1 | 0.52 (0.36 to 0.76) |
| EDU vs REM | WPRO | 1 | 0.61 (0.23 to 1.64) |
| EDU vs SoC | AFRO | 2 | 0.91 (0.60 to 1.40) |
| EI vs eSoC | AFRO | 1 | 2.01 (1.38 to 2.94) |
| EI vs eSoC | SEARO | 1 | 0.91 (0.40 to 2.08) |
| EI vs mHealth | AFRO | 1 | 1.14 (0.87 to 1.50) |
| EI vs SoC | AFRO | 4 | 1.30 (1.02 to 1.65) |
| EI vs SoC | AMRO | 4 | 1.40 (1.13 to 1.74) |
| eSoC vs SUP | AFRO | 1 | 0.45 (0.29 to 0.69) |
| mHealth vs REM | AFRO | 2 | 0.99 (0.80 to1.22) |
| mHealth vs SoC | AFRO | 9 | 1.14 (0.98 to 1.32) |
| mHealth vs SoC | AMRO | 5 | 0.90 (0.72 to1.12) |
| MM vs SoC | AMRO | 1 | 0.62 (0.27 to 1.42) |
| MM vs SoC | EURO | 1 | 0.70 (0.55 to 0.90) |
| REM vs SoC | AFRO | 6 | 1.15 (0.97 to 1.37) |
| REM vs SoC | AMRO | 3 | 0.84 (0.63 to 1.13) |
| SoC vs SUP | AFRO | 6 | 0.69 (0.56 to 0.86) |
| SoC vs SUP | AMRO | 2 | 1.01 (0.74 to 1.38) |
| SoC vs TS | AFRO | 1 | 0.40 (0.26 to 0.62) |
| SoC vs TS | AMRO | 1 | 0.80 (0.49 to 1.29) |
| SUP vs TS | AFRO | 1 | 0.58 (0.38 to 0.89) |

Subgroup analysis for WHO geographical regions^100^. Estimates were descriptive because many subgroups had k ≤ 1.

#### Table S29. Study design (parallel vs cluster): Subgroup analysis for retention in HIV

| Comparison | Study design | k | OR (95% CI) |
| --- | --- | --- | --- |
| BC vs DSD | Parallel | 1 | 0.80 (0.61 to 1.04) |
| BC vs EDU | Parallel | 2 | 0.98 (0.80 to 1.21) |
| BC vs EDU | Parallel | 1 | 3.91 (1.16 to13.16) |
| BC vs EI | Parallel | 1 | 0.78 (0.26 to 2.37) |
| BC vs eSoC | Parallel | 1 | 2.66 (1.55 to 4.58) |
| BC vs REM | Parallel | 1 | 1.21 (1.02 to 1.43) |
| BC vs SoC | Cluster | 2 | 1.76 (1.19 to 2.60) |
| BC vs SoC | Parallel | 6 | 1.39 (1.21 to 1.60) |
| BC vs SUP | Parallel | 1 | 0.98 (0.78 to 1.24) |
| BC vs SUP | Parallel | 1 | 0.82 (0.29 to 2.29) |
| BC vs TS | Parallel | 1 | 0.80 (0.54 to 1.19) |
| DSD vs DSD+MMD | Cluster | 2 | 1.02 (0.66 to 1.57) |
| DSD vs DSD+MMD | Parallel | 1 | 1.68 (0.41 to 6.95) |
| DSD vs eSoC | Parallel | 1 | 2.88 (1.15 to 7.23) |
| DSD vs SoC | Cluster | 3 | 0.77 (0.52 to 1.14) |
| DSD vs SoC | Parallel | 4 | 1.75 (1.39 to 2.18) |
| DSD vs SoC | Parallel | 1 | 0.81 (0.16 to 4.11) |
| DSD vs SUP | Cluster | 1 | 1.09 (0.71 to 1.66) |
| DSD vs SUP | Parallel | 1 | 1.23 (0.91 to 1.67) |
| DSD+MMD vs SoC | Cluster | 1 | 0.75 (0.46 to 1.22) |
| DSD+MMD vs SoC | Parallel | 1 | 0.48 (0.12 to 1.97) |
| EDU vs EI | Parallel | 1 | 0.57 (0.38 to 0.84) |
| EDU vs eSoC | Parallel | 1 | 2.71 (1.54 to 4.75) |
| EDU vs REM | Parallel | 2 | 1.23 (0.99 to 1.51) |
| EDU vs REM | Parallel | 1 | 0.54 (0.37 to 0.78) |
| EDU vs SoC | Cluster | 2 | 0.91 (0.60 to 1.40) |
| EI vs eSoC | Cluster | 2 | 1.42 (0.95 to 2.11) |
| EI vs mHealth | Cluster | 1 | 0.97 (0.66 to 1.45) |
| EI vs SoC | Cluster | 2 | 1.19 (0.97 to 1.45) |
| EI vs SoC | Parallel | 5 | 2.50 (1.76 to 3.55) |
| EI vs SoC | Parallel | 1 | 1.29 (0.80 to 2.07) |
| eSoC vs SUP | Parallel | 1 | 0.37 (0.21 to 0.64) |
| mHealth vs REM | Cluster | 1 | 1.02 (0.59 to 1.75) |
| mHealth vs REM | Parallel | 1 | 0.93 (0.76 to 1.14) |
| mHealth vs SoC | Cluster | 1 | 1.22 (0.84 to 1.77) |
| mHealth vs SoC | Cluster | 1 | 1.06 (0.62 to 1.81) |
| mHealth vs SoC | Parallel | 10 | 1.08 (0.93 to 1.24) |
| mHealth vs SoC | Parallel | 2 | 0.59 (0.36 to 0.96) |
| MM vs SoC | Parallel | 1 | 0.62 (0.27 to 1.42) |
| MM vs SoC | Parallel | 1 | 0.70 (0.55 to 0.90) |
| REM vs SoC | Cluster | 1 | 1.04 (0.61 to 1.77) |
| REM vs SoC | Parallel | 7 | 1.15 (0.99 to 1.34) |
| REM vs SoC | Parallel | 1 | 0.48 (0.18 to 1.31) |
| SoC vs SUP | Cluster | 1 | 1.42 (0.87 to 2.31) |
| SoC vs SUP | Parallel | 6 | 0.71 (0.57 to 0.88) |
| SoC vs SUP | Parallel | 1 | 0.81 (0.51 to 1.29) |
| SoC vs TS | Parallel | 2 | 0.58 (0.39 to 0.84) |
| SUP vs TS | Cluster | 1 | 0.53 (0.31 to 0.91) |

Estimates were descriptive because many subgroups had k ≤ 1

Table S16 | Retention measures: Subgroup analysis for retention in HIV

| Comparisons | Components | k | OR (95% CI) |
| --- | --- | --- | --- |
| BC vs CM | Clinic visits | 1 | 1.05 (0.42 to 2.63) |
| BC vs DSD | Clinic visits | 1 | 1.19 (0.30 to 4.75) |
| BC vs EDU | Composite | 1 | 2.11 (0.37 to 11.94) |
| BC vs EDU | Kept visit | 2 | 1.01 (0.81 to 1.26) |
| BC vs EI | Lab | 1 | 5.60 (0.54 to 57.95) |
| BC vs eSoC | Scheduled | 1 | 1.60 (0.71 to 3.61) |
| BC vs REM | Kept visit | 1 | 1.28 (1.01 to 1.62) |
| BC vs SoC | Ambiguous | 1 | 0.67 (0.42 to 1.05) |
| BC vs SoC | Clinic visits | 1 | 4.54 (2.80 to 7.35) |
| BC vs SoC | Gap score | 1 | 2.13 (1.49 to 3.03) |
| BC vs SoC | Kept visit | 3 | 0.70 (0.43 to 1.12) |
| BC vs SoC | Pharmacy | 1 | 2.15 (1.27 to 3.62) |
| BC vs SoC | Record | 1 | 1.10 (0.85 to 1.43) |
| BC:SUP | Group | 1 | 0.62 (0.17 to 2.33) |
| BC:SUP | Record | 1 | 1.13 (0.76 to 1.68) |
| BC:TS | Record | 1 | 1.21 (0.70 to 2.11) |
| CM vs SoC | Clinic visits | 1 | 4.30 (1.72 to 10.79) |
| CM vs SoC | Kept visit | 1 | 1.96 (1.15 to 3.34) |
| DSD vs DSD+MMD | Missed visit | 2 | 1.26 (0.80 to 1.98) |
| DSD vs DSD+MMD | Pharmacy | 1 | 1.89 (0.56 to 6.39) |
| DSD vs SoC | Ambiguous | 1 | 0.06 (0.02 to 0.21) |
| DSD vs SoC | Follow up | 1 | 1.20 (0.61 to 2.37) |
| DSD vs SoC | Gap score | 1 | 1.39 (0.77 to 2.52) |
| DSD vs SoC | Group | 1 | 1.73 (1.28 to 2.33) |
| DSD vs SoC | Lab | 1 | 2.98 (1.66 to 5.34) |
| DSD vs SoC | Missed visit | 1 | 1.43 (0.80 to 2.57) |
| DSD vs SoC | Pharmacy | 2 | 1.02 (0.49 to 2.12) |
| DSD vs SUP | Follow up | 1 | 1.12 (0.56 to 2.25) |
| DSD vs SUP | Lab | 1 | 0.94 (0.58 to 1.54) |
| DSD+MMD vs SoC | Missed visit | 1 | 1.14 (0.65 to 2.01) |
| DSD+MMD vs SoC | Pharmacy | 1 | 0.54 (0.16 to 1.82) |
| EDU vs EI | Follow up | 1 | 0.83 (0.34 to 1.99) |
| EDU vs eSoC | Record | 1 | 4.24 (1.15 to 15.61) |
| EDU vs REM | Ambiguous | 1 | 0.61 (0.23 to 1.64) |
| EDU vs REM | Kept visit | 1 | 1.26 (1.00 to 1.60) |
| EDU vs REM | Pharmacy | 1 | 0.52 (0.36 to 0.76) |
| EDU vs SoC | Clinic visits | 1 | 1.11 (0.63 to 1.97) |
| EDU vs SoC | Follow up | 1 | 0.71 (0.37 to 1.36) |
| EI vs eSoC | Gap score | 1 | 0.91 (0.40 to 2.08) |
| EI vs eSoC | Kept visit | 1 | 1.62 (1.03 to 2.56) |
| EI vs mHealth | Pharmacy | 1 | 0.74 (0.46 to 1.17) |
| EI vs SoC | Kept visit | 2 | 2.22 (0.80 to 6.16) |
| EI vs SoC | Lab | 1 | 1.37 (1.08 to 1.73) |
| EI vs SoC | Missed visit | 2 | 3.48 (2.03 to 5.97) |
| EI vs SoC | Pharmacy | 1 | 0.81 (0.55 to 1.20) |
| EI vs SoC | Record | 1 | 2.01 (1.06 to 3.80) |
| EI vs SoC | Scheduled | 1 | 1.40 (0.87 to 2.27) |
| eSoC vs SUP | Ambiguous | 1 | 0.26 (0.11 to 0.60) |
| mHealth vs REM | Clinic visits | 1 | 0.99 (0.74 to 1.33) |
| mHealth vs REM | Scheduled | 1 | 0.87 (0.52 to 1.43) |
| mHealth vs SoC | Ambiguous | 1 | 0.73 (0.36 to 1.51) |
| mHealth vs SoC | Clinic visits | 3 | 1.14 (0.89 to 1.47) |
| mHealth vs SoC | Follow up | 1 | 1.36 (0.46 to 4.03) |
| mHealth vs SoC | Gap score | 2 | 1.16 (0.90 to 1.50) |
| mHealth vs SoC | Group | 1 | 2.56 (1.21 to 5.41) |
| mHealth vs SoC | Kept visit | 2 | 0.93 (0.65 to 1.33) |
| mHealth vs SoC | Missed visit | 1 | 0.61 (0.37 to 1.01) |
| mHealth vs SoC | Pharmacy | 1 | 1.10 (0.75 to 1.61) |
| mHealth vs SoC | Scheduled | 2 | 0.77 (0.53 to 1.12) |
| MM vs SoC | Gap score | 1 | 0.62 (0.27 to 1.42) |
| MM vs SoC | Kept visit | 1 | 0.70 (0.55 to 0.90) |
| REM vs SoC | Clinic visits | 5 | 1.16 (0.95 to 1.42) |
| REM vs SoC | Group | 2 | 2.05 (1.15 to 3.67) |
| REM vs SoC | Missed visit | 1 | 0.61 (0.35 to 1.04) |
| REM vs SoC | Scheduled | 1 | 0.89 (0.54 to 1.47) |
| SoC vs SUP | Follow up | 2 | 0.94 (0.58 to 1.52) |
| SoC vs SUP | Gap score | 2 | 0.95 (0.67 to 1.36) |
| SoC vs SUP | Group | 1 | 0.35 (0.18 to 0.67) |
| SoC vs SUP | Missed visit | 1 | 0.13 (0.02 to 1.12) |
| SoC vs SUP | Pharmacy | 1 | 1.94 (0.94 to 4.02) |
| SoC vs SUP | Scheduled | 1 | 0.54 (0.31 to 0.92) |
| SoC vs TS | Composite | 1 | 0.46 (0.24 to 0.88) |
| SoC vs TS | Record | 1 | 1.10 (0.65 to 1.86) |
| SUP vs TS | Clinic visits | 1 | 0.53 (0.31 to 0.91) |

Retention measures were predefined into components based on a review on retention measures.^105^ Estimates were descriptive because many subgroups had k ≤ 1.

#### Table S30. Pragmatism (effective vs efficacy) : Subgroup analysis for retention in HIV

| Comparison | Subgroup | k | OR (95% CI) |
| --- | --- | --- | --- |
| BC vs DSD | Both | 1 | 1.45 (0.50 to 4.19) |
| BC vs EDU | Both | 2 | 1.05 (0.84 to 1.32) |
| BC vs EDU | Effective | 1 | 1.37 (0.98 to 1.92) |
| BC vs EI | Effective | 1 | 1.03 (0.80 to 1.31) |
| BC vs eSoC | Effective | 1 | 1.92 (1.31 to 2.81) |
| BC vs REM | Both | 1 | 1.04 (0.84 to 1.30) |
| BC vs SoC | Both | 3 | 1.55 (1.23 to 1.94) |
| BC vs SoC | Effective | 5 | 1.32 (1.11 to 1.57) |
| BC vs SUP | Both | 2 | 1.26 (0.97 to 1.65) |
| BC vs TS | Effective | 1 | 1.38 (0.81 to 2.36) |
| CM vs TS | Efficacious | 1 | 0.24 (0.10 to 0.58) |
| DSD vs DSD+MMD | Both | 1 | 1.93 (0.54 to 6.93) |
| DSD vs DSD+MMD | Effective | 2 | 1.29 (0.85 to 1.95) |
| DSD vs eSoC | Effective | 1 | 2.26 (1.54 to 3.32) |
| DSD vs SoC | Both | 1 | 1.07 (0.37 to 3.08) |
| DSD vs SoC | Effective | 7 | 1.55 (1.28 to 1.88) |
| DSD vs SUP | Effective | 2 | 0.93 (0.69 to 1.26) |
| DSD+MMD vs SoC | Both | 1 | 0.55 (0.15 to 1.97) |
| DSD+MMD vs SoC | Effective | 1 | 1.20 (0.78 to 1.85) |
| EDU vs EI | Both | 1 | 0.93 (0.63 to 1.39) |
| EDU vs eSoC | Both | 1 | 1.35 (0.64 to 2.85) |
| EDU vs REM | Both | 2 | 0.99 (0.81 to 1.21) |
| EDU vs REM | Efficacious | 1 | 0.61 (0.23 to 1.64) |
| EDU vs SoC | Effective | 2 | 0.96 (0.69 to 1.33) |
| EI vs eSoC | Both | 1 | 1.45 (0.71 to 2.92) |
| EI vs eSoC | Effective | 1 | 1.87 (1.31 to 2.65) |
| EI vs mHealth | Effective | 1 | 1.20 (0.96 to 1.50) |
| EI vs SoC | Both | 4 | 1.58 (1.15 to 2.17) |
| EI vs SoC | Effective | 4 | 1.28 (1.07 to 1.54) |
| eSoC vs SUP | Effective | 1 | 0.41 (0.27 to 0.63) |
| mHealth vs REM | Effective | 2 | 1.01 (0.82 to 1.24) |
| mHealth vs SoC | Both | 3 | 0.95 (0.71 to 1.26) |
| mHealth vs SoC | Effective | 11 | 1.07 (0.93 to 1.23) |
| MM vs SoC | Effective | 1 | 0.62 (0.27 to 1.42) |
| MM vs SoC | Efficacious | 1 | 0.70 (0.55 to 0.90) |
| REM vs SoC | Both | 3 | 1.48 (1.12 to 1.96) |
| REM vs SoC | Effective | 6 | 1.07 (0.89 to 1.27) |
| SoC vs SUP | Both | 5 | 0.81 (0.64 to 1.03) |
| SoC vs SUP | Effective | 3 | 0.60 (0.44 to 0.80) |
| SoC vs TS | Both | 1 | 0.46 (0.24 to 0.88) |
| SoC vs TS | Effective | 1 | 1.05 (0.62 to 1.77) |
| SUP vs TS | Definitely effective | 1 | 0.53 (0.31 to 0.91) |

Pragmatism was scored using the RITES tool (Rating of Included Trials on the Efficacy–Effectiveness Spectrum). A higher score indicates greater pragmatism (1 = more explanatory, 5 = more pragmatic).^96^Estimates were descriptive because many subgroups had k ≤ 1.

#### Table S31. Intensity of interventions: Subgroup analysis for retention in HIV

| Intervention | Coefficient | p-value |
| --- | --- | --- |
| BC | 0.17 (-0.48 to 0.84) | 0.603 |
| CM | 2.45 (0.30 to 4.60) | 0.025 |
| DSD | 0.88 (-0.13 to .90) | 0.089 |
| DSD.MMD | 0.59 (-0.84 to 2.03) | 0.414 |
| EDU | 0.02 (-0.57 to 0.63) | 0.929 |
| EI | 0.37 (-0.14 to 0.89) | 0.161 |
| eSoC | -0.21 (-1.06 to 0.62) | 0.611 |
| mHealth | -0.10 (-0.49 to 0.29) | 0.619 |
| MM | -0.50 (-2.19 to 1.18) | 0.557 |
| MMD | 0.70 (-0.43 to 1.83) | 0.224 |
| REM | 0.54 (-0.00 to 1.07) | 0.050 |
| SUP | 0.39 (-0.04 to 0.84) | 0.080 |
| TS | 0.55 (-0.57 to 1.68) | 0.337 |
| Intervention | Change in coefficient  (95 % CI) | p-value |
| BC | 0.04 (-0.06 to 0.15) | 0.464 |
| CM | -0.26 (-0.52 to -0.02) | 0.048 |
| DSD | -0.09 (-0.23 to 0.04) | 0.180 |
| EDU | 0.01 (-0.02 to 0.03) | 0.767 |
| EI | 0.01 (-0.01 to 0.01) | 0.625 |
| eSoC | -0.02 (-0.08 to 0.04) | 0.524 |
| mHealth | 0.01 (-0.01 to 0.01) | 0.473 |
| MM | 0.01 (-0.13 to 0.15) | 0.886 |
| REM | -0.01 (-0.01 to -0.01) | 0.028 |
| SUP | -0.01 (-0.0125to 0.01) | 0.1261 |
| TS | 0.01 (-0.14 to 0.16) | 0.9002 |

Positive values indicate larger benefits with higher intensity; negative values indicate attenuation with higher intensity.

Positive values imply OR > 1 after exponentiation; negative values imply OR < 1

### Subgroup analysis for viral load suppression

#### Table S32. Risk of bias: Subgroup analysis for viral load suppression

| Comparisons | Risk of bias | k | OR (95%-CI) |
| --- | --- | --- | --- |
| BC vs EDU | Probably high | 1 | 1.42 (0.71 to 2.81) |
| BC vs eSoC | Probably high | 1 | 5.20 (1.17 to23.05) |
| BC vs SoC | Definitely high | 2 | 1.54 (1.04 to 2.29) |
| BC vs SoC | Probably high | 1 | 0.78 (0.35 to 1.74) |
| BC vs SoC | Probably low | 2 | 1.49 (1.09 to 2.05) |
| BC vs SUP | Probably high | 1 | 0.57 (0.25 to 1.29) |
| BC vs SUP | Probably low | 1 | 0.77 (0.52 to 1.13) |
| DSD vs DSD+MMD | Definitely high | 1 | 2.15 (1.10 to 4.19) |
| DSD vs DSD+MMD | Probably low | 2 | 2.96 (1.45 to 6.02) |
| DSD vs SoC | Definitely high | 2 | 0.76 (0.56 to 1.05) |
| DSD vs SoC | Definitely low | 1 | 0.71 (0.39 to 1.32) |
| DSD vs SoC | Probably high | 2 | 1.51 (1.29 to 1.76) |
| DSD vs SoC | Probably low | 3 | 1.85 (1.14 to 2.98) |
| DSD vs SUP | Definitely low | 1 | 1.00 (0.54 to 1.84) |
| DSD vs SUP | Probably high | 1 | 1.09 (0.83 to 1.45) |
| DSD+MMD vs SoC | Probably low | 2 | 0.62 (0.32 to 1.24) |
| EDU vs EI | Probably high | 1 | 0.41 (0.18 to 0.89) |
| EDU vs REM | Probably low | 1 | 0.90 (0.74 to 1.10) |
| EDU vs SoC | Definitely low | 1 | 1.33 (0.88 to 2.00) |
| EI vs eSoC | Definitely high | 1 | 1.79 (1.06 to 3.01) |
| EI vs mHealth | Definitely high | 1 | 0.44 (0.24 to 0.81) |
| EI vs SoC | Definitely high | 1 | 0.58 (0.33 to 1.01) |
| EI vs SoC | Definitely low | 1 | 2.55 (1.35 to 4.80) |
| EI vs SoC | Probably high | 2 | 1.35 (0.85 to 2.14) |
| mHealth vs REM | Probably high | 1 | 0.84 (0.56 to 1.25) |
| mHealth vs SoC | Definitely high | 2 | 1.30 (0.87 to 1.96) |
| mHealth vs SoC | Definitely low | 3 | 0.99 (0.79 to 1.24) |
| mHealth vs SoC | Probably high | 4 | 0.56 (0.55 to 0.57) |
| mHealth vs SoC | Probably low | 1 | 1.79 (0.90 to 3.58) |
| REM vs SoC | Definitely high | 1 | 1.80 (0.40 to 8.18) |
| REM vs SoC | Probably high | 2 | 0.67 (0.45 to 1.00) |
| REM vs SoC | Probably low | 2 | 1.42 (0.52 to 3.86) |
| SoC vs SUP | Definitely low | 1 | 1.40 (0.76 to 2.58) |
| SoC vs SUP | Probably high | 3 | 0.73 (0.56 to 0.95) |
| SUP vs TS | Definitely high | 1 | 1.00 (0.48 to 2.06) |

Due to k ≤ 1 in many subgroups, estimates were interpreted descriptively. Risk of bias assessed with RoBUST-RCT(100)

#### Table S33. Follow-up time (< 12 vs 12 mos): Subgroup analysis for viral load suppression

| Comparisons | Follow-up time (mos) | k | OR 95%-CI |
| --- | --- | --- | --- |
| BC vs EDU | 12 | 1 | 2.43 (1.54 to 3.85) |
| BC vs eSoC | < 12 | 1 | 5.20 (1.17 to 23.05) |
| BC vs SoC | < 12 | 2 | 1.26 (0.91 to 1.75) |
| BC vs SoC | 12 | 3 | 1.35 (1.01 to 1.81) |
| BC vs SUP | < 12 | 2 | 0.88 (0.64 to 1.21) |
| DSD vs SoC | < 12 | 1 | 1.02 (0.58 to 1.78) |
| DSD vs SoC | > 12 | 1 | 3.05 (1.68 to 5.54) |
| DSD vs SoC | 12 | 6 | 1.30 (1.13 to 1.49) |
| DSD vs SUP | < 12 | 1 | 0.71 (0.41 to 1.24) |
| DSD vs SUP | 12 | 1 | 1.00 (0.75 to 1.35) |
| EDU vs EI | < 12 | 1 | 0.68 (0.42 to 1.11) |
| EDU vs REM | 12 | 1 | 0.93 (0.76 to 1.13) |
| EDU vs SoC | < 12 | 1 | 1.26 (0.86 to 1.84) |
| EI vs eSoC | 12 | 1 | 1.79 (1.06 to 3.01) |
| EI vs mHealth | 12 | 1 | 0.77 (0.45 to 1.33) |
| EI vs SoC | < 12 | 3 | 1.85 (1.29 to 2.65) |
| EI vs SoC | 12 | 1 | 0.43 (0.25 to 0.75) |
| mHealth vs REM | 12 | 1 | 0.94 (0.64 to 1.39) |
| mHealth vs SoC | < 12 | 3 | 1.26 (0.84 to 1.87) |
| mHealth vs SoC | 12 | 7 | 0.56 (0.55 to 0.57) |
| REM vs SoC | < 12 | 3 | 1.86 (0.92 to 3.76) |
| REM vs SoC | > 12 | 1 | 1.80 (0.40 to 8.18) |
| REM vs SoC | 12 | 1 | 0.60 (0.41 to 0.88) |
| SoC vs SUP | < 12 | 2 | 0.70 (0.49 to 0.98) |
| SoC vs SUP | 12 | 2 | 0.78 (0.58 to 1.03) |

Due to k ≤ 1 in many subgroups, estimates were interpreted descriptively. Follow‑up was grouped as <12 or >12 months versus 12 months, the duration used for the original pooling.

#### Table S34. Country income level: Subgroup analysis for viral load suppression

| Comparisons | Country income-level | k | OR (95% CI) |
| --- | --- | --- | --- |
| BC vs EDU | HIC | 1 | 0.89 (0.52 to 1.52) |
| BC vs SoC | HIC | 3 | 0.91 (0.62 to 1.33) |
| BC vs SoC | UMIC | 2 | 1.72 (1.30 to 2.27) |
| BC vs SUP | HIC | 1 | 0.89 (0.63 to 1.26) |
| BC vs SUP | LMIC | 1 | 0.37 (0.08 to 1.66) |
| DSD vs DSD+MMD | LMIC | 2 | 1.89 (0.95 to 3.76) |
| DSD vs DSD+MMD | UMIC | 1 | 2.15 (1.10 to 4.19) |
| DSD vs SoC | HIC | 1 | 0.86 (0.48 to 1.52) |
| DSD vs SoC | LIC | 1 | 1.16 (0.80 to 1.69) |
| DSD vs SoC | LMIC | 4 | 0.86 (0.59 to 1.24) |
| DSD vs SoC | UMIC | 2 | 1.55 (1.33 to 1.82) |
| DSD vs SUP | HIC | 1 | 0.84 (0.47 to 1.48) |
| DSD vs SUP | UMIC | 1 | 0.85 (0.59 to 1.22) |
| DSD+MMD vs SoC | LMIC | 2 | 0.45 (0.23 to 0.89) |
| EDU vs EI | HIC | 1 | 0.67 (0.37 to 1.21) |
| EDU vs REM | HIC | 1 | 0.86 (0.71 to 1.04) |
| EDU vs SoC | UMIC | 1 | 1.33 (0.88 to 2.00) |
| EI vs eSoC | LMIC | 1 | 1.79 (1.06 to 3.01) |
| EI vs mHealth | UMIC | 1 | 0.60 (0.31 to 1.14) |
| EI vs SoC | HIC | 2 | 1.53 (0.98 to 2.38) |
| EI vs SoC | LIC | 1 | 2.55 (1.35 to 4.80) |
| EI vs SoC | UMIC | 1 | 0.49 (0.28 to 0.88) |
| mHealth vs REM | LMIC | 1 | 1.57 (0.98 to 2.50) |
| mHealth vs SoC | HIC | 3 | 0.56 (0.55 to 0.57) |
| mHealth vs SoC | LIC | 3 | 1.39 (1.00 to 1.93) |
| mHealth vs SoC | LMIC | 3 | 1.14 (0.83 to 1.56) |
| mHealth vs SoC | UMIC | 1 | 0.83 (0.49 to 1.40) |
| REM vs SoC | HIC | 2 | 1.19 (0.72 to 1.98) |
| REM vs SoC | LIC | 1 | 1.80 (0.40 to 8.18) |
| REM vs SoC | LMIC | 1 | 0.73 (0.46 to 1.15) |
| REM vs SoC | UMIC | 1 | 0.04 (0.00 to 0.68) |
| SoC vs SUP | HIC | 1 | 0.97 (0.64 to 1.49) |
| SoC vs SUP | LIC | 1 | 1.22 (0.68 to 2.18) |
| SoC vs SUP | LMIC | 1 | 0.74 (0.46 to 1.18) |
| SoC vs SUP | UMIC | 1 | 0.55 (0.38 to 0.79) |

Country income level was classified using the World Bank income groups.^101^ Several subgroups had k ≤ 1, so estimates were interpreted descriptively.

#### Table S35. Population groups (general vs key populations) Subgroup analysis for viral load suppression

| Comparisons | Populations | k | OR (95% CI) |
| --- | --- | --- | --- |
| BC vs EDU | Key | 1 | 1.62 (0.75 to 3.51) |
| BC vs eSoC | Key | 1 | 5.20 (1.17 to 23.05) |
| BC vs SoC | General | 4 | 1.52 (1.17 to 1.96) |
| BC vs SoC | Key | 1 | 0.94 (0.62 to 1.43) |
| BC vs SUP | General | 1 | 1.06 (0.72 to 1.56) |
| BC vs SUP | Key | 1 | 0.78 (0.55 to 1.11) |
| DSD vs SoC | General | 5 | 1.31 (1.13 to 1.52) |
| DSD vs SoC | Key | 3 | 1.41 (1.09 to 1.84) |
| DSD vs SUP | General | 1 | 0.91 (0.67 to 1.24) |
| DSD vs SUP | Key | 1 | 1.17 (0.80 to 1.72) |
| EDU vs REM | General | 1 | 0.88 (0.73 to 1.07) |
| EDU vs SoC | General | 1 | 1.28 (0.92 to 1.79) |
| mHealth vs REM | Key | 1 | 1.55 (0.96 to 2.51) |
| mHealth vs SoC | General | 7 | 0.56 (0.55 to 0.57) |
| mHealth vs SoC | Key | 3 | 1.13 (0.78 to 1.62) |
| REM vs SoC | General | 4 | 1.45 (1.02 to 2.08) |
| REM vs SoC | Key | 1 | 0.72 (0.45 to 1.16) |
| SoC vs SUP | General | 2 | 0.70 (0.52 to 0.94) |
| SoC vs SUP | Key | 2 | 0.83 (0.60 to 1.15) |

General versus key population classification followed study reporting.^99^ Estimates were descriptive because many subgroups had k ≤ 1

#### Table S36. WHO geographical regions: Subgroup analysis for viral load suppressions

| Comparisons | Geographical regions | k | OR (95% CI) |
| --- | --- | --- | --- |
| BC vs EDU | AMRO | 1 | 0.89 (0.52 to 1.52) |
| BC vs SoC | AFRO | 2 | 1.66 (1.26 to 2.18) |
| BC vs SoC | AMRO | 3 | 0.91 (0.62 to 1.33) |
| BC vs SUP | AFRO | 1 | 1.16 (0.80 to 1.67) |
| BC vs SUP | AMRO | 1 | 0.89 (0.63 to 1.26) |
| DSD vs SoC | AFRO | 7 | 1.36 (1.19 to 1.56) |
| DSD vs SoC | AMRO | 1 | 0.86 (0.48 to 1.52) |
| DSD vs SUP | AFRO | 1 | 0.95 (0.73 to 1.24) |
| DSD vs SUP | AMRO | 1 | 0.84 (0.47 to 1.48) |
| EDU vs EI | AMRO | 1 | 0.67 (0.37 to 1.21) |
| EDU vs REM | AMRO | 1 | 0.86 (0.71 to 1.04) |
| EDU vs SoC | AFRO | 1 | 1.33 (0.88 to 2.00) |
| EI vs eSoC | SEARO | 1 | 1.79 (1.06 to 3.01) |
| EI vs mHealth | AFRO | 1 | 0.89 (0.57 to 1.38) |
| EI vs SoC | AFRO | 2 | 1.07 (0.71 to 1.63) |
| EI vs SoC | AMRO | 2 | 1.53 (0.98 to 2.38) |
| mHealth vs REM | AFRO | 1 | 1.61 (1.05 to 2.48) |
| mHealth vs SoC | AFRO | 7 | 1.21 (0.99 to 1.49) |
| mHealth vs SoC | AMRO | 3 | 0.56 (0.55 to 0.57) |
| REM vs SoC | AFRO | 3 | 0.75 (0.49 to 1.15) |
| REM vs SoC | AMRO | 2 | 1.19 (0.72 to 1.98) |
| SoC vs SUP | AFRO | 3 | 0.70 (0.54 to 0.90) |
| SoC vs SUP | AMRO | 1 | 0.97 (0.64 to 1.49) |

Subgroup analysis for WHO geographical regions^100^. Estimates were descriptive because many subgroups had k ≤ 1

#### Table S37. Study design (parallel vs cluster): Subgroup analysis for viral load suppression

| Comparisons | Designs | k | OR (95 % CI) |
| --- | --- | --- | --- |
| BC vs EDU | parallel | 1 | 1.09 (0.67 to 1.76) |
| BC vs eSoC | parallel | 1 | 5.20 (1.17 to 23.05) |
| BC vs SoC | cluster | 1 | 0.85 (0.35 to 2.02) |
| BC vs SoC | parallel | 4 | 1.45 (1.16 to 1.80) |
| BC vs SUP | parallel | 2 | 0.94 (0.71 to 1.23) |
| DSD vs DSD+MMD | cluster | 2 | 3.01 (1.67 to 5.39) |
| DSD vs DSD+MMD | parallel | 1 | 1.22 (0.56 to 2.69) |
| DSD vs SoC | cluster | 3 | 0.83 (0.59 to 1.18) |
| DSD vs SoC | parallel | 5 | 1.41 (1.23 to 1.63) |
| DSD vs SUP | cluster | 1 | 0.97 (0.65 to 1.43) |
| DSD vs SUP | parallel | 1 | 0.91 (0.69 to 1.21) |
| DSD+MMD vs SoC | cluster | 1 | 0.28 (0.15 to 0.53) |
| DSD+MMD vs SoC | parallel | 1 | 1.15 (0.52 to 2.54) |
| EDU vs EI | parallel | 1 | 0.44 (0.24 to 0.78) |
| EDU vs REM | parallel | 1 | 0.89 (0.73 to 1.08) |
| EDU vs SoC | cluster | 1 | 1.33 (0.88 to 2.00) |
| EI vs eSoC | cluster | 1 | 1.79 (1.06 to 3.01) |
| EI vs mHealth | cluster | 1 | 0.63 (0.38 to 1.05) |
| EI vs SoC | cluster | 2 | 0.65 (0.44 to 0.98) |
| EI vs SoC | parallel | 2 | 3.04 (1.92 to 4.82) |
| mHealth vs REM | cluster | 1 | 1.48 (0.92 to 2.40) |
| mHealth vs SoC | cluster | 2 | 1.03 (0.71 to 1.50) |
| mHealth vs SoC | parallel | 8 | 0.56 (0.55 to 0.57) |
| REM vs SoC | cluster | 1 | 0.70 (0.43 to 1.11) |
| REM vs SoC | parallel | 4 | 1.50 (0.94 to 2.39) |
| SoC vs SUP | cluster | 1 | 1.16 (0.76 to 1.77) |
| SoC vs SUP | parallel | 3 | 0.65 (0.50 to 0.83) |
| SoC vs TS | parallel | 1 | 0.35 (0.18 to 0.69) |
| SUP vs TS | cluster | 1 | 1.00 (0.48 to 2.06) |

Estimates were descriptive because many subgroups had k ≤ 1

### Figures

These figures provide graphical displays of intervention distributions, risk-of-bias assessments, and direct pairwise comparisons. Forest plots (Figures S7–S24 and S26–S30) are provided for transparency; interpretation is based on network estimates rather than individual direct comparisons

### Figures for distributions of intervention categories

#### Figure S1. Distribution of intervention categories across World Bank income levels.

#### Figure S2. Distribution of intervention categories across WHO geographical regions.

#### Figure S3. Distribution of intervention categories across population types.

### Figures for risk of bias assessments

#### Figure S4. Risk‑of‑bias assessment for retention outcomes using the ROBUST‑RCT instrument.

#### Figure S5. Risk‑of‑bias assessment for viral load outcomes using the ROBUST‑RCT instrument.

#### Figure S6. Risk‑of‑bias assessment for quality‑of‑life outcomes using the ROBUST‑RCT instrument.

### Forest plots for retention in HIV care

#### Figure S7. Forest plot of direct comparisons of BC vs EDU for retention in HIV care.

#### Figure S8. Forest plot of direct comparisons of BC vs SoC for retention in HIV care.

#### Figure S9. Forest plot of direct comparisons of CM vs SoC for retention in HIV care.

#### Figure S10. Forest plot of direct comparisons of DSD vs DSD_MMD for retention in HIV care.

#### Figure S11. Forest plot of direct comparisons of DSD vs SoC for retention in HIV care.

#### Figure S12. Forest plot of direct comparisons of DSD vs SUP for retention in HIV care.

#### Figure S13. Forest plot of direct comparisons of EDU vs SoC for retention in HIV care.

#### Figure S14. Forest plot of direct comparisons of EI vs eSoC for retention in HIV care.

#### Figure S15. Forest plot of direct comparisons of EI vs SoC for retention in HIV care.

#### Figure S16. Forest plot of direct comparisons of mHealth vs REM for retention in HIV care.

#### Figure S17. Forest plot of direct comparisons of mHealth vs SoC for retention in HIV care.

#### Figure S18. Forest plot of direct comparisons of MM vs SoC for retention in HIV care.

#### Figure S19. Forest plot of direct comparisons of REM vs EDU for retention in HIV care.

#### Figure S20. Forest plot of direct comparisons of REM vs SoC for retention in HIV care.

#### Figure S21. Forest plot of direct comparisons of SoC vs SUP for retention in HIV care

#### Figure S22. Forest plot of direct comparisons of SUP vs BC for retention in HIV care.

#### Figure S23. Forest plot of direct comparisons of SUP vs SoC for retention in HIV care.

#### Figure S24. Forest plot of direct comparisons of TS vs SoC for retention in HIV care.

#### Figure S25. Net heat plot of network incoherence for retention in HIV care.

Warmer cells indicate a greater contribution to local incoherence. Patterns were local and did not suggest network‑wide incoherence

### Forest plots for viral load suppression

#### Figure S26. Forest plot of direct comparisons of BC vs SoC for viral load suppression.

#### Figure S27. Forest plot of direct comparisons of DSD vs DSD-MMD for viral load suppression.

#### Figure S28. Forest plot of direct comparisons of DSD vs SoC for viral load suppression.

#### Figure S29. Forest plot of direct comparisons of DSD vs SUP for viral load suppression.

#### Figure S30. Forest plot of direct comparisons of EI vs SoC for viral load suppression.

Figure S31. Forest plot of direct comparisons of mHealth vs SoC for viral load suppression.

Figure S32. Forest plot of direct comparisons of REM vs SoC for viral load suppression.

Figure S33. Forest plot of direct comparisons of SUP vs BC for viral load suppression.

Figure S34. Forest plot of direct comparisons of SUP vs SoC for viral load suppression.

References

1.	Fayorsey RN, Wang C, Chege D, Reidy W, Syengo M, Owino SO, et al. Effectiveness of a Lay Counselor-Led Combination Intervention for Retention of Mothers and Infants in HIV Care: A Randomised Trial in Kenya. J Acquir Immune Defic Syndr. 2019;80(1):56-63.

2.	Washington S, Owuor K, Turan JM, Steinfeld RL, Onono M, Shade SB, et al. The effect of integration of HIV care and treatment into antenatal care clinics on mother-to-child HIV transmission and maternal outcomes in Nyanza, Kenya: results from the SHAIP cluster randomised controlled trial. J Acquir Immune Defic Syndr. 2015;69(5):e164.

3.	Maskew M, Brennan AT, Fox MP, Vezi L, Venter WDF, Ehrenkranz P, et al. A clinical algorithm for same-day HIV treatment initiation in settings with high TB symptom prevalence in South Africa: the SLATE II individually randomised clinical trial. PLoS medicine. 2020;17(8):e1003226.

4.	Khan S, Spiegelman D, Walsh F, et al. Early access to antiretroviral therapy versus standard of care among HIV-positive participants in Eswatini in the public health sector: the MaxART stepped-wedge randomised controlled trial. Journal of the International AIDS Society. 2020;23(9):e25610.

5.	Jani IV, Meggi B, Loquiha O, Tobaiwa O, Mudenyanga C, Zitha A, et al. Effect of point-of-care early infant diagnosis on antiretroviral therapy initiation and retention of patients. AIDS. 2018;32(11):1453-63.

6.	Sarna A, Saraswati LR, Okal J, Matheka J, Owuor D, Singh RJ, et al. Cell Phone Counseling Improves Retention of Mothers With HIV Infection in Care and Infant HIV Testing in Kisumu, Kenya: a Randomised Controlled Study. Global health, science and practice. 2019;7(2):171‐88.

7.	Audet CM, Graves E, Emílio AM, Matino A, Paulo P, Aboobacar AM, et al. Effect of a storytelling intervention on the retention of serodiscordant couples in ART/PrEP services at antenatal clinic in Namacurra province in Zambézia, Mozambique. Contemporary Clinical Trials Communications. 2021;22:100782.

8.	Odeny TA, Hughes JP, Bukusi EA, Akama E, Geng EH, Holmes KK, et al. Text messaging for maternal and infant retention in prevention of mother-to-child HIV transmission services: A pragmatic steppedwedge cluster-randomised trial in Kenya. PLoS Medicine. 2019;16(10):e1002924.

9.	Rotheram-Borus MJ, Rice E, Comulada WS, Best K, Elia C, Peters K, et al. Intervention outcomes among HIV-affected families over 18 months. AIDS Behav. 2012;16(5):1265-75.

10.	Dorvil N, Rivera VR, Riviere C, Berman R, Severe P, Bang H, et al. Same-day testing with initiation of antiretroviral therapy or tuberculosis treatment versus standard care for persons presenting with tuberculosis symptoms at HIV diagnosis: A randomised open-label trial from Haiti. PLoS medicine. 2023;20(6):e1004246.

11.	Chandra DK, Bazazi AR, Nahaboo Solim MA, Kamarulzaman A, Altice FL, Culbert GJ. Retention in clinical trials after prison release: results from a clinical trial with incarcerated men with HIV and opioid dependence in Malaysia. HIV Res Clin Pract. 2019;20(1):12-23.

12.	Rosen S, Maskew M, Larson BA, Brennan AT, Tsikhutsu I, Fox MP, et al. Simplified clinical algorithm for identifying patients eligible for same-day HIV treatment initiation (SLATE): Results from an individually randomised trial in South Africa and Kenya. PLoS Med. 2019;16(9):e1002912.

13.	Mbuagbaw L, Thabane L, Ongolo-Zogo P, et al. The Cameroon Mobile Phone SMS (CAMPS) trial: a randomised trial of text messaging versus usual care for adherence to antiretroviral therapy. PLoS One. 2012;7(12):e46909.

14.	Lucas GM, Chaudhry A, Hsu J, Woodson T, Lau B, Olsen Y, et al. Clinic-based treatment of opioid-dependent HIV-infected patients versus referral to an opioid treatment program: a randomised trial. Annals of internal medicine. 2010;152(11):704-11.

15.	Keitz SA, Box TL, Homan RK, Bartlett JA, Oddone EZ. Primary care for patients infected with human immunodeficiency virus: a randomised controlled trial. Journal of general internal medicine. 2001;16(9):573-82.

16.	Odeny TA, Bukusi EA, Cohen CR, Yuhas K, Camlin CS, McClelland RS. Texting improves testing: a randomised trial of two-way SMS to increase postpartum prevention of mother-to-child transmission retention and infant HIV testing. Aids. 2014;28(15):2307-12.

17.	Chang LW, Kagaayi J, Nakigozi G, Ssempijja V, Packer AH, Serwadda D, et al. Effect of peer health workers on AIDS care in Rakai, Uganda: a cluster-randomised trial. PloS one. 2010;5(6):e10923.

18.	Wohl DA, Scheyett A, Golin CE, White B, Matuszewski J, Bowling M, et al. Intensive case management before and after prison release is no more effective than comprehensive pre-release discharge planning in linking HIV-infected prisoners to care: a randomised trial. AIDS and Behaviour. 2011;15(2):356-64.

19.	Naar-King S, Outlaw A, Green-Jones M, Wright K, Parsons JT. Motivational interviewing by peer outreach workers: a pilot randomised clinical trial to retain adolescents and young adults in HIV care. AIDS Care. 2009;21(7):868-73.

20.	Gwadz M, Cleland CM, Applegate E, et al. Behavioural intervention improves treatment outcomes among HIV-infected individuals who have delayed, declined, or discontinued antiretroviral therapy: a randomised controlled trial of a novel intervention. AIDS Behav. 2015;19(10):1801-17.

21.	Norton BL, Person AK, Castillo C, Pastrana C, Subramanian M, Stout JE. Barriers to using text message appointment reminders in an HIV clinic. Telemed J E Health. 2014;20(1):86-9.

22.	MacGowan RJ, Lifshay J, Mizuno Y, Johnson WD, McCormick L, Zack B. Positive Transitions (POST): Evaluation of an HIV Prevention Intervention for HIV-Positive Persons Releasing from Correctional Facilities. AIDS Behav. 2015;19(6):1061-9.

23.	Gardner LI, Giordano TP, Marks G, Wilson TE, Craw JA, Drainoni ML, et al. Enhanced personal contact with HIV patients improves retention in primary care: a randomised trial in 6 US HIV clinics. Clin Infect Dis. 2014;59(5):725-34.

24.	Konkle-Parker DJ, Amico KR, McKinney VE. Effects of an intervention addressing information, motivation, and behavioural skills on HIV care adherence in a southern clinic cohort. AIDS Care. 2014;26(6):674-83.

25.	Huang D, Sangthong R, McNeil E, Chongsuvivatwong V, Zheng W, Yang X. Effects of a Phone Call Intervention to Promote Adherence to Antiretroviral Therapy and Quality of Life of HIV/AIDS Patients in Baoshan, China: A Randomised Controlled Trial. AIDS Res Treat. 2013;2013:580974.

26.	Wamalwa DC, Farquhar C, Obimbo EM, Selig S, Mbori-Ngacha DA, Richardson BA, et al. Medication diaries do not improve outcomes with highly active antiretroviral therapy in Kenyan children: a randomised clinical trial. J Int AIDS Soc. 2009;12:8.

27.	Wohl AR, Garland WH, Valencia R, Squires K, Witt MD, Kovacs A, et al. A Randomised Trial of Directly Administered Antiretroviral Therapy and Adherence Case Management Intervention. Clinical Infectious Diseases. 2006;42(11):1619-27.

28.	Chander G, Hutton HE, Lau B, Xu X, McCaul ME. Brief intervention decreases drinking frequency in HIV-infected, heavy drinking women: results of a randomised controlled trial. JAIDS Journal of Acquired Immune Deficiency Syndromes. 2015;70(2):137-45.

29.	Kunutsor S, Walley J, Katabira E, Muchuro S, Balidawa H, Namagala E, et al. Improving clinic attendance and adherence to antiretroviral therapy through a treatment supporter intervention in Uganda: a randomised controlled trial. AIDS and Behaviour. 2011;15(8):1795-802.

30.	Dulli L, Ridgeway K, Packer C, et al. A Social Media-Based Support Group for Youth Living with HIV in Nigeria (SMART Connections): Randomised Controlled Trial. Journal of Medical Internet Research. 2020;22(6):e18343.

31.	Ammassari A, Stöhr W, Antinori A, Molina J-M, Schwimmer C, Domingo P, et al. Patient self-reported adherence to ritonavir-boosted darunavir combined with either raltegravir or tenofovir disoproxil fumarate/emtricitabine in the NEAT001/ANRS143 trial. JAIDS Journal of Acquired Immune Deficiency Syndromes. 2018;79(4):481-90.

32.	Graves JC, Elyanu P, Schellack CJ, Asire B, Prust ML, Prescott MR, et al. Impact of a Family Clinic Day intervention on paediatric and adolescent appointment adherence and retention in antiretroviral therapy: A cluster randomised controlled trial in Uganda. PLoS One. 2018;13(3):e0192068.

33.	El-Sadr WM, Beauchamp G, Hall HI, Torian LV, Zingman BS, Lum G, et al. Brief Report: Durability of the Effect of Financial Incentives on HIV Viral Load Suppression and Continuity in Care: HPTN 065 Study. J Acquir Immune Defic Syndr. 2019;81(3):300-3.

34.	Fahey C, Njau P, Katabaro E, Mfaume R, Ulenga N, Mwenda N, et al. Effects of financial incentives for clinic attendance on HIV viral suppression among adults initiating antiretroviral therapy in Tanzania: A three-arm randomised controlled trial. Journal of the International Aids Society. 2020;23:67-.

35.	Myer L, Phillips TK, Zerbe A, Brittain K, Lesosky M, Hsiao NY, et al. Integration of postpartum healthcare services for HIV-infected women and their infants in South Africa: A randomised controlled trial. PLoS Med. 2018;15(3):e1002547.

36.	McLaughlin MM, Franke MF, Muñoz M, Nelson AK, Saldaña O, Cruz JS, et al. Community-Based Accompaniment with Supervised Antiretrovirals for HIV-Positive Adults in Peru: a Cluster-Randomised Trial. AIDS and behaviour. 2018;22(1):287‐96.

37.	Kadota JL, Fahey CA, Njau PF, Kapologwe N, Padian NS, Dow WH, et al. The heterogeneous effect of short-term transfers for improving ART adherence among HIV-infected Tanzanian adults. AIDS care. 2018;30(sup3):18-26.

38.	Kalichman SC, Cherry C, Kalichman MO, Eaton LA, Kohler JJ, Montero C, et al. Mobile health intervention to reduce HIV transmission: a randomised trial of behaviourally enhanced HIV treatment as prevention (B-TasP). JAIDS Journal of Acquired Immune Deficiency Syndromes. 2018;78(1):34-42.

39.	Neduzhko O, Postnov O, Sereda Y, Kulchynska R, Bingham T, Myers JJ, et al. Modified Antiretroviral Treatment Access Study (MARTAS): A Randomised Controlled Trial of the Efficacy of a Linkage-to-Care Intervention Among HIV-Positive Patients in Ukraine. AIDS Behav. 2020;24(11):3142-54.

40.	Kim MH, Ahmed S, Tembo T, Sabelli R, Flick R, Yu X, et al. VITAL Start: Video-Based intervention to Inspire Treatment Adherence for Life—pilot of a novel video-based approach to HIV counseling for pregnant women living with HIV. AIDS and Behaviour. 2019;23:3140-51.

41.	Mavhu W, Willis N, Mufuka J, et al. Effect of a differentiated service delivery model on virological failure in adolescents with HIV in Zimbabwe (Zvandiri): a cluster-randomised controlled trial. The Lancet Global Health. 2020;8(2):e264-e75.

42.	Horvath KJ, Lammert S, Maclehose RF, Danh T, Baker JV, Carrico AW. A Pilot Study of a Mobile App to Support HIV Antiretroviral Therapy Adherence Among Men Who Have Sex with Men Who Use Stimulants. AIDS and Behaviour. 2019;23(11):3184-98.

43.	Samet JH, Blokhina E, Cheng DM, Walley AY, Lioznov D, Gnatienko N, et al. A strengths-based case management intervention to link HIV-positive people who inject drugs in Russia to HIV care. AIDS. 2019;33(9):1467-76.

44.	Sherman EM, Niu J, Elrod S, Clauson KA, Alkhateeb F, Eckardt P. Effect of mobile text messages on antiretroviral medication adherence and patient retention in early HIV care: an open-label, randomised, single center study in south Florida. AIDS Res Ther. 2020;17(1):16.

45.	Willis N, Milanzi A, Mawodzeke M, Dziwa C, Armstrong A, Yekeye I, et al. Effectiveness of community adolescent treatment supporters (CATS) interventions in improving linkage and retention in care, adherence to ART and psychosocial well-being: a randomised trial among adolescents living with HIV in rural Zimbabwe. BMC Public Health. 2019;19(1):117.

46.	Sabin LL, Halim N, Hamer DH, et al. Retention in HIV Care Among HIV-Seropositive Pregnant and Postpartum Women in Uganda: results of a Randomised Controlled Trial. AIDS and behaviour. 2020;24(11):3164‐75.

47.	Pascoe SJ, Fox MP, Huber AN, Murphy J, Phokojoe M, Gorgens M, et al. Differentiated HIV care in South Africa: the effect of fast-track treatment initiation counselling on ART initiation and viral suppression as partial results of an impact evaluation on the impact of a package of services to improve HIV treatment adherence. J Int AIDS Soc. 2019;22(11):e25409.

48.	Silverman K, Holtyn AF, Rodewald AM, Siliciano RF, Jarvis BP, Subramaniam S, et al. Incentives for Viral Suppression in People Living with HIV: A Randomised Clinical Trial. AIDS Behav. 2019;23(9):2337-46.

49.	Goodrich S, Siika A, Mwangi A, Nyambura M, Naanyu V, Yiannoutsos C, et al. Development, assessment, and outcomes of a community-based model of antiretroviral care in western Kenya through a cluster-randomised control trial. JAIDS Journal of Acquired Immune Deficiency Syndromes. 2021;87(2):e198-e206.

50.	Byonanebye DM, Nabaggala MS, Naggirinya AB, Lamorde M, Oseku E, King R, et al. An Interactive Voice Response Software to Improve the Quality of Life of People Living With HIV in Uganda: randomised Controlled Trial. JMIR mhealth and uhealth. 2021;9(2):e22229.

51.	Hoffman RM, Moyo C, Balakasi KT, Siwale Z, Hubbard J, Bardon A, et al. Multimonth dispensing of up to 6 months of antiretroviral therapy in Malawi and Zambia (INTERVAL): a cluster-randomised, non-blinded, non-inferiority trial. The lancet Global health. 2021;9(5):e628‐e38.

52.	Cassidy T, Grimsrud A, Keene C, Lebelo K, Hayes H, Orrell C, et al. Twenty-four-month outcomes from a cluster-randomised controlled trial of extending antiretroviral therapy refills in ART adherence clubs. Journal of the International AIDS Society. 2020;23(12):e25649.

53.	Kinuthia J, Ronen K, Unger JA, Jiang W, Matemo D, Perrier T, et al. RCT of 2-way vs 1-way sms messaging to improve efficacy of pmtct-art in Kenya. Topics in antiviral medicine. 2021;29(1):215‐.

54.	Ayer R, Poudel KC, Kikuchi K, Ghimire M, Shibanuma A, Jimba M. Nurse-Led Mobile Phone Voice Call Reminder and On-Time Antiretroviral Pills Pick-Up in Nepal: A Randomised Controlled Trial. AIDS Behav. 2021;25(6):1923-34.

55.	Bien-Gund CH, Ho JI, Bair EF, Marcus N, Choi RJ, Szep Z, et al. Brief report: financial incentives and real-time adherence monitoring to promote daily adherence to HIV treatment and viral suppression among people living with HIV: a pilot study. JAIDS Journal of Acquired Immune Deficiency Syndromes. 2021;87(1):688-92.

56.	Graham SM, Micheni M, Chirro O, Nzioka J, Secor AM, Mugo PM, et al. A Randomised Controlled Trial of the Shikamana Intervention to Promote Antiretroviral Therapy Adherence Among Gay, Bisexual, and Other Men Who Have Sex with Men in Kenya: Feasibility, Acceptability, Safety and Initial Effect Size. AIDS Behav. 2020;24(7):2206-19.

57.	Ndhlovu CE, Kouamou V, Nyamayaro P, Dougherty L, Willis N, Ojikutu BO, et al. The transient effect of a peer support intervention to improve adherence among adolescents and young adults failing antiretroviral therapy in Harare, Zimbabwe: a randomised control trial. AIDS Res Ther. 2021;18(1):32.

58.	Fatti G, Ngorima-Mabhena N, Mothibi E, et al. Outcomes of Three- Versus Six-Monthly Dispensing of Antiretroviral Treatment (ART) for Stable HIV Patients in Community ART Refill Groups: A Cluster-Randomised Trial in Zimbabwe. J Acquir Immune Defic Syndr. 2020;84(2):162-72.

59.	Wagner GJ, Hoffman R, Linnemayr S, Schneider S, Ramirez D, Gordon K, et al. START (Supporting Treatment Adherence Readiness through Training) Improves Both HIV Antiretroviral Adherence and Viral Reduction, and is Cost Effective: results of a Multi-site Randomised Controlled Trial. AIDS and behaviour. 2021.

60.	Stephenson R, Garofalo R, Sullivan PS, Hidalgo MA, Bazzi AR, Hoehnle S, et al. Stronger Together: Results from a Randomised Controlled Efficacy Trial of a Dyadic Intervention to Improve Engagement in HIV Care Among Serodiscordant Male Couples in Three US Cities. AIDS Behav. 2021;25(8):2369-81.

61.	Tukei BB, Fatti G, Tiam A, et al. Community-based multimonth dispensing of ART: a cluster randomised trial in Lesotho. Topics in antiviral medicine. 2020;28(1):15‐.

62.	Roy M, Glidden DV, Geng E, Sikazwe I, Mukumbwa-Mwenechanya M, Efronson E, et al. Participation in adherence clubs and on-time drug pickup among HIV-infected adults in Zambia: A matched-pair cluster randomised trial. PLoS Medicine. 2020;17(7):e1003116.

63.	Drain PK, Dorward J, Violette LR, Quame-Amaglo J, Thomas KK, Samsunder N, et al. Point-of-care HIV viral load testing combined with task shifting to improve treatment outcomes (STREAM): findings from an open-label, non-inferiority, randomised controlled trial. The lancet HIV. 2020;7(4):e229-e37.

64.	Giordano TP, Cully J, Amico KR, Davila JA, Kallen MA, Hartman C, et al. A randomised trial to test a peer mentor intervention to improve outcomes in persons hospitalized with HIV infection. Clinical Infectious Diseases. 2016;63(5):678-86.

65.	Liu H, Wang Y, Huang Y, Xiong D, Shen J, Siqueiros L, et al. Ingestible sensor system for measuring, monitoring and enhancing adherence to antiretroviral therapy: an open-label, usual care-controlled, randomised trial. EBioMedicine. 2022;86.

66.	Hickey MD, Ouma GB, Mattah B, Pederson B, DesLauriers NR, Mohamed P, et al. The Kanyakla study: Randomised controlled trial of a microclinic social network intervention for promoting engagement and retention in HIV care in rural western Kenya. PloS one. 2021;16(9):e0255945.

67.	Kebaya LM, Wamalwa D, Kariuki N, Admani B, Ayieko P, Nduati R. Efficacy of Mobile phone use on adherence to Nevirapine prophylaxis and retention in care among the HIV-exposed infants in prevention of mother to child transmission of HIV: a randomised controlled trial. BMC pediatrics. 2021;21(1):186.

68.	Hightow-Weidman L, Muessig KE, Egger JR, Vecchio A, Platt A. Epic allies: a gamified mobile app to improve engagement in HIV care and antiretroviral adherence among young men who have sex with men. AIDS and Behaviour. 2021;25:2599-617.

69.	Lewis MA, Harshbarger C, Bann C, Marconi VC, Somboonwit C, Dalla Piazza M, et al. Effectiveness of an interactive, highly tailored “video doctor” intervention to suppress viral load and retain patients with HIV in clinical care: a randomised clinical trial. JAIDS Journal of Acquired Immune Deficiency Syndromes. 2022;91(1):58-67.

70.	Chang C, Agbaji O, Mitruka K, Olatunde B, Sule H, Dajel T, et al. Clinical outcomes in a randomised controlled trial comparing point-of-care with standard human immunodeficiency virus (HIV) viral load monitoring in Nigeria. Clinical Infectious Diseases. 2023;76(3):e681-e91.

71.	Fahey CA, Njau PF, Kelly NK, Mfaume RS, Bradshaw PT, Dow WH, et al. Durability of effects from short-term economic incentives for clinic attendance among HIV positive adults in Tanzania: long-term follow-up of a randomised controlled trial. BMJ Global Health. 2021;6(12).

72.	Metsch LR, Feaster DJ, Gooden LK, Masson C, Perlman DC, Jain MK, et al., editors. Care facilitation advances movement along the hepatitis C care continuum for persons with human immunodeficiency virus, hepatitis C, and substance use: a randomised clinical trial (CTN-0064). Open forum infectious diseases; 2021: Oxford University Press US.

73.	Amone A, Gabagaya G, Wavamunno P, Rukundo G, Namale-Matovu J, Malamba SS, et al. Enhanced peer-group strategies to support the prevention of mother-to-child HIV transmission leads to increased retention in care in Uganda: A randomised controlled trial. PLoS One. 2024;19(4):e0297652.

74.	Ayieko J, Balzer LB, Inviolata C, Kakande E, Opel F, Wafula EM, et al. Randomised trial of a “dynamic choice” patient-centred care intervention for Mobile persons with HIV in east Africa. JAIDS Journal of Acquired Immune Deficiency Syndromes. 2024;95(1):74-81.

75.	Bwanika Naggirinya A, Meya DB, Nabaggala MS, Banturaki G, Kiragga A, Rujumba J, et al. Effectiveness of interactive voice response-call for life mHealth tool on adherence to anti-retroviral therapy among young people living with HIV: A randomised trial in Uganda. PloS one. 2024;19(11):e0308923.

76.	Derose KP, Then-Paulino A, Han B, Armenta G, Palar K, Jimenez-Paulino G, et al. Preliminary effects of an urban gardens and peer nutritional counseling intervention on HIV treatment adherence and detectable viral load among people with HIV and food insecurity: evidence from a pilot cluster randomised controlled trial in the Dominican Republic. AIDS and Behaviour. 2023;27(3):864-74.

77.	Inghels M, Kim HY, Mathenjwa T, Shahmanesh M, Seeley J, Wyke S, et al. Population impacts of conditional financial incentives and a male‐targeted digital decision support application on the HIV treatment cascade in rural KwaZulu Natal: findings from the HITS cluster randomised clinical trial. Journal of the International AIDS Society. 2024;27(5):e26248.

78.	Luoma JB, Rossi SL, Sereda Y, Pavlov N, Toussova O, Vetrova M, et al. An acceptance-based, intersectional stigma coping intervention for people with HIV who inject drugs—a randomised clinical trial. The Lancet Regional Health–Europe. 2023;28.

79.	Mabuto T, Woznica DM, Ndini P, Moyo D, Abraham M, Hanrahan C, et al. Transitional community adherence support for people leaving incarceration in South Africa: a pragmatic, open-label, randomised controlled trial. The Lancet HIV. 2024;11(1):e11-e9.

80.	Martin TC, Smith LR, Anderson C, Little SJ. Randomised Controlled Trial of 60 minutes for Health With Rapid Antiretroviral Therapy to Reengage Persons With HIV Who Are Out of Care. JAIDS Journal of Acquired Immune Deficiency Syndromes. 2024;96(5):486-93.

81.	Ndongo FA, Noah J-PYA, Kana R, Ndie J, Nono M, Ndzie P, et al. A community-based peer-facilitated psychological and social support model to improve retention in care among Cameroonian adolescents perinatally infected with human immunodeficiency virus: A randomised controlled trial. Journal of Epidemiology and Population Health. 2024;72(6):202792.

82.	Njau PF, Katabaro E, Winters S, Sabasaba A, Hassan K, Joseph B, et al. Impact of financial incentives on viral suppression among adults initiating HIV treatment in Tanzania: a hybrid effectiveness–implementation trial. The Lancet HIV. 2024;11(9):e586-e97.

83.	Njuguna C, Long L, Mistri P, Chetty-Makkan C, Maughan-Brown B, Buttenheim A, et al. A randomised trial of ‘fresh start’text messaging to improve return to care in people with HIV who missed appointments in South Africa. Aids. 2024;38(10):1579-88.

84.	Novak MD, Holtyn AF, Toegel F, Rodewald AM, Leoutsakos J-M, Fingerhood M, et al. Long-Term effects of incentives for HIV viral suppression: A randomised clinical trial. AIDS and Behaviour. 2024;28(2):625-35.

85.	Onoya D, Sineke T, Mokhele I, Vujovic M, Holland K, Ruiter RA. Improving Retention and HIV Viral Suppression: A Cluster Randomised Pilot Trial of a Lay Counsellor Motivational Interviewing Training in South Africa. medRxiv. 2024.

86.	Palar K, Sheira LA, Frongillo EA, O’Donnell AA, Nápoles TM, Ryle M, et al. Food is medicine for human immunodeficiency virus: improved health and hospitalizations in the Changing Health through Food Support (CHEFS-HIV) pragmatic randomised trial. The Journal of Infectious Diseases. 2025;231(3):573-82.

87.	Marc JB, Pierre S, Ducatel O, Homeus F, Zion A, Rivera VR, et al. Early initiation of fast‐track care for persons living with HIV initiating dolutegravir‐based regimens during a period of severe civil unrest in Port‐au‐Prince, Haiti: a pilot randomised trial. Journal of the International AIDS Society. 2025;28(2):e26419.

88.	Parry CD, Myers B, Londani M, Shuper PA, Janse van Rensburg C, Manda SO, et al. Motivational interviewing and problem‐solving therapy intervention for patients on antiretroviral therapy for HIV in Tshwane, South Africa: A randomised controlled trial to assess the impact on alcohol consumption. Addiction. 2023;118(11):2164-76.

89.	Peck RN, Issarow B, Kisigo GA, Kabakama S, Okello E, Rutachunzibwa T, et al. Linkage case management and posthospitalization outcomes in people with HIV: the Daraja randomised clinical trial. JAMA. 2024;331(12):1025-34.

90.	Reid MJ, Steenhoff AP, Thompson J, Gabaitiri L, Cary MS, Steele K, et al. Evaluation of the effect of cellular SMS reminders on consistency of antiretroviral therapy pharmacy pickups in HIV-infected adults in Botswana: a randomised controlled trial. Health psychology and behavioural medicine. 2017;5(1):101-9.

91.	Samet JH, Blokhina E, Cheng DM, Rosen S, Lioznov D, Lunze K, et al. Rapid access to antiretroviral therapy, receipt of naltrexone, and strengths-based case management versus standard of care for HIV viral load suppression in people with HIV who inject drugs in Russia (LINC-II): an open-label, randomised controlled trial. The Lancet HIV. 2023;10(9):e578-e87.

92.	Solomon SS, McFall AM, Srikrishnan AK, Verma V, Anand S, Khan RT, et al. Voucher incentives to improve viral suppression among HIV-positive people who inject drugs and men who have sex with men in India: a cluster randomised trial. Lancet HIV. 2024;11(5):e309-e20.

93.	Zani B, Fairall L, Petersen I, Folb N, Bhana A, Hanass-Hancock J, et al. Effectiveness of a task-sharing collaborative care model for the detection and management of depression among adults receiving antiretroviral therapy in primary care facilities in South Africa: A pragmatic cluster randomised controlled trial. J Affect Disord. 2025;370:499-510.

94.	Giovenco D, Pettifor A, Qayiya Y, Jones J, Bekker LG. The Acceptability, Feasibility, and Preliminary Effectiveness of a Courier HIV-Treatment Delivery and SMS Support Intervention for Young People Living With HIV in South Africa. J Acquir Immune Defic Syndr. 2024;95(2):161-9.

95.	Limbada M, Bwalya C, Macleod D, Floyd S, Schaap A, Situmbeko V, et al. A comparison of different community models of antiretroviral therapy delivery with the standard of care among stable HIV+ patients: rationale and design of a non-inferiority cluster randomised trial, nested in the HPTN 071 (PopART) study. Trials. 2021;22(1):52.

96.	Wieland LS, Berman BM, Altman DG, Barth J, Bouter LM, D'Adamo CR, et al. Rating of included trials on the efficacy–effectiveness spectrum: development of a new tool for systematic reviews. Journal of clinical epidemiology. 2017;84:95-104.

97.	Montgomery P, Grant S, Mayo-Wilson E, Macdonald G, Michie S, Hopewell S, et al. Reporting randomised trials of social and psychological interventions: the CONSORT-SPI 2018 Extension. Trials. 2018;19(1):407.

98.	World Health Organization. HIV statistics, globally and by WHO region, 2025: . information sheet. World Health Organization; 2025.

99.	World Health Organization. Retention in HIV programmes: defining the challenges and identifying solutions: meeting report, 13–15 September 2011. 2012.

100.	World Health Organization. Classifications and standards - Country groupings [Available from: <https://www.who.int/observatories/global-observatory-on-health-research-and-development/classifications-and-standards/country-groupings>

101.	World Bank. World Bank income groups. 2023.

102.	Kanters S, Park JJ, Chan K, Socias ME, Ford N, Forrest JI, et al. Interventions to improve adherence to antiretroviral therapy: a systematic review and network meta-analysis. Lancet HIV. 2017;4(1):e31-e40.

103.	Schandelmaier S, Briel M, Varadhan R, Schmid CH, Devasenapathy N, Hayward RA, et al. Development of the Instrument to assess the Credibility of Effect Modification Analyses (ICEMAN) in randomised controlled trials and meta-analyses. CMAJ. 2020;192(32):E901-E6.

104.	Wang Y KS, Briel M, Glasziou P, Brignardello-Petersen R,. Development of the Risk of Bias Instrument for Use in Systematic Reviews - for Randomised Controlled Trials (ROBUST-RCT). Under review by BMJ. 2025.

105.	Rehman N, Wu M, Garcia C, Leenus A, El-Kechen H, Bhandari M, et al. Measures of retention in HIV care: a study within a review. AIDS Patient Care STDs. 2023;37(4):192–8.
