## Supplementary File 1. Prisma checklist for "Interventions to improve retention in HIV care: a systematic review and network meta-analysis of randomised controlled trials"

*[1]* **PRISMA NMA Checklist of Items to Include When Reporting A Systematic Review Involving a Network Meta-analysis-2015**

| **Section/Topic** | **Item #** | **Checklist Item** | **Reported on Page #** |
| --- | --- | --- | --- |
| **TITLE** |  |  |  |
| Title | 1 | Identify the report as a systematic review *incorporating a network meta-analysis (or related form of meta-analysis).* | Title page |
| **ABSTRACT** |  |  |  |
| Structured summary | 2 | Provide a structured summary including, as applicable:  **Background:** main objectives  **Methods:** data sources; study eligibility criteria, participants, and interventions; study appraisal; and *synthesis methods, such as network meta-analysis.*  **Results:** number of studies and participants identified; summary estimates with corresponding confidence/credible intervals; *treatment rankings may also be discussed. Authors may choose to summarize pairwise comparisons against a chosen treatment included in their analyses for brevity.*  **Discussion/Conclusions:** limitations; conclusions and implications of findings.  **Other:** primary source of funding; systematic review registration number with registry name. | Abstract |
| **INTRODUCTION** |  |  |  |
| Rationale | 3 | Describe the rationale for the review in the context of what is already known*, including mention of why a network meta-analysis has been conducted.* | Introduction, paragraphs 1-4; pages 5- 6 |
| Objectives | 4 | Provide an explicit statement of questions being addressed, with reference to participants, interventions, comparisons, outcomes, and study design (PICOS). | Introduction, final paragraph; Page 6 |
| **METHODS** |  |  |  |
| Protocol and registration | 5 | Indicate whether a review protocol exists and if and where it can be accessed (e.g., Web address); and, if available, provide registration information, including registration number. | Methods - first paragraph; PROSPERO CRD42024589177; Page 6 |
| Eligibility criteria | 6 | Specify study characteristics (e.g., PICOS, length of follow-up) and report characteristics (e.g., years considered, language, publication status) used as criteria for eligibility, giving rationale. *Clearly describe eligible treatments included in the* ***treatment network, and note whether any have been clustered or merged into the same node (with justification).*** | Methods - Eligibility criteria; Page 6; Interventions; Table 1 |
| Information sources | 7 | Describe all information sources (e.g., databases with dates of coverage, contact with study authors to identify additional studies) in the search and date last searched. | Methods - Data sources and searches; Page 6-7 |
| Search | 8 | Present full electronic search strategy for at least one database, including any limits used, such that it could be repeated. | Appendix S1 (Supplementary File 2) |
| Study selection | 9 | State the process for selecting studies (i.e., screening, eligibility, included in systematic review, and, if applicable, included in the meta-analysis). | Methods - Data collection; Figure 1 |
| Data collection process | 10 | Describe method of data extraction from reports (e.g., piloted forms, independently, in duplicate) and any processes for obtaining and confirming data from investigators. | Methods - Data collection; Page 8 |
| Data items | 11 | List and define all variables for which data were sought (e.g., PICOS, funding sources) and any assumptions and simplifications made. | Methods - Data collection; Tables S2-S4 |
| **Geometry of the network** | **S1** | Describe methods used to explore the geometry of the treatment network under study and potential biases related to it. This should include how the evidence base has been graphically summarized for presentation, and what characteristics were compiled and used to describe the evidence base to readers. | Methods - Network visualization and structural assessment; Page 11; Figures S1-S3 |
| Risk of bias within individual studies | 12 | Describe methods used for assessing risk of bias of individual studies (including specification of whether this was done at the study or outcome level), and how this information is to be used in any data synthesis. | Methods - Risk of Bias Assessment; Page 9-10 |
| Summary measures | 13 | State the principal summary measures (e.g., risk ratio, difference in means). | Methods - Data synthesis and analysis; Page 11-12 |
| Planned methods of analysis | 14 | Describe the methods of handling data and combining results of studies for each network meta-analysis. This should include, but not be limited to:   - *Handling of multi-arm trials;* - *Selection of variance structure;* - *Selection of prior distributions in Bayesian analyses; and* - *Assessment of model fit.* | Methods - Data synthesis and analysis; Assumptions; Page 10-11 |
| **Assessment of Inconsistency** | **S2** | Describe the statistical methods used to evaluate the agreement of direct and indirect evidence in the treatment network(s) studied. Describe efforts taken to address its presence when found. | Methods - Assumptions; Incoherence; Page 11 |
| Risk of bias across studies | 15 | Specify any assessment of risk of bias that may affect the cumulative evidence (e.g., publication bias, selective reporting within studies). | Methods - Certainty of evidence (GRADE); Page 14-16 |
| Additional analyses | 16 | Describe methods of additional analyses if done, indicating which were pre-specified. This may include, but not be limited to, the following:   - Sensitivity or subgroup analyses; - Meta-regression analyses; - *Alternative formulations of the treatment network; and* - *Use of alternative prior distributions for Bayesian analyses (if applicable).* | Methods - Subgroup analysis; Page 12-14 |
| **RESULTS†** |  |  |  |
| Study selection | 17 | Give numbers of studies screened, assessed for eligibility, and included in the review, with reasons for exclusions at each stage, ideally with a flow diagram. | Results - Study selection; Figure 1 |
| **Presentation of network structure** | **S3** | Provide a network graph of the included studies to enable visualization of the geometry of the treatment network. | Results - Figures 2 and 4 |
| **Summary of network geometry** | **S4** | Provide a brief overview of characteristics of the treatment network. This may include commentary on the abundance of trials and randomized patients for the different interventions and pairwise comparisons in the network, gaps of evidence in the treatment network, and potential biases reflected by the network structure. | Results - Retention in care; Viral load suppression; Page 18-21 |
| Study characteristics | 18 | For each study, present characteristics for which data were extracted (e.g., study size, PICOS, follow-up period) and provide the citations. | Results - Study characteristics; Table 2; Tables S2-S4 |
| Risk of bias within studies | 19 | Present data on risk of bias of each study and, if available, any outcome level assessment. | Results - Risk of Bias; Figures S4-S6 |
| Results of individual studies | 20 | For all outcomes considered (benefits or harms), present, for each study: 1) simple summary data for each intervention group, and 2) effect estimates and confidence intervals. *Modified approaches may be needed to deal with information from larger networks.* | Supplementary File 2 - Tables S2, S5, S13 |
| Synthesis of results | 21 | Present results of each meta-analysis done, including confidence/credible intervals. *In larger networks, authors may focus on comparisons versus a particular comparator (e.g. placebo or standard care), with full findings presented in an appendix. League tables and forest plots may be considered to summarize pairwise comparisons.* If additional summary measures were explored (such as treatment rankings), these should also be presented. | Results - Findings per outcome; Table 3; Figures 3 and 5 |
| **Exploration for inconsistency** | **S5** | Describe results from investigations of inconsistency. This may include such information as measures of model fit to compare consistency and inconsistency models, *P* values from statistical tests, or summary of inconsistency estimates from different parts of the treatment network. | Results - Incoherence; Tables S21-S22; Figure S25 |
| Risk of bias across studies | 22 | Present results of any assessment of risk of bias across studies for the evidence base being studied. | Results - Risk of bias (summary); Certainty of evidence |
| Results of additional analyses | 23 | Give results of additional analyses, if done (e.g., sensitivity or subgroup analyses, meta-regression analyses*, alternative network geometries studied, alternative choice of prior distributions for Bayesian analyses,* and so forth). | Results - Subgroup findings; Tables S24-S37 |
| **DISCUSSION** |  |  |  |
| Summary of evidence | 24 | Summarize the main findings, including the strength of evidence for each main outcome; consider their relevance to key groups (e.g., healthcare providers, users, and policy-makers). | Discussion - paragraphs 1-3; Page 21-22 |
| Limitations | 25 | Discuss limitations at study and outcome level (e.g., risk of bias), and at review level (e.g., incomplete retrieval of identified research, reporting bias). *Comment on the validity of the assumptions, such as transitivity and consistency. Comment on any concerns regarding network geometry (e.g., avoidance of certain comparisons).* | Discussion – Limitations; paragraphs 22-23 |
| Conclusions | 26 | Provide a general interpretation of the results in the context of other evidence, and implications for future research. | Discussion - Conclusion section; Page 25 |
| **FUNDING** |  |  | Funding section; Page 25 |
| Funding | 27 | Describe sources of funding for the systematic review and other support (e.g., supply of data); role of funders for the systematic review. This should also include information regarding whether funding has been received from manufacturers of treatments in the network and/or whether some of the authors are content experts with professional conflicts of interest that could affect use of treatments in the network. |  |

PICOS = population, intervention, comparators, outcomes, study design.

* Text in italics indicateS wording specific to reporting of network meta-analyses that has been added to guidance from the PRISMA statement.

† Authors may wish to plan for use of appendices to present all relevant information in full detail for items in this section.

**Box. Terminology: Reviews With Networks of Multiple Treatments**

Different terms have been used to identify systematic reviews that incorporate a network of multiple treatment comparisons. A brief overview of common terms follows.

*Indirect treatment comparison:* Comparison of 2 interventions for which studies against a common comparator, such as placebo or a standard treatment, are available (i.e., indirect information). The direct treatment effects of each intervention against the common comparator (i.e., treatment effects from a comparison of interventions made within a study) may be used to estimate an indirect treatment comparison between the 2 interventions (**Appendix Figure 1, A**). An indirect treatment comparison (ITC) may also involve multiple links. For example, in **Appendix Figure 1, B**, treatments B and D may be compared indirectly on the basis of studies encompassing comparisons of B versus C, A versus C, and A versus D.

*Network meta-analysis* or *mixed treatment comparison*: These terms, which are often used interchangeably, refer to situations involving the simultaneous comparison of 3 or more interventions. Any network of treatments consisting of strictly unclosed loops can be thought of as a series of ITCs (**Appendix Figure 1, A and B**). In mixed treatment comparisons, both direct and indirect information is available to inform the effect size estimates for at least some of the comparisons; visually, this is shown by closed loops in a network graph (**Appendix Figure 1, C**). Closed loops are not required to be present for every comparison under study. "Network meta-analysis" is an inclusive term that incorporates the scenarios of both indirect and mixed treatment comparisons.

*Network geometry evaluation:* The description of characteristics of the network of interventions, which may include use of numerical summary statistics. This does not involve quantitative synthesis to compare treatments. This evaluation describes the current evidence available for the competing interventions to identify gaps and potential bias. Network geometry is described further in **Appendix Box 4**.

**Appendix Box 1. The Assumption of Transitivity for Network Meta-Analysis-**

Methods for indirect treatment comparisons and network meta-analysis enable learning about the relative treatment effects of, for example, treatments A and B through use of studies where these interventions are compared against a common therapy, C.

When planning a network meta-analysis, it is important to assess patient and study characteristics across the studies that compare pairs of treatments. These characteristics are commonly referred to as *effect modifiers* and include traits such as average patient age, gender distribution, disease severity, and a wide range of other plausible features.

For network meta-analysis to produce valid results, it is important that the distribution of effect modifiers is similar, for example, across studies of A versus B and A versus C. This balance increases the plausibility of reliable findings from an indirect comparison of B versus C through the common comparator A. When this balance is present, the assumption of transitivity can be judged to hold.

Authors of network meta-analyses should present systematic (and even tabulated) information regarding patient and study characteristics whenever available. This information helps readers to empirically evaluate the validity of the assumption of transitivity by reviewing the distribution of potential effect modifiers across trials.

**Appendix Box 2. Differences in Approach to Fitting Network Meta-Analyses**

Network meta-analysis can be performed within either a frequentist or a Bayesian framework. Frequentist and Bayesian approaches to statistics differ in their definitions of probability. Thus far, the majority of published network meta-analyses have used a Bayesian approach.

Bayesian analyses return the posterior probability distribution of all the model parameters given the data and prior beliefs (e.g., from external information) about the values of the parameters. They fully encapsulate the uncertainty in the parameter of interest and thus can make direct probability statements about these parameters (e.g., the probability that one intervention is superior to another).

Frequentist analyses calculate the probability that the observed data would have occurred under their sampling distribution for hypothesized values of the parameters. This approach to parameter estimation is more indirect than the Bayesian approach.

Bayesian methods have been criticized for their perceived complexity and the potential for subjectivity to be introduced by choice of a prior distribution that may affect study findings. Others argue that explicit use of a prior distribution makes transparent how individuals can interpret the same data differently. Despite these challenges, Bayesian methods offer considerable flexibility for statistical modeling.

In-depth introductions to Bayesian methods and discussion of these and other issues can be found elsewhere.

### Why didn’t we used Bayesian

Vague prior, what prior

Always test Bayesian with frequentist

**Appendix Box 3. Network Meta-Analysis and Assessment of Consistency**

Network meta-analysis often involves the combination of direct and indirect evidence. In the simplest case, we wish to compare treatments A and B and have 2 sources of information: direct evidence via studies comparing A versus B, and indirect evidence via groups of studies comparing A and B with a common intervention, C. Together, this evidence forms a closed loop, ABC.

Direct and indirect evidence for a comparison of interventions should be combined only when their findings are similar in magnitude and interpretation. For example, for a comparison of mortality rates between A and B, an odds ratio determined from studies of A versus B should be similar to the odds ratio comparing A versus B estimated indirectly based on studies of A versus C and B versus C. This assumption of comparability of direct and indirect evidence is referred to as *consistency* of treatment effects.

When a treatment network contains a closed loop of interventions, it is possible to examine statistically whether there is agreement between the direct and indirect estimates of intervention effect.

Different methods to evaluate potential differences in relative treatment effects estimated by direct and indirect comparisons are grouped as *local approaches* and *global approaches.* Local approaches (e.g., the Bucher method or the node-splitting method) assess the presence of inconsistency for a particular pairwise comparison in the network, whereas global approaches (e.g., inconsistency models, *I*^2^ measure for inconsistency) consider the potential for inconsistency in the network as a whole.

Tests for inconsistency can have limited power to detect a true difference between direct and indirect evidence. When multiple loops are being tested for inconsistency, one or a few may show inconsistency simply by chance. Further discussions of consistency and related concepts are available elsewhere.

Inconsistency in a treatment network can indicate lack of transitivity (see **Appendix Box 1**).

**Appendix Box 4. Network Geometry and Considerations for Bias**

The term *network geometry* is used to refer to the architecture of the treatment comparisons that have been made for the condition under study. This includes what treatments are involved in the comparisons in a network, in what abundance they are present, the respective numbers of patients randomly assigned to each treatment, and whether particular treatments and comparisons may have been preferred or avoided.

Networks may take on different shapes. Poorly connected networks depend extensively on indirect comparisons. Meta-analyses of such networks may be less reliable than those from networks where most treatments have been compared against each other.

Qualitative description of network geometry should be provided and accompanied by a network graph. Quantitative metrics assessing features of network geometry, such as *diversity* (related to the number of treatments assessed and the balance of evidence among them), *co-occurrence* (related to whether comparisons between certain treatments are more or less common), and *homophily* (related to the extent of comparisons between treatments in the same class versus competing classes), can also be mentioned.

Although common, established steps for reviewing network geometry do not yet exist, however examples of in-depth evaluations have been described related to treatments for tropical diseases and basal cell carcinoma and may be of interest to readers. An example based on 75 trials of treatments for pulmonary arterial hypertension (**Appendix Figure 3**) suggests that head-to-head studies of active therapies may prove useful to further strengthen confidence in interpretation of summary estimates of treatment comparisons.

**Appendix Box 5. Probabilities and Rankings in Network Meta-Analysis**

Systematic reviews incorporating network meta-analyses can provide information about the hierarchy of competing interventions in terms of treatment rankings.

The term *treatment ranking probabilities* refers to the probabilities estimated for each treatment in a network of achieving a particular placement in an ordering of treatment effects from best to worst. A network of 10 treatments provides a total of 100 ranking probabilities—that is, for each intervention, the chance of being ranked first, second, third, fourth, fifth, and so forth).

Several techniques are feasible to summarize relative rankings, and include graphical tools as well as different approaches for estimating ranking probabilities. **Appendix Figure 6** shows 2 approaches to presenting such information, on the basis of a comparison of adjuvant interventions for resected pancreatic adenocarcinoma.

Robust reporting of rankings also includes specifying median ranks with uncertainty intervals, cumulative probability curves, and the surface under the cumulative ranking (SUCRA) curve.

Rankings can be reported along with corresponding estimates of pairwise comparisons between interventions. Rankings should be reported with probability estimates to minimize misinterpretation from focusing too much on the most likely rank.

Rankings may exaggerate small differences in relative effects, especially if they are based on limited information. An objective assessment of the strength of information in the network and the magnitude of absolute benefits should accompany rankings to minimize potential biases.

**Appendix Figure 1A-1C**

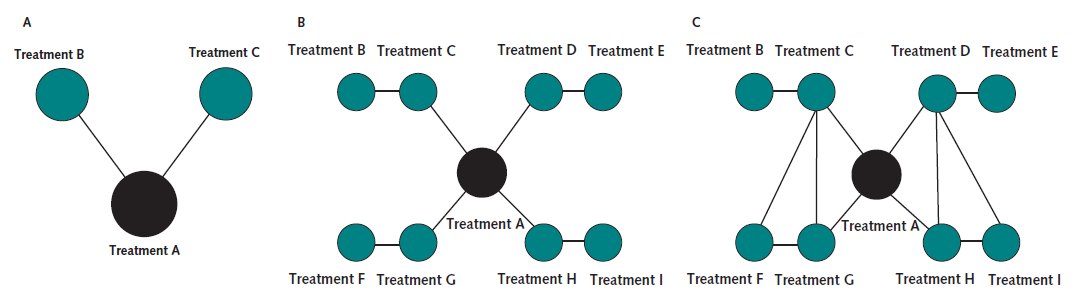

**Appendix Figure 3**

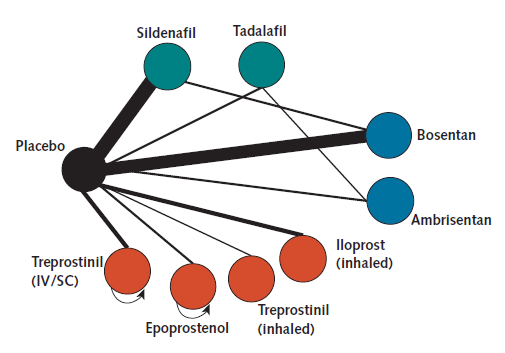

**Appendix Figure 6**

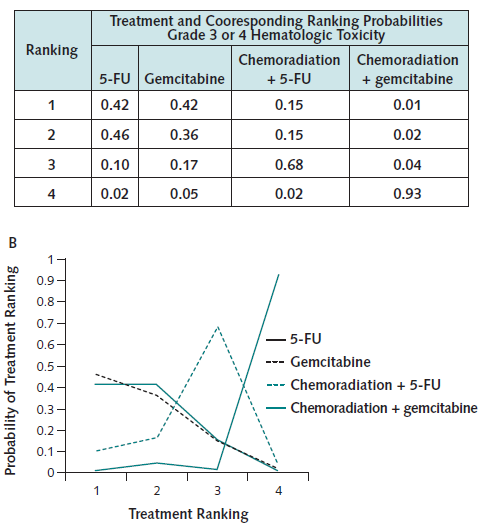

References

1.	*<Bucher 1997- the results of direct and indirect.pdf>.*
